# How do the indices based on the EAT-Lancet recommendations measure adherence to healthy and sustainable diets? A comparison of measurement performance in adults from a French national survey

**DOI:** 10.1101/2024.05.14.24307335

**Authors:** Agustín R. Miranda, Florent Vieux, Matthieu Maillot, Eric O. Verger

**Affiliations:** MoISA, Univ Montpellier, CIHEAM-IAMM, CIRAD, INRAE, Institut Agro, IRD, Montpellier, France; MS-Nutrition, Marseille, France

## Abstract

Measuring adherence to EAT-Lancet recommendations for healthy and sustainable diets is challenging, leading to diverse methods and a lack of consensus on standardized metrics. Available indices vary mainly in scoring systems, food components, units, energy adjustments, and cut-off points. We aimed to evaluate and compare the measurement performance of six dietary indices for assessing adherence to EAT-Lancet reference diet. Food consumption data of 1,723 adults were obtained from the French Third Individual and National Study on Food Consumption Survey (INCA3, 2014-2015). Sociodemographic, nutritional, and environmental data were used to assess the validity and reliability of dietary indices. Results showed that the four indices assessing their food components with quantitative scoring captured dietary variability, were less dependent on energy intake and converged to a large extent with nutritional indicators. While the two binary indices showed a stronger correlation with environmental indicators, one quantitative index converged with both domains. Indices had valid unidimensional structures, meaning that the combination of food components within each index accurately reflects the same construct and supporting the use of total scores. Furthermore, the indices differed between sociodemographic groups, demonstrating concurrent criterion validity. Higher scores were associated with higher nutritional quality and lower environmental impact, but with unfavourable results for zinc intake, vitamin B12 and water use. A low concordance rate (from 32% to 43%) indicated that indices categorized individuals differently. Researchers must align study objectives with the applicability, assumptions, and functional significance of chosen indices. Indices using quantitative scoring allow a global understanding of dietary health and sustainability, being advantageous in precision-focused research, such as clinical trials or epidemiological research. Conversely, indices based on binary scoring offer a simplified perspective, being valuable tools for surveys, observational studies, and public health. Recognizing their strengths and limitations is crucial for a comprehensive assessment of diets and understanding their implications.

## 1. Introduction

Currently, global policy agenda emphasizes nutritional strategies focused on supplying vital nutrients, reducing environmental impact, and advancing long-term sustainability [1]. Organizations such as the Food and Agriculture Organization (FAO) and the World Health Organization (WHO) call for healthy and sustainable food patterns that are accessible, affordable, safe, equitable and culturally acceptable [2]. This issue aligns with the 17 United Nations’ Sustainable Development Goals (SDG) targeting hunger eradication, improved nutrition, and food system sustainability [3]. However, meeting the goals of operating within planetary boundaries and promoting healthy diets remains a challenge. Planetary boundaries are essential environmental limits that must be respected to maintain the global balance and overall functioning of the Earth system, but current food systems, particularly food production practices, threaten these limits significantly [4]. Food systems include all the aspects of feeding and nourishing people, from production to consumption, involving multiple resources and their impact on nutrition, health, economy, society and environment [5]. Food systems contribute significantly to climate change with 34% of greenhouse gas emissions (GHGE) [6], 70% of freshwater consumption, leading to resource depletion, and contribute to pollution, land use, and biodiversity loss [7].

To advance the achievement of the SDGs and Paris Climate Agreement commitments, the EAT-Lancet Commission introduced a planetary health diet in 2019 as a global standard for adults [7]. This reference diet, based on 2500 kcal per day for a 70 kg 30-year-old man or 60 kg 30-year-old woman with moderate to high physical activity, sets ranges for specific food groups to promote healthy eating and sustainable food production. In this regard, the planetary health diet prioritizes the consumption of vegetables, fruits, legumes, whole grains, nuts and fish, while limiting the intake of red meat and tubers. It also promotes moderate consumption of eggs, poultry, and dairy products [7]. While this diet can serve as benchmarks, criticisms include impracticality for poor settings, and its adult-focused targets may not apply directly to vulnerable groups [8].

Upon the release of the EAT-Lancet report, measuring adherence to the planetary health diet faced challenges, leading to diverse methods without consensus in their development [9]. Early instruments assess adherence using binary scoring (i.e., for each food component within the index, a score of 1 is assigned when meeting the recommendation and 0 for not meeting). Among these are the EAT-Lancet Diet Score (ELDS) and the Healthy and Sustainable Diet Index (HSDI), based on data from the United Kingdom and Mexico, respectively [10,11]. Although they are associated with health outcomes, the validity of both indices has not been explored. Recent indices have refined their designs by incorporating quantitative scoring, adjustments for energy intake, and interchangeability between food components. Among these, the World Index for Sustainability and Health (WISH) was developed using data from Vietnam and is positively associated with health indicators [12]. The Planetary Health Diet Index (PHDI), is based on data from a large Brazilian cohort and is related to cardiovascular, nutritional and environmental indicators [13–15]. Likewise, the EAT-Lancet Dietary Index (ELD-I), developed with data from France, shows a positive correlation with nutritional quality and a negative correlation with environmental impact [16]. Finally, the EAT-Lancet Index (ELI), developed with Swedish data, shows associations with reduced mortality and a lower risk of chronic diseases [17,18]. Despite notable progress in the development of these indices, there are still gaps on their validity, and they have not yet been comprehensively compared using data from the same sample.

While efforts have been made to develop methods for measuring adherence to EAT-Lancet diet, further research is needed for the assessment of various measurement properties to ensure their validity and reliability [19,20]. Some indices like PHDI and ELD-I have undergone validation, but most have not reported their validity indicators. This is crucial because many indices may lack representativity due to study design, participant characteristics, or sample size. Dietary indices, particularly those based on EAT-Lancet diet, require validation using nutritional and environmental indicators. When selecting a dietary index, aligning it with research goals, comprehending the scoring system, and ensuring a robust and unbiased validation process is crucial to enhance reliability in nutritional epidemiology [21].

In this context, the aim of this study was to assess and compare the measurement performance, focusing on the aspects of validity and reliability, of six dietary indices representing the EAT-Lancet reference diet using a national representative sample from France.

## 2. Materials and methods

### 2.1. Population

Data were extracted from the French Third Individual and National Study on Food Consumption Survey (INCA3). The INCA3 is a nationally representative cross-sectional survey conducted on 4,114 individuals in mainland France between February 2014 and September 2015. The methodology and study design of this survey are described in detail elsewhere [22]. In the present study, participants aged ≥ 18 years old were included and mis-reporters were excluded, using the basal metabolic rate estimated using the Henry equation and the cut-off values proposed by Black [23,24]. Thus, the final sample contained 1,723 adults (723 men and 1,000 women). The flow chart of study participants is detailed in Supplementary Material (Fig S1).

The INCA3 study was carried out in accordance with the Declaration of Helsinki and received approval from the French Data Protection Authority (Decision DR 2013–228) on May 2, 2013, following a favorable opinion from the Advisory Committee on Information Processing in Health Research on January 30, 2013 (Opinion 13.055). Verbal informed consent was obtained from all participants before their voluntary inclusion in the study. Verbal consent was witnessed and formally recorded.

### 2.2. Dietary data

Dietary intake was assessed through three non-consecutive dietary recalls, two weekdays and one weekend, over a three weeks period. Participants were contacted by phone to declare all the foods and beverages they had consumed during the previous day using validated photographs of standard portion sizes in France [22]. The interview days were not disclosed to the participants to prevent them from predicting and adjusting their food intake. Energy and nutrient contents of foods were based on the 2016 database from the French Centre d’Information sur la Qualité des Aliments [25]. The traditional recipes or dishes containing various foods were disaggregated into their ingredients based on average recipes obtained from an existing recipe database [20], and on recipes sourced from the most popular cooking website in France (i.e., marmiton.org) [26]. The dietary data were used to calculate EAT-Lancet adherence indices, nutritional quality and environmental scores for each individual from the INCA3.

### 2.3. Estimation of indices of adherence to the EAT-Lancet Diet

#### 2.3.1. World Index for Sustainability and Health (WISH)

The WISH is an index based on a quantitative scoring consisting of 13 food components [12]. Each food component is scored on a scale ranging from 0 (non-compliance) to 10 (full compliance) points, using reference values in grams based on both the healthiness and environmental sustainability of the food component. Subsequently, the scores for the food components are summed to calculate the total score, with the total score ranging from 0 to 130 (the higher, the greater adherence to a healthy and sustainable diet). Concerning saturated fats and added sugars, both components are scored using a binary scoring: 10 points if consumption is equal to or below the recommended intake and 0 points if it exceeds it. More information about the WISH index is available elsewhere [12].

#### 2.3.2. Planetary Health Diet Index (PHDI)

The PHDI is an index comprised of 16 food components that are scored using a quantitative system based on reference values expressed as ratios of energy intake [13]. Energy intake ratios are defined as the sum of calories from all foods in a food component divided by the total calories from all foods in the PHDI index (i.e., energy within a food component in the numerator, while the sum the energy of all foods included in the PHDI in the denominator). Each food component was categorized either in an adequacy component (nuts and peanuts, fruits, legumes, vegetables, and whole grain cereals), optimum component (eggs, dairy products, fish and seafood, tubers and potatoes, and vegetable oils), moderation component (red meat, chickens and substitutes, animal fats, and added sugars) or ratio component (dark green vegetables/total vegetables and red–orange vegetables/total vegetables). Adequacy, moderation and optimum components could score a maximum of 10 points, while the ratio ones could score a maximum of 5 points. The sum of these components results in a total score that ranges from 0 to 150 points. Higher scores indicate greater adherence to the EAT-Lancet diet. More information is available elsewhere [13].

#### 2.3.3. EAT-Lancet Diet Index (ELD-I)

The ELD-I index assesses the proximity of a diet to the EAT-Lancet reference for 14 food components using a quantitative scoring [16]. ELD-I calculations are adjusted by individual energy intake, with a reference of 2,500 kcal. Additionally, this index distinguishes between different food components, awarding positive scores when the consumption of recommended foods exceeds the reference and when the consumption of foods to limit is below it.

Conversely, it assigns negative scores when the consumption of recommended foods is below the reference and when the consumption of foods to limit exceeds the reference. As a result, ELD-I calculation yields a continuous unbounded score that can be either positive or negative. Higher scores reflect a greater alignment with the EAT-Lancet recommendations. More information about the development of ELD-I index is available elsewhere [16].

#### 2.3.4. EAT-Lancet Index (ELI)

The ELI index consists of 14 food components divided into two groups: 7 positive components or “emphasized foods” and 7 negative components or “limited foods” [17]. The alignment of dietary intake in grams per day (without adjustment on daily energy) with the EAT-Lancet recommendations is assessed using a scoring system based on a semi-quantitative scale ranging from 0 (non-compliance) to 3 points (high compliance). As a result, the total scores of the ELI dietary index range from 0 to 42 points. More information about the ELI index, scoring criteria, and cut-offs points, is available elsewhere [17].

#### 2.3.5. Healthy and Sustainable Diet Index (HSDI)

The HSDI assesses the degree of compliance with EAT-Lancet recommendations through a binary scoring [11]. This index analyzes the percentage of energy intake from 13 food components (based on a daily energy of 2,500kcal), assigning one point when the recommended energy percentage is met and zero points otherwise. The food components are then summed, resulting in a scale ranging from 0 to 13 points, where a higher score reflects greater adherence to EAT-Lancet recommendations. More information about the HSDI index is available elsewhere [11].

#### 2.3.6. EAT-Lancet Diet Score (ELDS)

The ELDS index consists of 14 food components and relies on a binary scoring [10]. One point is assigned to each component for meeting each of the recommended intakes in terms of grams per day without energy adjustment. The sum of all components results in a total score that ranges from 0 to 14 points. Higher scores indicate a greater level of adherence to the EAT-Lancet recommendations. More information about the development of ELDS index, scoring criteria, and cut-offs points, is available elsewhere [10].

Table 1 displays the correspondences between the food components and the reference values for the six indices based on EAT-Lancet recommendations. Supplementary Material lists the food items, scoring criteria, and cut-offs points that were considered in each food component.

**Table 1.** Equivalences between the food components of the indices based on the EAT-Lancet recommendations and their scoring standards.

| Quantitative scoring |  |  |  |  |  | Semi-quantitative scoring |  | Binary scoring |  |  |  |
| --- | --- | --- | --- | --- | --- | --- | --- | --- | --- | --- | --- |
| WISH<sup>a</sup><br>from 0 to 130<br>13 food components |  | PHDI<sup>b</sup><br>from 0 to 150<br>16 food components |  | ELD-I<sup>a</sup><br>from -∞ to +∞<br>14 food components |  | ELI<sup>a</sup><br>from 0 to 42<br>14 food components |  | HSDI<sup>b</sup><br>from 0 to 13<br>13 food components |  | ELDS<sup>a</sup><br>from 0 to 14<br>14 food components |  |
| Whole grains | ≥125 (100-150) | Whole cereals | ≥32.4 | Whole grains | ≤464 | Whole grains | 232 | Whole grains | ≥32.44 | Whole grains | ≤464 |
| — |  | Tubers and potatoes | 1.6 (0-3.1) | Potatoes and tuber | ≤100 | Potatoes | 50 (0-100) | Tubers or starchy vegetables | ≤1.56 | Tubers and starchy vegetables | ≤100 |
| Vegetables | 300 (200-600) | Vegetables | ≥3.1 | Vegetables | ≥200 | Vegetables | 300 (200-600) | Vegetables | ≥3.12 | Vegetables | ≥200 |
| Fruits | 200 (100-300) | Fruits | ≥5.0 | Fruits | ≥100 | Fruits | 200 (100-300) | Fruits | ≥5.02 | Fruits | ≥100 |
| Dairy foods | 250 (0-500) | Dairy | 6.1 (0-12.2) | Dairy foods | ≤500 | Dairy | 250 (0-500) | Milk and dairy | ≤6.12 | Dairy foods | ≤500 |
| Red meat | 14 (0-28) | Red meat | 0 (0-2.4) | Beef, lamb, pork | ≤28 | Beef and lamb | 7 (0-14) | Beef and pork | ≤0.64 | Beef, lamb, pork | ≤28 |
|  |  |  |  |  |  | Pork | 7 (0-14) |  |  |  |  |
| Chicken and other poultry | 29 (0-58) | Chicken and substitutes | 0 (0-5.0) | Chicken and poultry | ≤58 | Poultry | 29 (0-58) | Chicken and other poultry | ≤2.48 | Chicken, other poultry | ≤58 |
| Eggs | 13 (0-25) | Eggs | 0.8 (0-1.5) | Eggs | ≤25 | Eggs | 13 (0-25) | Eggs | ≤1.00 | Eggs | ≤25 |
| Fish | 28 (0-100) | Fish and seafood | 1.6 (0-5.7) | Fish | ≤100 | Fish | 28 (0-100) | Fish and seafood | ≤1.60 | Fish | ≤100 |
| Legumes | 75 (0-100) | Legumes | ≥11.3 | Legumes | ≤100 | Legumes | 75 (0-150) | Legumes, soybeans and tree nuts | ≥23.0 | Dry beans, lentils, peas | ≤100 |
| Nuts | 50 (0-75) | Nuts and peanuts | ≥11.6 | Nuts | ≥25 | Nuts | 50 (0-100) |  |  | Peanuts or tree nuts | ≥25 |
| Unsaturated oils | 40 (20-80) | Vegetable oils | 16.5 (0-30.7) | Unsaturated oils | ≤80 | Unsaturated oils | 40 (20-80) | Unsaturated fats | ≥14.16 | Added fats | 0.8 |
| Saturated oils | 11.8 (0-11.8) | Animal fats | 0 (0-1.4) | Saturated oils | ≤11.8 |  |  | Saturated fats | ≤3.84 |  |  |
| Added sugars | 31 (0-31) | Added sugars | 0 (0-4.8) | Added sugars | ≤31 | Added sugar | 31 (0-31) | Added sugars | ≤5.00 | Added sugar | ≤31 |
| — |  | DGV/total ratio | 29 (0-100) | — |  | — |  | — |  | — |  |
| — |  | ReV/total ratio | 38.5 (0-100) | — |  | — |  | — |  | — |  |
| — |  | — |  | — |  | — |  | — |  | Soy foods | ≤50 |

*Note.* WISH = World Index for Sustainability and Health; PHDI = Planetary Health Diet Index; ELD-I = EAT-Lancet Diet Index; ELI = EAT-Lancet index; HSDI = Healthy and Sustainable Diet Index; ELDS = EAT-Lancet Diet Score; a = values expressed as g/d; b = values expressed as caloric percentage from the reference diet proposed by the EAT-Lancet Commission; DGV/total ratio = ratio between the energy intake of dark green vegetables and the total of vegetables; ReV/total ratio = ratio between the energy intake of red and orange vegetables and the total of vegetables. The reference intervals, applicable to certain indexes in their calculations, are presented within parentheses.

### 2.4. Nutritional quality assessments

#### 2.4.1. Assessment of nutrient adequacy

The PANDiet quality index, used for evaluating nutritional adequacy [27], is a comprehensive measure assessing the probability of meeting recommended intake levels for 33 nutrients. Comprising two sub-scores – “adequacy” and “moderation” – the former involves averaging the probabilities for 27 nutrients, while the latter averages probabilities for 6 nutrients to be kept within upper limits. Nutrients for adequacy sub-score include proteins, total carbohydrates, dietary fibre, total fats, 4 essential fatty acids, 11 vitamins, and 10 minerals. In contrast, moderation sub-score focuses on limiting intake of proteins, total carbohydrates, total sugars, total fats, saturated fatty acids, and sodium. Each sub-score undergoes multiplication by 100, followed by the calculation of their average. This process yields the total PANDiet score, which ranges from 0 to 100. The PANDiet metrics are expressed as likelihood of adequacy, with higher scores implying superior diet quality and greater nutrient adequacy. The dietary reference values used to calculate PANDiet were based on guidelines issued by the French Agency for Food, Environmental and Occupational Health and Safety (ANSES) (see Supplementary Material, Table S7).

#### 2.4.2. Global Diet Quality Score (GDQS)

The GDQS is a recently developed score designed to assess diet quality based on 25 food categories considered critical for nutrient supply and chronic disease risk reduction [28]. This score is composed of 16 healthy food components (higher intake results in a higher score), 7 unhealthy food components (higher intake results in a lower score), and 2 food components considered unhealthy when consumed in excess (resulting in a score of 0 for both insufficient and excessive intake). The total GDQS score is calculated by adding up the points assigned to the 25 food components, and ranges from 0 to 49 points. It is further divided down into two sub-scores: GDQS+, which reflects the sum of the healthy food components with a range between 0 and 32, and GDQS-, which is based on the unhealthy or over-consumed food components, with a score ranging from 0 to 17.

#### 2.4.3. Comprehensive Diet Quality Index (cDQI)

The cDQI discriminates between the intake of plant and animal food components considered beneficial or detrimental to health [29]. The cDQI is composed of 11 components of plant-based foods (pDQI) and 6 components of animal-based foods (aDQI). Healthy foods receive positive scores, in contrast to unhealthy foods that receive negative scores. The total cDQI score ranges from 0 to 85.

#### 2.4.4. Dietary Inflammatory Index (DII)

The DII was used to assess the inflammatory potential of the diet [30]. The DII was developed following an extensive literature review that identified 45 dietary parameters (foods or nutrients) associated with six inflammatory biomarkers: interleukin (IL)-1β, IL-4, IL-6, IL-10, tumour necrosis factor (TNF)-α, and C-reactive protein. In this study, a total of 34 out of the possible dietary parameters were used to calculate the DII. The computation includes comparing dietary intake with the global standard, calculating Z-scores, converting them to centred proportions, multiplying by inflammatory effect scores for each parameter, and summing to get the total DII score (i.e., a higher DII indicates a more proinflammatory diet) [31]. The specific steps for the calculation are available in Supplementary Material.

#### 2.4.5. Composite Dietary Antioxidant Index (CDAI)

The CDAI was calculated with the aim of assessing the overall exposure to dietary antioxidants [32]. The CDAI is a score that integrates various dietary antioxidants, including vitamins A, C, and E, manganese, selenium, and zinc, and it reflects an individual’s dietary antioxidant profile. To obtain the CDAI, each of these six dietary antioxidants was standardized by subtracting the sex-specific mean and dividing by the sex-specific standard deviation, and then they were summed up. The higher the CDAI, the greater the bioavailability of antioxidants in the diet, suggesting a potentially higher level of defence against oxidative stress and health protection [33].

### 2.5. Environmental data

The analysis of environmental impact was carried out using the Agribalyse 3.1.1 database, which was developed by the French Agency for the Environment and Energy Management [34]. Agribalyse 3.1.1 provides reference data on the environmental impacts of agricultural and food products through a database constructed using the Life Cycle Assessment methodology, considering different stages of the food chain. In the current study, the aggregated score product environmental footprint (PEF) and the following 14 metrics were used: greenhouse gas emissions (kg CO2 eq), exposure ionizing radiation (kg U235 eq), photochemical ozone formation (kg NMVOC eq), ozone depletion (Freon-11), emission of particulate matter in change (mortality due to particulate matter emissions), acidification (mol H+ eq), terrestrial eutrophication (mol N eq), freshwater eutrophication (kg P eq), marine eutrophication (kg N eq), freshwater ecotoxicity in (CTUe), water use (m3 world eq), land use (loss of soil organic matter content in kg carbon deficit), fossils resource use (MJ), metals and minerals resource use (kg Sb eq). The complete methodology of Agribalyse 3.1.1 is explained elsewhere [35].

### 2.6. Other data

Sociodemographic variables included sex (woman or man), age group (18–44 years, 45–64 years, or 65–79 years), educational level (primary and middle school, high school, 1 to 3 years of post-secondary education, or ≥4 years of post-secondary education), income per consumption unit (< €900/month/CU, €900–€1340/month/CU, €1340–€1850/month/CU, or ≥ €1850/month/CU), weight status according to WHO body mass index categories (underweight, normal, overweight, obesity, and morbid obesity), smoking habit (smoker or non-smoker), and level of physical activity assessed by an adapted version of the Recent Physical Activity Questionnaire (categorized as low, moderate, or high) [22,36]. Additionally, the ratio between PANDiet score and PEF was calculated to combine both nutritional adequacy and global environmental indicator into a single indicator.

### 2.7. Statistical analysis

Statistical analyses were performed using Stata (version 18, StataCorp, College Station, TX, USA) and the threshold for statistical significance was p[<[0.05. The weighting factors supplied within the INCA3 survey were employed in the analyses to address the complex survey design and to guarantee national representativeness [22]. Mean and standard deviation (SD) were calculated for numerical variables, and percentages for categorical ones. Q-Q plot and Kernel density histograms were used to assess the distribution of the indices against normal distribution. Correlation coefficients were interpreted following established guidelines, where coefficients below 0.2, between 0.2 and 0.39, 0.4 and 0.59, 0.6 and 0.79, and 0.8 to 1 indicate very weak, weak, moderate, strong, and very strong associations, respectively [37].

#### 2.7.1. Assessment of reliability and structure validity of indices

To assess homogeneity of food components within each score, correlations between them (i.e., inter-component correlations) were calculated using different methods according to the data to test how each food component behaves individually with respect to the others. Pearson’s correlation was used for quantitative indices, polychoric correlation for those using a semi-quantitative scale, and tetrachoric correlation for dichotomous data. Also, Pearson’s correlation and point-biserial correlation were used to calculate the relationships between each component and the total score (i.e., component-total correlations), with values above 0.80 considered as redundant.

Internal consistency reliability is a measure of the extent to which the food components in a dietary index measure the same underlying construct. The internal consistency reliability was assessed by calculating Guttman’s lambda (λ) coefficients. These coefficients encompass six measures (λ_1_-λ_6_) based on the analysis of the total variance of the index, the variance of each of its components, and the covariance between them. Briefly, to calculate each λ coefficient, the three parameters are set differently, such as summing the variance or computing the variance on a per-component basis, and different adjustments are applied, considering factors like the number of food components [38]. Special attention is focused on λ_4_, which represents the maximum split-half reliability and measures how all parts of an instrument contribute equally to what is being measured.

Structural equation modelling (SEM) was used to test the structural validity of the indices. Maximum likelihood estimation was adopted to determine the model fit when predicting the correlations among food components (observed variables) through a single underlying continuous latent variable (total index score). Recommended goodness-of-fit indices were calculated: Chi-square to degree of freedom ratio (χ2/df), root mean square error of approximation (RMSEA), comparative fit index (CFI), standardised root mean square residual (SRMR), coefficient of determination (CD). For HSDI and ELDS, generalised structural equation modelling (GSEM) was used, as they are based on a binary scoring, thus goodness-of-fit indices could not be calculated.

#### 2.7.2. Index variability and relationship with total energy intake

The percentile distribution, ranging from the 1st to the 99th percentile, was computed for the six indices to assess their ability to capture dietary variability, which is crucial for nutritional metrics as it indicates sensitivity to detect sufficient data variation [19]. Likewise, Pearson’s coefficients were calculated between the indices and total energy intake (TEI) to test if these are independent of the TEI.

#### 2.7.3. Examination of concordance between indices

Furthermore, the degree of concordance among indices was analysed [39]. For this, we determined the proportion of participants classified in the same quintile, the adjacent quintile, and the opposite extreme quintile in relation to the indices based on the total score. Quartiles were employed instead of quintiles for binary indices due to the limited range of variation in total scores, which can result in imbalanced categories. Alluvial diagrams were used to visualize transitions between quintiles/quartiles. Additionally, Fleiss’ kappa (κ) coefficients were calculated, with a score of 1 indicating perfect concordance, while a score close to 0 suggests poor concordance between the indices. The κ coefficients were interpreted as follows: <0.00 as poor; 0.00-0.20 as slight; 0.21-0.40 as fair; 0.41-0.60 as moderate; 0.61-0.80 as substantial, and 0.81-1.0 as almost perfect concordance [40].

#### 2.7.4. Analyses of indices according to sociodemographic variables

Comparing different demographic groups with known differences allows for the evaluation of concurrent criterion validity (i.e., measurement performance when assessed against an external criterion) [19]. Therefore, the means of the total scores were compared across different sociodemographic groups using analysis of covariance (ANCOVA). Additionally, the Jonckheere-Terpstra trend test was conducted to examine the trend of the indices across demographic groups.

#### 2.7.5. Analysis of convergent validity using validated nutritional quality and environmental indicators

Measures related to a particular phenomenon are expected to be highly correlated, suggesting that they converge and are measuring the same underlying construct [20]. Therefore, since the six indices based on the EAT-Lancet recommendations have been proposed as a measure of healthy and sustainable diets, correlations with nutritional and environmental indicators were analysed by calculating Spearman’s coefficients (ρ).

#### 2.7.6. Analysis of trends across quintiles

A series of analyses of variance (ANOVA) were used to test the means of PANDiet and environmental impact indicators among quintiles/quartiles of the six indices. ANOVA effect sizes were expressed as partial eta-squared coefficients (η²) to describe the proportion of the total variation of score that can be attributed to each variable. The η2 cut-off points were as follows: 0.01 (small effect), 0.06 (moderate effect), and 0.14 (large effect). Trends were assessed using the Jonckheere-Terpstra test. Violin plots were used to visually illustrate the differences in PEF and PANDiet between the lowest and the highest quintiles/quartiles.

## 3. Results

### 3.1. Descriptive characteristics

The six indices presented a normal distribution (see Fig S2 in Supplementary Material). As for the WISH index, the score ranged from 2 to 97 points, with a mean of 40.42 points (SD = 15.50). Differences were observed in the scores for the food components (Fig 1). In particular, the chicken and other poultry, dairy foods and eggs food components obtained mean scores above five points. In contrast, whole grains, unsaturated oils, nuts and legumes were the groups with the lowest scores, all with means below one point.

**Fig 1.**
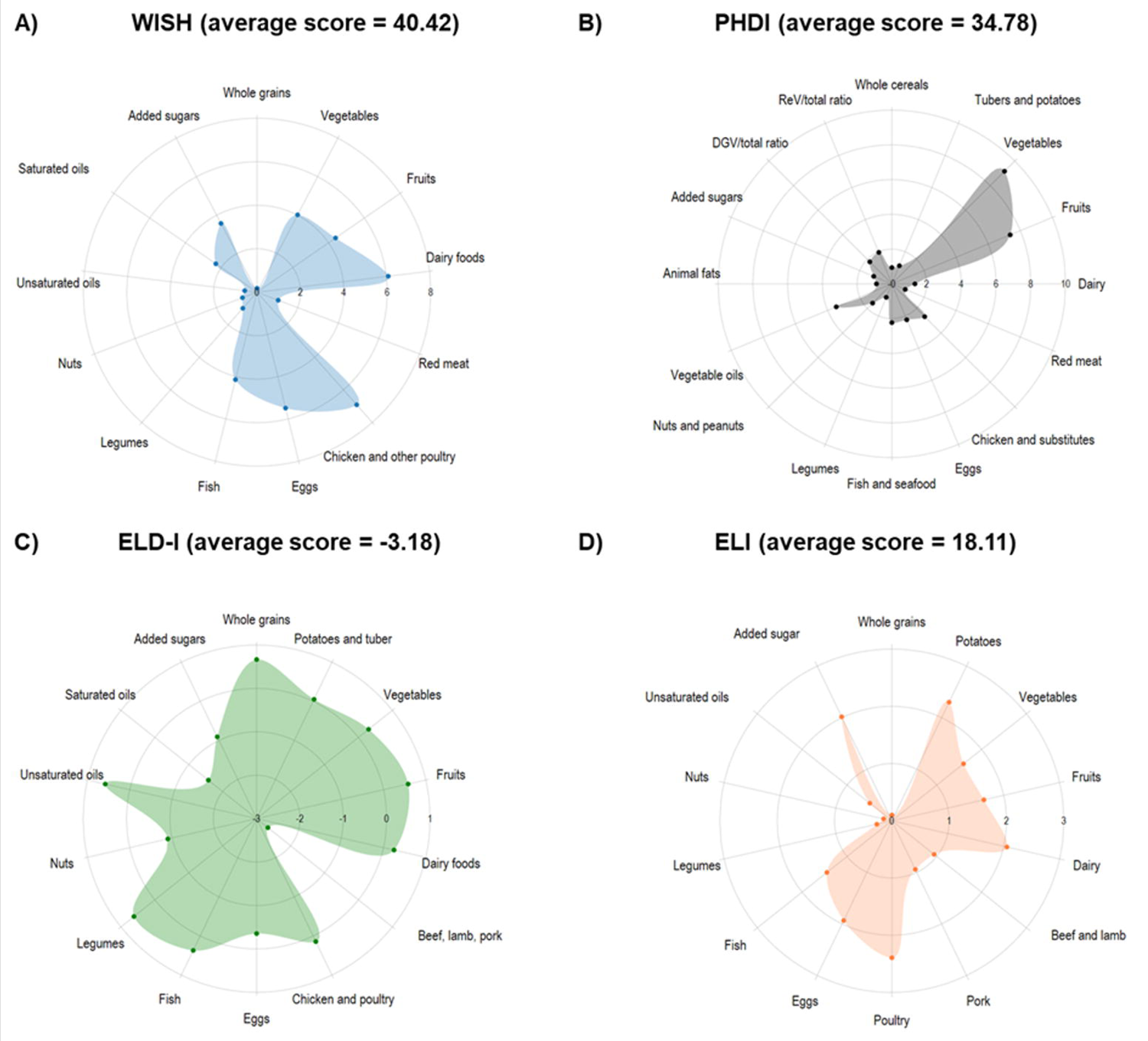
Mean Scores of the food components of EAT-Lancet-Based Indices in the French Third Individual and National Study on Food Consumption Survey (INCA3, n = 1,723).

Regarding the PHDI index, the mean total score was 34.78 points (SD = 11.53), within a range that varied between 2.89 and 78.60 The food components that obtained the highest scores were vegetables, fruits, vegetable oils, and chicken and substitutes, while the red meat, legumes, animal fats, whole cereals, tubers and potatoes, and added sugars components showed scores below one point (Fig 1).

Regarding the ELD-I index, the mean for the total score was −3.18 points (SD = 33.72), ranging between −113.11 and 104.55 points. The food components with the highest scores were unsaturated oils, whole grains, legumes, fish and fruits, while the lowest scores were for beef, lamb and pork, saturated oils, added sugars, nuts and eggs, all with negative scores (Fig 1).

In relation to the ELI index, the total score ranged from 7 to 32 points, with a mean of 18.11 (SD = 4.04). The food components with the highest scores were poultry, potatoes, dairy and added sugars, while whole grains, nuts, legumes, unsaturated oils, beef, and lamb and pork registered lower scores (Fig 1).

On the other hand, the indices based on a binary scoring showed a dissimilar behaviour. In this sense, the HSDI had a mean of 3.93 points (SD = 1.62) in a range that varied between 0 and 10 points. The components with the highest proportion of participants meeting the recommendations were fish and seafood, vegetables and chicken and other poultry, with over 50% compliance (Fig 2). In contrast, less than 1% of participants met the target intake for legumes, soybeans and tree nuts, and whole grain foods. For the ELDS, the mean was 8.10 points (SD = 1.49, range = 4 to 12), with dry beans, lentils and peas, soy foods, dairy foods, and fish being the groups with the highest target compliance (Fig 2). Conversely, peanuts and tree nuts, added fats and beef, lamb and pork were less compliant.

**Fig 2.**
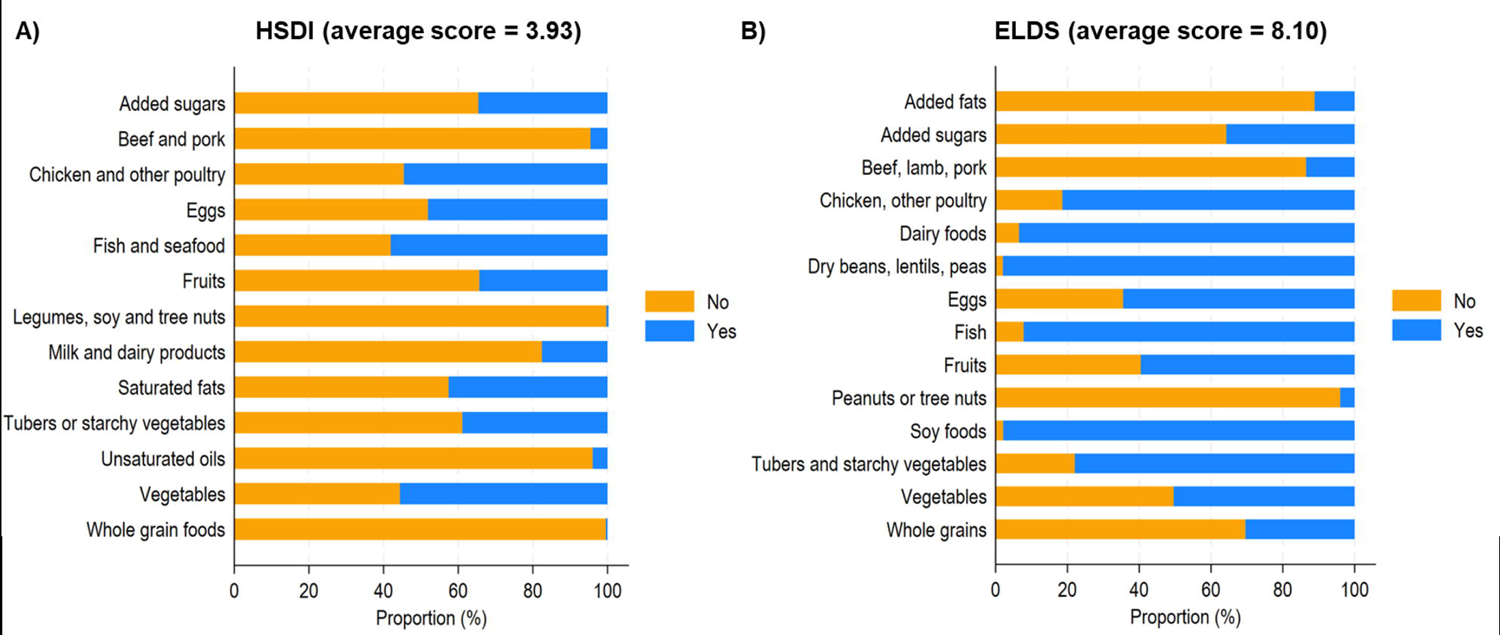
Proportion of participants adhering to EAT-Lancet Recommendations through binary indices in the French Third Individual and National Study on Food Consumption Survey (INCA3, n = 1,723).

### 3.2. Reliability and structural validity

ELD-I was the index with the highest λ coefficients, mainly for split-half reliability (λ4 = 0.57), followed by WISH (λ4 = 0.47), ELI (λ4 = 0.47) and PHDI (λ4 = 0.46). Conversely, the ELDS index had the lowest λ coefficients. Moreover, all inter-component correlations were below 0.80, suggesting an absence of redundancy. Further, all items contributed significantly to the total score as the component-total correlations were statistically significant, with the exception of chicken and poultry from the ELD-I, legumes from the HSDI, and soy foods from the ELDS. Curves with λ coefficients and correlation matrices are presented in Supplementary Material (Fig S3 and Tables S11-S16).

SEM models confirmed the unidimensional structural validity of the WISH, PHDI, ELD-I, and ELI indices (Table 2). Although the indices showed a similar fit profile, the PHDI and ELD-I were the most robust in explaining the data variability (CD = 0.568 and 0.466, respectively) and had high incremental indices (CFI > 0.90). Because HSDI and ELDS are composed of dichotomous items, they were modelled by GSEM, which confirmed their unidimensional structure (results not shown).

**Table 2.** Fit indices for Confirmatory Factor Analysis.

|  | Expected values | WISH | PHDI | ELD-I | ELI |
| --- | --- | --- | --- | --- | --- |
| $\chi^2/df$ | 2.50 | 1.65 | 1.77 | 2.34 | 1.60 |
| RMSEA | <0.08 | 0.019 | 0.021 | 0.028 | 0.019 |
| CFI | $\geq 0.90$ | 0.922 | 0.904 | 0.911 | 0.928 |
| SRMR | <0.05 | 0.025 | 0.027 | 0.028 | 0.023 |
| CD | The higher, the better | 0.323 | 0.568 | 0.466 | 0.364 |

*Note.* $\chi^2/df$ : Chi-square to degree of freedom ratio; RMSEA: Root Mean Square Error of Approximation; CFI: Comparative Fit Index; SRMR: Standardized Root Mean Square Residual; CD: the higher, the better.

### 3.3. Capture of diet variability and energy independence

ELD-I presented the greatest difference in scores between the 1st percentile (−84.01) versus the 99th percentile (74.72), followed by WISH (6.92 vs. 75.44) and PHDI (10.31 vs. 63.62). HSDI and ELDS showed the lowest change across percentiles (Supplementary Material Fig S4). In addition, the correlation of the indices with total energy intake was analysed. Total energy intake showed significant but negligible correlations with ELD-I (r = −0.08) and PHDI (r = −0.09). Regarding the other indices, correlations with energy intake were low (p < 0.0001): for HSDI r = −0.23, for WISH r = −0.25, for ELI r = −0.28, and for ELDS r = −0.30. More details are available in Table S17 (Supplementary Material).

### 3.4. Inter-index concordance

The alluvial plots in Fig 3 show the concordance among the indices. Total concordance (i.e., individuals classified in the same quintile/quartile) was below 50% for all paired comparisons. Moreover, between 23 and 28% of the participants were classified in adjacent quintiles/quartiles. The classification percentages in the opposite extreme quintile/quartile ranged from 1 to 7.5 %. Moreover, κ coefficients indicated slight or fair concordance between the indices (Fig 3).

**Fig 3.**
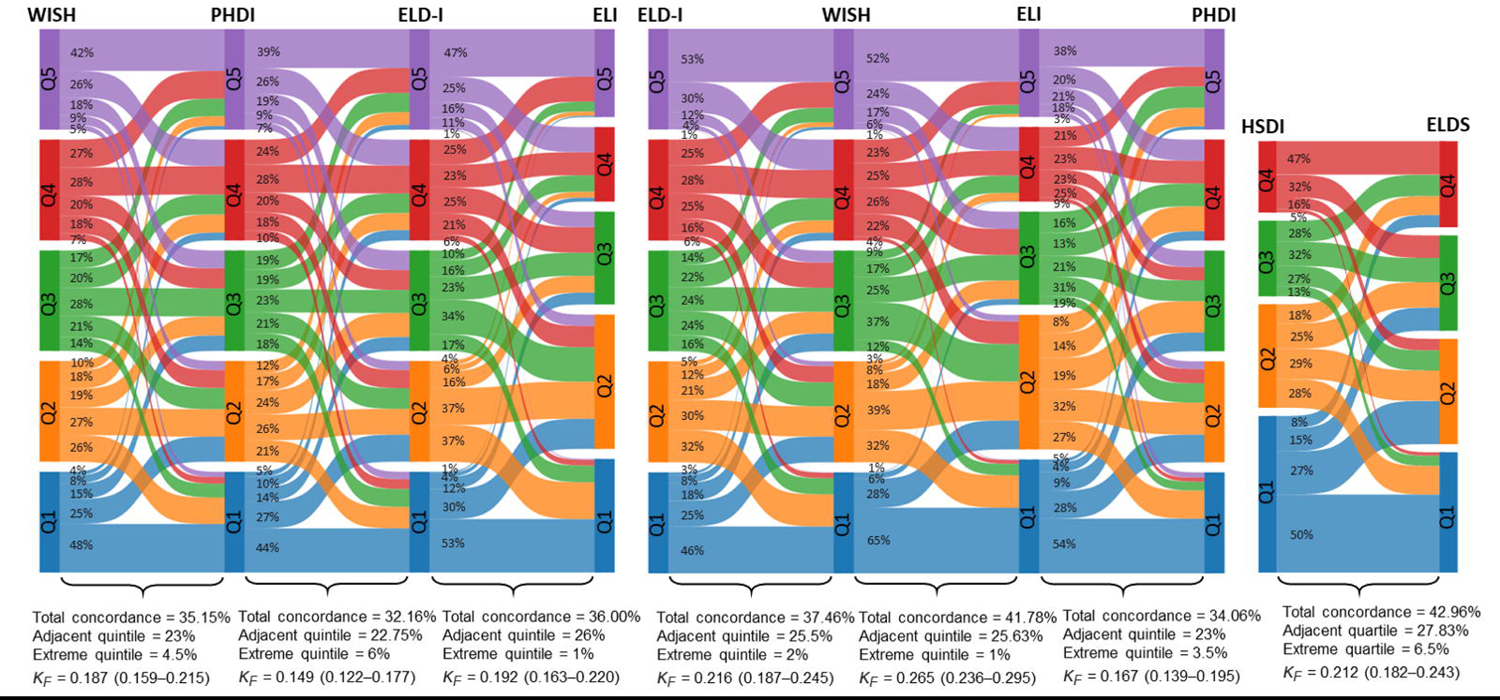
Inter-index concordance among EAT-Lancet-Based Indices quintiles/quartiles in the French Third Individual and National Study on Food Consumption Survey (INCA3, n = 1,723). Proportion of participants classified in the same quintile/quartile (total concordance), the adjacent quintile/quartile, and the opposite extreme quintile/quartile. KF = Fleiss’s kappa.

#### 3.4.1. Concurrent-criterion validity

According to ANCOVA models (Table 3), women had significantly higher means in WISH, ELI, PHDI, and ELDS scores than men. In addition, older age groups scored higher on all indices. On the other hand, the means in PHDI, ELD-I and ELI were significantly different according to educational level, with those with higher level having a higher mean score than those with lower formal education. Likewise, those individuals with higher income had higher scores in all indices, with the exception of HSDI. Regarding weight status, individuals in the lower BMI groups had higher scores, however, the trend was only confirmed for ELD-I and ELI. Conversely, non-smokers have higher means of WISH, ELD-I, and ELI. The level of physical activity was only related to the WISH and ELD-I indices. Values for means and standard deviations are available in Table S18 (Supplementary Material).

**Table 3.** Association between each EAT-Lancet indices and sociodemographic characteristics in adults form the French Third Individual and National Study on Food Consumption Survey (INCA3).

|  | **WISH** | **PHDI** | **ELD-I** | **ELI** | **HSDI** | **ELDS** |
| --- | --- | --- | --- | --- | --- | --- |
| **Sex** |  |  |  |  |  |  |
| Women | 41.49 (15.50) | 37.16 (11.39) | -1.37 (35.32) | 18.72 (3.94) | 3.94 (1.65) | 8.34 (1.46) |
| Men | 39.34 (15.45) | 36.47 (12.18) | -5.01 (31.94) | 17.48 (4.05) | 3.92 (1.59) | 7.85 (1.47) |
| *F* | 7.88 | 4.45 | 0.69 | 25.09 | 0.67 | 45.04 |
| *p-value* | 0.0051 | 0.0351 | 0.4059 | <0.0001 | 0.4129 | <0.0001 |
| *p-for-trend* | n/a | n/a | n/a | n/a | n/a | n/a |
| **Age** |  |  |  |  |  |  |
| 18-44 years-old | 35.61 (14.82) | 33.99 (11.62) | -9.75 (31.56) | 16.95 (4.00) | 3.61 (1.58) | 7.78 (1.46) |
| 45-64 years-old | 42.49 (14.85) | 38.64 (11.32) | -1.59 (35.26) | 18.82 (3.83) | 4.12 (1.64) | 8.22 (1.46) |
| ≥ 65 years-old | 48.93 (14.01) | 40.54 (11.46) | 10.99 (31.26) | 19.72 (3.68) | 4.39 (1.50) | 8.69 (1.38) |
| *F* | 49.36 | 30.65 | 26.70 | 49.26 | 13.8 | 25.44 |
| *p-value* | <0.0001 | <0.0001 | <0.0001 | <0.0001 | <0.0001 | <0.0001 |
| *p-for-trend* | <0.0001 | <0.0001 | <0.0001 | <0.0001 | <0.0001 | <0.0001 |
| **Education** |  |  |  |  |  |  |
| Primary and middle school | 41.89 (14.86) | 36.91 (11.74) | -3.75 (34.59) | 18.17 (3.84) | 4.08 (1.64) | 8.12 (1.40) |
| High school | 37.13 (16.44) | 33.94 (11.48) | -11.04 (34.68) | 17.10 (3.97) | 3.69 (1.66) | 7.83 (1.61) |
| 1 to 3 years of post-secondary education | 40.64 (16.65) | 38.48 (12.33) | 1.92 (32.24) | 18.66 (4.49) | 3.94 (1.55) | 8.11 (1.60) |
| ≥ 4 years of post-secondary education | 39.65 (14.55) | 37.75 (11.22) | 0.93 (30.41) | 18.37 (3.99) | 3.78 (1.54) | 8.29 (1.42) |
| *F* | 1.37 | 7.01 | 4.24 | 5.71 | 0.82 | 2.34 |
| *p-value* | 0.2501 | 0.0001 | 0.0054 | 0.0007 | 0.4815 | 0.0714 |
| *p-for-trend* | 0.5636 | 0.0003 | 0.0042 | 0.0076 | 0.5076 | 0.0008 |
| **Monthly income <sup>a</sup>** |  |  |  |  |  |  |
| <900 €/month/CU | 37.47 (14.28) | 34.89 (12.54) | -13.23 (32.10) | 17.28 (3.85) | 3.94 (1.68) | 7.71 (1.53) |
| 900-1,340 €/month/CU | 40.07 (15.82) | 35.33 (11.69) | -2.85 (37.25) | 17.99 (4.00) | 3.81 (1.67) | 8.12 (1.32) |
| 1,340-1,850 €/month/CU | 41.89 (16.05) | 37.83 (12.52) | 0.99 (33.54) | 18.65 (4.07) | 3.99 (1.56) | 8.20 (1.52) |
| ≥ 1,850 €/month/CU | 42.02 (15.51) | 38.46 (10.54) | 0.88 (30.80) | 18.56 (4.02) | 4.01 (1.59) | 8.33 (1.49) |
| *F* | 3.2 | 3.52 | 6.83 | 2.94 | 1.30 | 6.86 |
| *p-value* | 0.0225 | 0.0146 | 0.0001 | 0.0322 | 0.2729 | 0.0001 |
| *p-for-trend* | <0.0001 | <0.0001 | 0.0002 | <0.0001 | 0.0748 | <0.0001 |
| **Weight status** |  |  |  |  |  |  |
| Underweight | 33.96 (17.36) | 32.15 (14.44) | -17.92 (40.94) | 16.22 (5.39) | 3.29 (1.52) | 7.41 (1.64) |
| Normal | 39.42 (15.72) | 36.07 (12.04) | -0.83 (31.88) | 18.30 (4.22) | 3.86 (1.62) | 8.12 (1.53) |
| Overweight | 42.68 (15.11) | 38.47 (11.17) | -3.24 (33.47) | 18.26 (3.63) | 4.10 (1.58) | 8.19 (1.41) |
| Obesity | 40.17 (13.90) | 37.19 (11.41) | -9.43 (38.20) | 17.58 (3.87) | 3.90 (1.76) | 8.01 (1.38) |
| Morbid obesity | 38.51 (15.97) | 37.53 (10.55) | -1.31 (34.31) | 17.37 (3.70) | 4.02 (1.57) | 7.85 (1.65) |
| *F* | 4.77 | 2.78 | 5.95 | 7.92 | 3.30 | 4.20 |
| *p-value* | 0.0008 | 0.0256 | 0.0001 | <0.0001 | 0.0105 | 0.0022 |
| *p-for-trend* | 0.7019 | 0.7524 | 0.0853 | 0.0180 | 0.1508 | 0.1261 |
| **Smoking status** |  |  |  |  |  |  |
| No | 41.66 (15.52) | 37.55 (11.79) | -0.25 (33.07) | 18.43 (4.00) | 3.96 (1.58) | 8.20 (1.47) |
| Yes | 36.68 (14.85) | 35.17 (11.49) | -12.24 (34.27) | 17.12 (4.05) | 3.87 (1.75) | 7.80 (1.47) |
| *F* | 7.82 | 2.20 | 14.01 | 9.59 | 0.27 | 3.65 |
| *p-value* | 0.0052 | 0.1387 | 0.0002 | 0.0020 | 0.6047 | 0.0561 |
| *p-for-trend* | n/a | n/a | n/a | n/a | n/a | n/a |
| **Physical activity** |  |  |  |  |  |  |
| Low | 39.17 (15.24) | 35.84 (11.17) | -5.14 (32.67) | 17.90 (4.06) | 3.79 (1.53) | 8.01 (1.53) |
| Moderate | 41.38 (15.78) | 37.36 (12.44) | -2.57 (34.41) | 18.31 (4.03) | 4.02 (1.69) | 8.14 (1.45) |
| High | 39.01 (15.71) | 35.75 (11.70) | -1.59 (33.16) | 17.61 (4.19) | 3.82 (1.65) | 7.93 (1.47) |
| *F* | 3.01 | 2.74 | 0.51 | 2.59 | 2.71 | 2.95 |
| *p-value* | 0.0498 | 0.0328 | 0.5985 | 0.0750 | 0.0671 | 0.0524 |
| *p-for-trend* | 0.0388 | 0.1345 | 0.0011 | 0.0692 | 0.1958 | 0.8481 |

Note. a = Income per consumption unit. p-value = P referred to ANCOVA; p-for-trend = P referred to Jonckheere–Terpstra test for trend ordered predictors. n/a = not applicable.

#### 3.4.2. Convergent validity: Correlation with nutritional measures

The average PANDiet score was 64.83 (SD = 5.44). Correlations were the lowest (ρ < 0.20) for the HSDI and ELDS binary indices and ranged from 0.22 to 0.34 among other indices. Moreover, significant positive correlations were found between the adequacy sub-score (Mean = 63.42; SD = 12.10) and the WISH, PHDI, ELD-I, and ELI indices, with ρ ranging from 0.07 to 0.17. Regarding the moderation sub-score (Mean = 66.23; SD = 10.04), all the indices were positively related (ρ between 0.06 and 0.21) with PHDI and ELD-I having the lowest correlation values.

When analysing the adequacy at nutrient level, the results behaved differently according to the scoring system used. In general, the HSDI and ELDS indices (both based on a binary scoring) correlated inversely with several nutrients: protein, DHA, EPA+DHA, riboflavin, niacin, pantothenic acid, vitamin B-6, vitamin B-12, vitamin D, iodine, phosphorus, zinc, calcium, and iron. Conversely, the WISH, PHDI and ELD-I indices, which use a quantitative scoring, were positively correlated with the adequacy of most nutrients, including: polyunsaturated fatty acids, vitamins (e.g., A, thiamine, B-6, and E), minerals (e.g., manganese, magnesium, copper, and selenium). However, certain negative associations were found between these three indices. In this sense, the ELD-I correlated inversely with linoleic acid, vitamin B12, phosphorus, carbohydrates, and sodium, while the WISH and PHDI correlated inversely with niacin and total fat, respectively. As for the ELI, this index based on semi-quantitative scores shared traits with both quantitative and binary ones. While it showed positive associations with several indicators such as polyunsaturated fatty acids and vitamins D and E, it was negatively related to others such as protein, B-complex vitamins, phosphorus, calcium, and iron. The likelihoods of adequacy of fibre, thiamine, folate, vitamin C, and manganese were positively related to all six indices. Likewise, the likelihood of zinc adequacy correlated inversely with all of them. More details of the correlations are shown in Fig 4.

**Fig 4.**
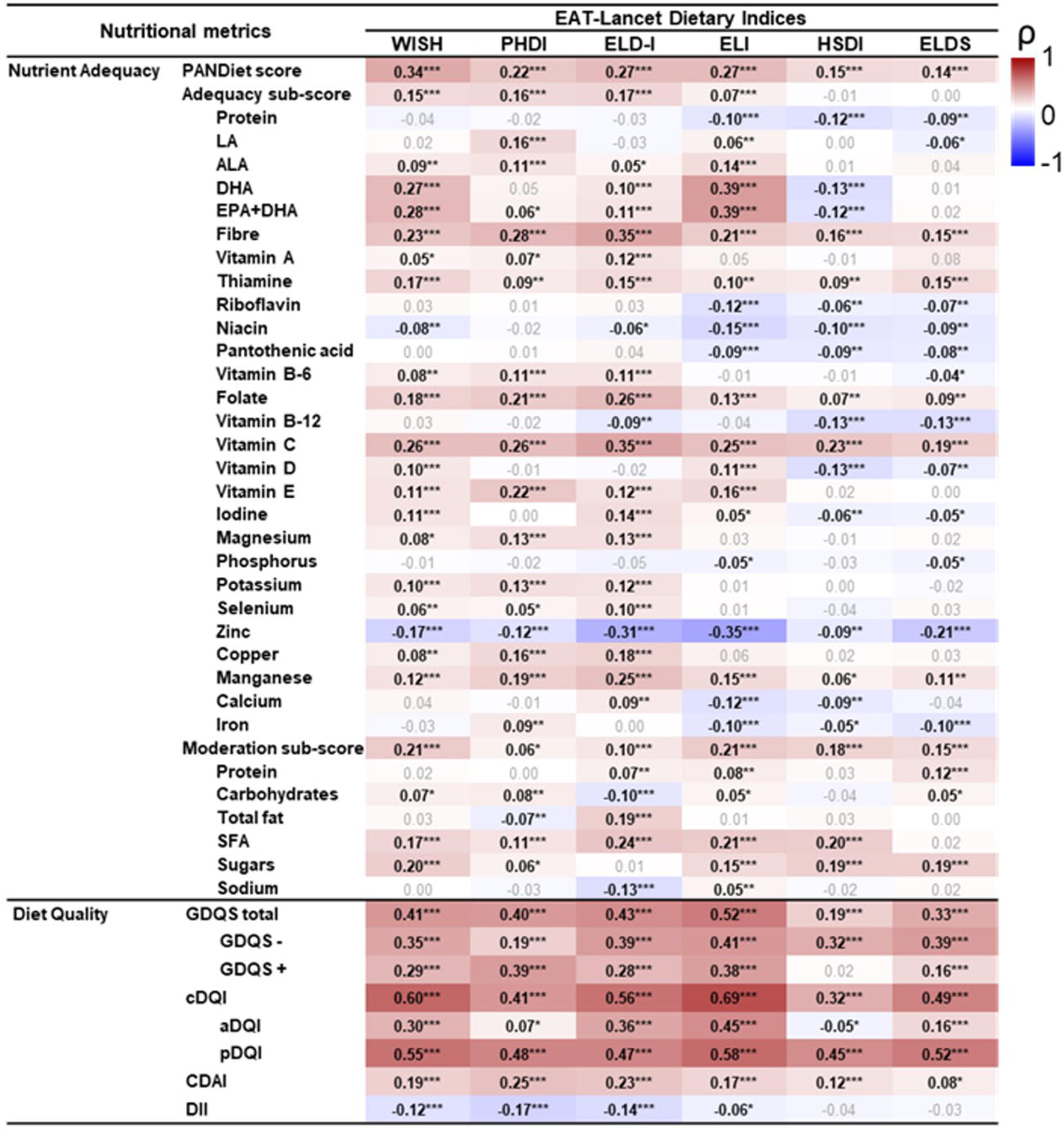
Correlations between the nutritional variables and the EAT-Lancet-Based Indices in the French Third Individual and National Study on Food Consumption Survey (INCA3, n = 1,723). Heat map plotting Spearman’s correlation coefficients (ρ): red indicates positive correlations, white indicates no correlations, and blue indicates negative correlations. PANDiet = Probability of Nutritional Adequacy. *p < 0.05, **p < 0.001, ***p < 0.0001.

Furthermore, the indices showed significant correlations with other nutritional quality scores. The six indices were positively associated with the GDQS (correlation coefficients between 0.19 and 0.52) and cDQI total scores (correlation coefficients between 0.32 and 0.69). In addition, the WISH, PHDI, and ELD-I indices were related to an antioxidant and anti-inflammatory diet. As such, these indices showed positive and weak correlations with CDAI (correlation coefficients between 0.19 and 0.25) and negative very weak correlations with DII (correlation coefficients between −0.12 and −0.17). Conversely, negligible or null correlations were found between the indices based on binary scoring and DII and CDAI.

#### 3.4.3. Convergent validity: Correlation with environmental impact indicators

Fig 5 shows a heat map for the correlation analysis between the indices and the aggregated indicator PEF and 14 individual metrics. Overall, the indices were negatively correlated with the indicators (highest ρ = −0.33), with the exception of water use and photochemical ozone formation, this latter only for WISH and ELI. However, it is important to mention that the highest correlations were found for ELD-I and ELI, while PHDI was the index that showed the weakest (ρ<-0.10) and least significant coefficients.

**Fig 5.**
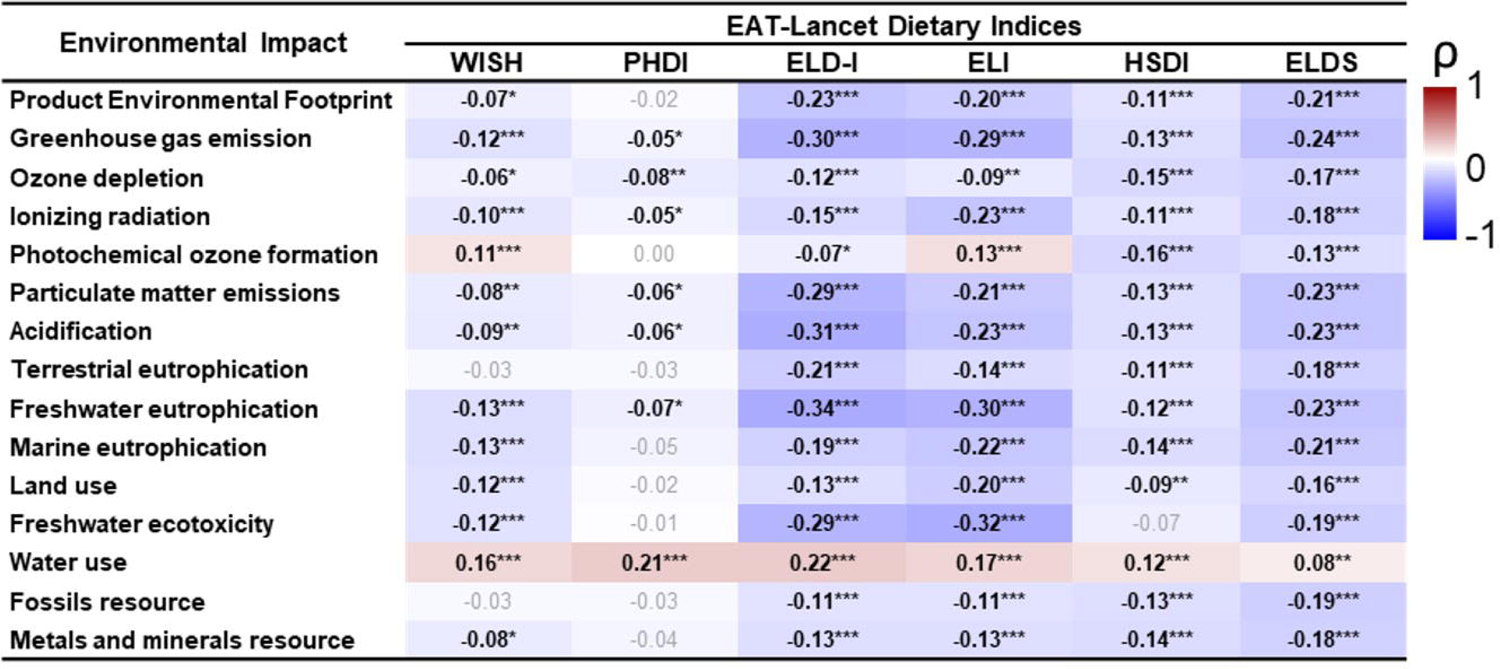
Correlations between the Environmental Impact and the EAT-Lancet-Based Indices in the French Third Individual and National Study on Food Consumption Survey (INCA3, n = 1,723). Heat map plotting Spearman correlation coefficients (ρ): red indicates positive correlations, white indicates no correlations, and blue indicates negative correlations. The Product Environmental Footprint is an aggregated indicator of the 14 environmental metrics. *p < 0.05, **p < 0.001, ***p < 0.0001.

### 3.5. Trends across level of adherence

The probability of nutritional adequacy was compared across quintiles/quartiles to identify trends. Regarding the PANDiet score, all indices exhibited significant differences between quantiles, with the indices using a quantitative scoring system showing the best performances (Fig. 6). However, the WISH and ELI indices revealed moderate effects on the PANDiet index (η^2^ = 0.121 and 0.083, respectively), whereas the effects of PHDI and ELD-I were small in magnitude (η^2^ = 0.059 and 0.053, respectively). Differences were also found when analyzing particular trends at the level of the PANDiet score components (Supplementary Material, Tables S18-S23). In this regard, the WISH index had its main effects on vitamin C, the moderation subscore and sugars, while the PHDI index had the least significant changes, with its main effect on fiber and vitamin C. As for ELD-I index, positive trends with moderate to large effect sizes were found for vitamin C, fiber, folate, and manganese. Additionally, significant differences were found, albeit of small magnitude, for polyunsaturated fatty acids (ALA, DHA, and EPA+DHA), vitamins (A, riboflavin, niacin, pantothenic acid, B-6, D, and E), and minerals (iodine, magnesium, potassium, selenium, copper, and calcium). While protein, LA, vitamin B-12, zinc, iron, carbohydrates, and sodium components showed a negative trend across the ELD-I quintiles, all were of small magnitude, except for zinc, which had a moderate effect size, or even showed no significant trends, as was the case with iron and proteins. On the other hand, noteworthy are the moderate-sized positive trends of the ELI index with respect to DHA, EPA+DHA, vitamin C, and negative trends with respect to zinc. Regarding the HSDI and ELDS indices, significant differences and trends were infrequent and of small magnitude.

**Fig 6.**
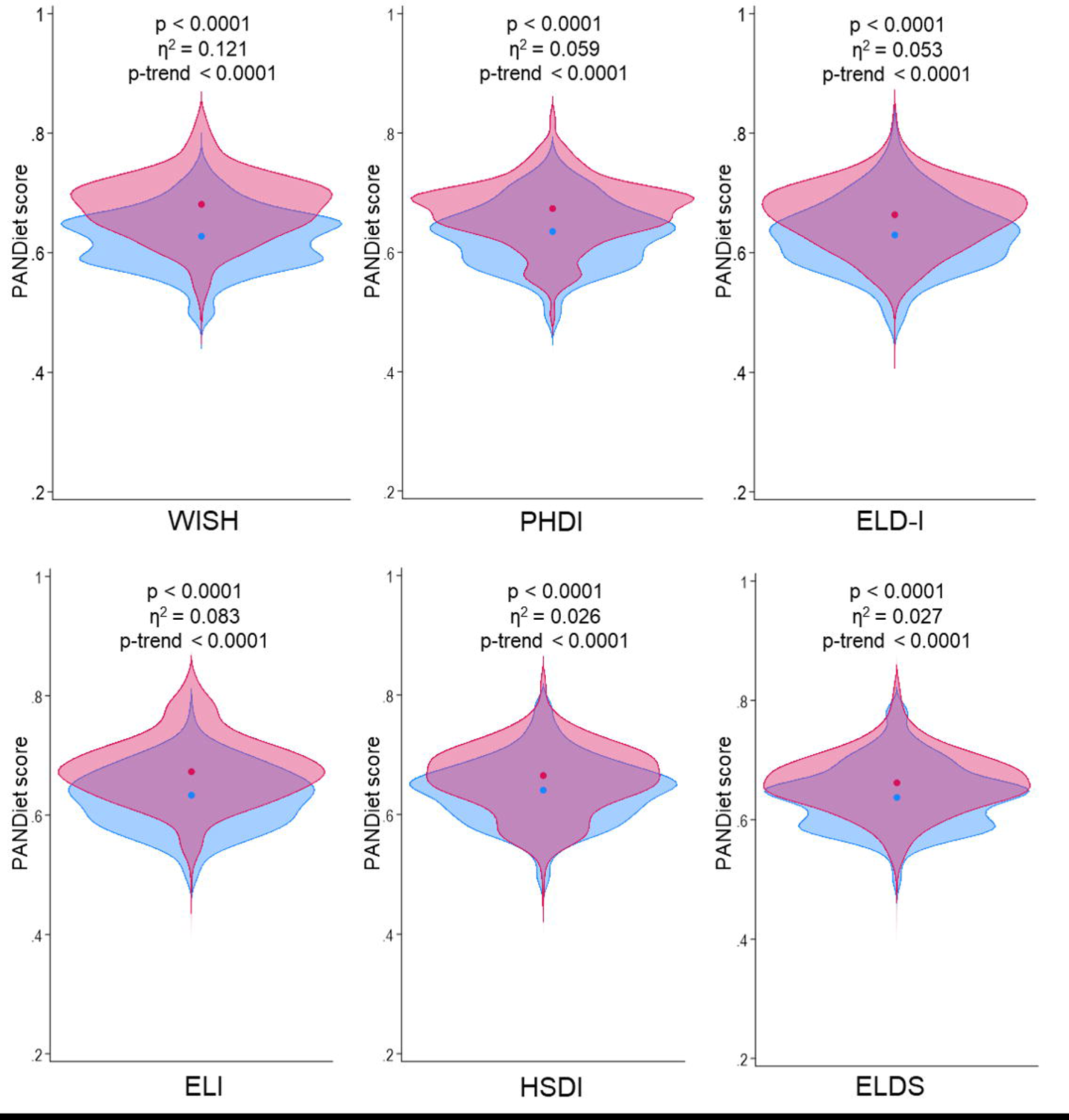
Violin plots comparing the distribution of PANDiet scores between the highest (in red) and lowest quintile/quartile (in blue) of EAT-Lancet-Based Indices in the French Third Individual and National Study on Food Consumption Survey (INCA3, n = 1,723). P-values and effect sizes obtained by ANOVA comparisons. Jonckheere–Terpstra test for trend was used. The circles denote the mean values.

As displayed in Fig 7, the greatest difference in PEF was found between the quintiles of ELD-I (p < 0.0001), with a moderate effect size (η² = 0.081) and an inverse trend (p < 0.0001). Significant results were also found in the other indices, except for the PHDI index. Regarding the specific environmental metrics (Supplementary Material, Tables S24-S29), the ELD-I index also showed the strongest differences, displaying negative trends with a strong effect on freshwater eutrophication and moderate effects on GHGE, particulate matter emissions, acidification, and freshwater ecotoxicity. Furthermore, it demonstrated a negative relationship with the other environmental indicators, excepting water usage that was positive. Among the other indices, ELI stood out, exhibiting negative trends in GHGE, freshwater eutrophication, and freshwater ecotoxicity, with small effects on the other indicators. Additionally, while significant differences were found across quartiles of WISH (all of small magnitude), trends were ruled out for Terrestrial eutrophication and Fossil resource. PHDI was the index least correlated with environmental indicators, as trends were ruled out for photochemical ozone formation, terrestrial eutrophication, freshwater ecotoxicity, and fossil resource. On the side of binary scoring indices, ELDS was associated with negative effects on a greater number of indicators than HSDI. Similar to ELD-I, ELDS showed negative trends with moderate effects for GHGE, particulate matter emissions, acidification, and freshwater eutrophication.

**Fig 7.**
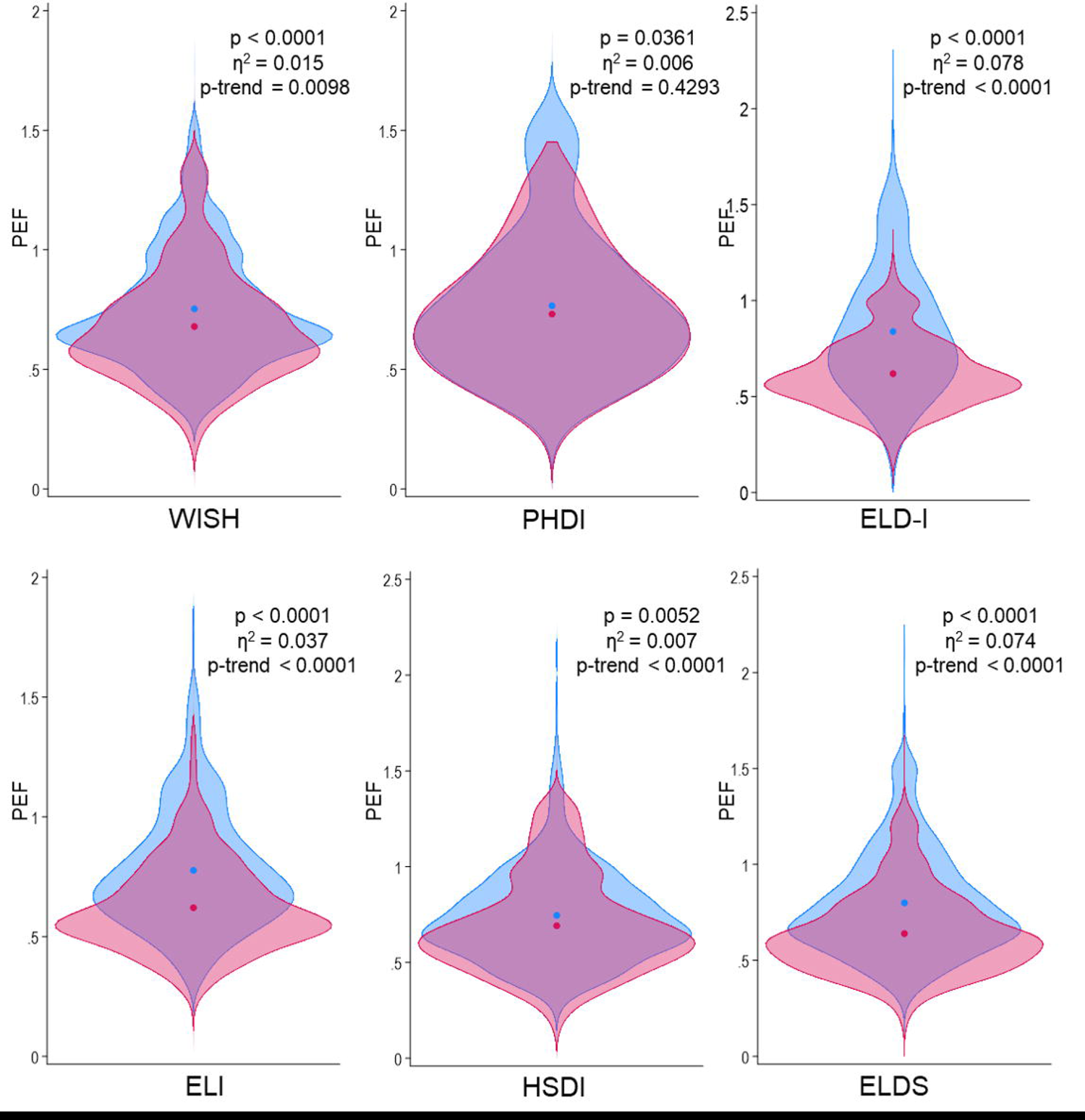
Violin plots comparing the distribution of Product Environmental Footprint (PEF) between the highest (in red) and lowest quintile/quartile (in blue) of EAT-Lancet-Based Indices in the French Third Individual and National Study on Food Consumption Survey (INCA3, n = 1,723). P-values and effect sizes obtained by ANOVA comparisons. Jonckheere–Terpstra test for trend was used. The circles denote the mean values. Finally, PANDiet/PEF ratio correlated significantly with WISH (ρ = 0.17, p < 0.0001), PHDI (ρ = 0.08, p = 0.0006), ELD-I (ρ = 0.31, p < 0.0001), ELI (ρ = 0.28, p < 0.0001), HSDI (ρ = 0.16, p < 0.0001), ELDS (ρ = 0.26, p < 0.0001). Further, positive and significant trend was observed in the relationship between the PANDiet/PEF ratio across quintiles/quartiles, with moderate size effects for ELD-I (η² = 0.088, p-for-trend <0.0001), ELDS (η² = 0.089, p-for-trend < 0.0001) and ELI (η² = 0.076, p-for-trend < 0.0001), and small effects for WISH (η² = 0.040, p-for-trend < 0.0001) and HSDI (η² = 0.026, p-for-trend < 0.0001). However, no significant differences were found in this variable for the PHDI index. More details are available in Supplementary Material.

## 4. Discussion

This study is the first to comprehensively evaluate the validity and reliability of six dietary indices representing the EAT-Lancet reference diet, using a national representative sample. While some of these indices have been partially validated in prior research, this study offers a comprehensive analysis of all indices on the same sample. Briefly, our findings indicate that the most reliable indices are those using quantitative scoring, especially those adjusted for energy intake (e.g., ELD-I), which robustly captured dietary variability, independently of energy intake. While the indices proved to be unidimensional and concurrently valid in differentiating scores based on sociodemographic factors, there was discordance in the classifications of individuals. In addition, the six indices demonstrated varied associations with nutrition and environmental impact, with significant weak correlations to nutritional adequacy and environmental impact but stronger correlations to diet quality. Notably, findings highlight that indices based on quantitative scoring were mainly associated with nutrition, while indices with binary scoring were more linked to environmental impact. Nonetheless, ELD-I was associated with both nutritional and environmental domains; however, WISH outperformed ELD-I concerning essential fatty acids. Furthermore, while all indices were associated with lower adequacy of certain nutrients, such as zinc, and higher water use, the magnitudes of these associations were relatively modest.

Reliability was evaluated by focusing on internal consistency and relationships between food components scores [19]. ELD-I and WISH exhibited the best internal consistency, while ELDS showed lower λ coefficients. Although there is no fixed rule for determining when λ is high enough, the context and researcher’s judgment are crucial, especially in nutrition, where lower coefficients are common due to the complexity of human diet [19,20,41]. Internal consistency is not strictly necessary, but knowing this property has implications for confidence in the indices [19].

Similar to previous reports, the impact of individual components on the total score varied significantly [12,17]. In this sense, fruits and vegetables demonstrated robust correlations, underscoring their importance in evaluating both health and sustainability. Conversely, whole grains and legumes exhibited weaker correlations, which could be due to the challenges associated with meeting targets for less frequently consumed foods. Although all food components contribute, the indices currently assign them equal weights despite variations in their impact on health and the environment [42,43], suggesting a potential improvement by assigning different weights to better reflect their relative impact [44,45]. Also, indices using quantitative scoring captured more the interindividual variability, especially ELD-I, increasing the validity in assessing and comparing diets [46]. This was expected, as the ability to capture data variability depends on the type of measurement used. Unbounded continuous measures such as ELD-I allow detailed representation by covering a full range of values within an infinite spectrum. On the other hand, bounded continuous measures such as WISH and PHDI restrict variability to specific values within a finite set. In addition, measures based on binary indices (i.e., HSDI and ELDS), by classifying food compounds into only two categories, further limit the representation of variability. Consequently, the selection of metric type can markedly shape the comprehensiveness and precision of the analysis. Further, ELD-I and PHDI were not affected by the amount of energy consumed, unlike other indices that showed moderate inverse correlations with energy intake. This energy adjustment in ELD-I and PHDI [13,16] avoids biases associated with unbalanced calorie diets, where high energy intake may result in high scores [47].

Despite the theoretical expectation of significant correlations between indices assessing adherence to EAT-Lancet recommendations, our study found low concordance among indices. The limited concordance may be attributed to differences in index design, including components, thresholds, weighting, and scoring systems. These findings align with previous studies comparing indices for Mediterranean diet adherence [48,49], emphasizing the importance of considering methodological differences in interpreting similar results.

In terms of structural validity, the unidimensionality of all indices was confirmed, with optimal fit, indicating plausible representations of the underlying relationships between food components [50]. Moreover, ELD-I, PHDI and WISH explained the variability of the food components at 57%, 47% and 32% supporting their robust structural validity. Unidimensionality, regardless of whether the concept encompasses multiple domains or facets, is a key requirement for instruments that rely on a “total score”, such as the EAT-Lancet indices. The unidimensionality found in the current study suggests that the food components within each index are associated with a single concept of a healthy and sustainable diet, thus supporting the use of a total score to simplify its comprehension and applicability [51,52]. However, it is recommended that the use of total scores be complemented by a detailed analysis of the food components. Furthermore, differences according to sociodemographics were found, supporting their concurrent criterion validity. Although some indices did not reach statistical significance, the general pattern indicated that scores were higher in women, older individuals, with higher income, higher education, lower BMI, no-smokers, and physically active. These differences among demographic groups are consistent with previous studies on EAT-Lancet recommendations [53–57].

Regarding convergent validity, the indices presented variations in their correlation with nutritional adequacy. The quantitative scoring indices, especially ELD-I and WISH, showed a positive, albeit weak, correlation with most PANDiet metrics (including, total PANDiet score, sub-scores and nutrient adequacies). This is similar to a previous study showing that, despite a reduction in animal food consumption, the highest ELD-I quintiles had an increase in PANDiet score [16]. In contrast, indices with binary or semi-quantitative scoring were negatively associated with nutritional adequacy for several nutrients, supporting the need to establish minimum intake values to improve the accuracy of nutritional index measurements [58,59]. Regardless of the scoring system, an inverse relationship was observed between the EAT-Lancet indices and the nutritional adequacy of zinc and vitamin B-12, corroborating previous findings [58,60]. Quantitative indices demonstrated greater validity by correlating closely and more strongly with dietary quality indicators, such as the GDQS and cDQI (ρ < 0.69). These associations were expected, given that the indices promote the intake of healthy animal and plant foods, sources of antioxidant and anti-inflammatory compounds [61–63]. Overall, the results of these analyses support the association of the indices with a healthy diet, promoting nutritional adequacy and the consumption of antioxidant and anti-inflammatory compounds, with potential health benefits.

As for convergent validity related to environmental impact, stronger correlations were found for the ELD-I and ELI. Several studies support the positive impact of EAT-Lancet recommendations on the environment, such as significant reductions in GHGE and land use [15,64]. In addition, the food components that contribute most to the indices (e.g., fruits and vegetables) are consistent with results on the effect of their increased intake on environmental aspects [65,66]. However, it is important to note that while these dietary patterns may have environmental benefits, the trade-off, such as increased water use [67], must be considered at the national and/or subnational level, given the water stress in a large number of countries [26,68].

Discrepancies in mean values of dietary index compared to the original studies suggest different consumption patterns according to population and geographic location. For instance, the WISH index was lower in our study compared to the original [12], potentially reflecting variations in dietary habits between France and Vietnam. Furthermore, while the original study reported perfect scores for added sugars and saturated oils, in our context, fewer participants indicated not consuming these food components [12]. While Vietnam has undergone a nutritional transition in recent decades, characterized by an increase in sugar and fat consumption, there is still evidence of lower consumption of these foods compared to France (46.5 g/day vs. 92.84 g/day and 8 g/day vs. 16 g/day, respectively) [69–71]. Moreover, the high compliance with the fruit and vegetable recommendations in the Brazilian validation study results in almost perfect scores for these groups [13], possibly because that study does not fully reflect the diet of the country. In a more recent evaluation, the authors applied the PHDI to a nationally representative survey from Brazil, finding that the mean scores in both groups are lower than those obtained in our study. This was expected, considering that fruit and vegetable consumption is higher in France than in Brazil (378 vs. 150 g/d, respectively) [72,73].

Regarding the ELD-I index, the pattern was similar to those of the study that developed the index, which was expected as samples are from the same geographical context [16]. In line with previous evidence, we observed a lack of variation in the unsaturated oils component, suggesting that the threshold based on the EAT-Lancet recommendations may not be appropriate for consumption levels in France [60]. These results indicate that the cut-off point established on the basis of the EAT-Lancet report exceeds the average consumption level observed in France (around 8 g/d) according to the ELD-I criteria (≤ 80 g/d) [71].

Regarding ELI index, we found a mean score similar to that of the Swedish study [16], although the food components with higher scores differed, possibly due to differences in consumption patterns between Sweden and France [74,75]. In this sense, the Swedish cohort used for the design of the ELI index reported a lower consumption of vegetables (< 200 g/d), and a higher consumption of potatoes (> 100 g/d) and fish (> 50 g/d) when compared to the INCA3 [71,72] These geographical divergences also were found for binary indices. For example, the average score in the HSDI was twice as high in the original study conducted in Mexico [11]. This is because a significant proportion of the participants met the recommendations for several food components, such as tubers, unsaturated fats, fish, saturated fats, and beef, which aligns with Mexican dietary patterns [76]. Similarly, in the case of the ELDS index, differences were observed in the high compliance groups in the original UK sample compared to our sample in France [10]. It should be noted that in the original studies there was no variability in the scores of the unsaturated fats in the HSDI and the dry beans, lentils and peas in the ELDS, which was not replicated in our sample.

Overall, the differences in scores between the studies reflect variations in dietary consumption patterns in different regions and populations, emphasizing the importance of considering the context and specific characteristics when interpreting the dietary indices [77]. However, these findings confirm the sensitivity of the indices in capturing dietary cultural variability. Likewise, discrepancies in dietary intake could stem from variances in the intrinsic features of the study designs, such as the method employed for nutritional assessment (i.e., 24-hour dietary recall, food frequency questionnaires, or food diaries) and the number of days covered (i.e., single or repeated measure).

These findings should be interpreted within the context of some limitations. First, the cross-sectional design hinders the estimation of predictive validity and the assessment of the association with health outcomes beyond anthropometry was not possible due the absence of these data in the INCA3 survey. Future studies are encouraged to analyze the link with non-communicable diseases to strengthen validity research. Second, the manual disaggregation of complex dishes may introduce errors, so a continuous effort in the construction of composition tables and standardized recipes is necessary to accurately estimate the population intake in France. Nevertheless, it is crucial to acknowledge inherent limitations in the EAT-Lancet recommendations that may result in methodological inconsistencies. In this sense, the imprecision and lack of clarity regarding specific food groups within the EAT-Lancet diet can present challenges in its operationalization, especially in estimating the quantities of fats and added sugars, fostering uncertainty and personal interpretation [78,79]. Also, it would be useful to clearly define and standardize the quantification method for certain food components, such as whole grains or legumes, specifying whether intake should be reported in grams of cooked or dry weight. This would improve comparisons and prevent discrepancies arising from user interpretation. In this study, results are presented using grams of food intake; however, we confirmed that findings remain consistent when utilizing grams of dry weight, as expected due to the low intake of these food groups (unpublished data). Third, acknowledging limitations in the Agribalyse v.3.1.1 database is crucial, including the absence of soil carbon measurement in GHGE, information on biodiversity, phytosanitary product impact, and waste [80]. Additionally, incomplete water use inventory data highlights the need for considering spatial and temporal variability [81]. This demonstrates the continuing need for more comprehensive databases, incorporating various estimates related to food production methods, for accurate assessments of dietary sustainability in future research.

As the results of the present study suggest, the measurement performance of indices assessing the adherence to the planetary health diet proposed by the EAT-Lancet commission may vary, potentially impacting the reliability and validity of these indices. Therefore, it is essential to establish clear criteria for the contribution of each food component in the indices, including the number of components, scoring criteria, the use of adequate cutoff points, energy adjustment, and component weighting, aiming to enhance coherence among existing indices. To achieve this, we recommend following the framework provided by Waijers et al. [43] regarding key considerations in constructing a dietary index: 1) it needs to have a clear objective, 2) a rationale for the choice of index components, 3) clear information on assigning foods to food groups, 4) include an exact quantification of the index components against cut-off values, 5) energy adjustment (or not), and 6) information on the relative contribution of individual components to the total score.

Furthermore, we also consider essential that, with the launch of the EAT-Lancet diet 2.0 in 2024, a consensus should be reached on how to measure adherence to its recommendations. This will help to avoid an “overdevelopment” of indices, similar to what has happened with the dietary indices assessing the adherence to the Mediterranean diet [48,82]. An excessive proliferation of indices may cause challenges in terms of consistency and comparability between studies, making it difficult to identify a common standard for assessing adherence to the EAT-Lancet diet and further complicate the interpretation of research results and the implementation of these recommendations.

## 5. Conclusion

The different approaches to assess adherence to a sustainable and healthy diet are complementary, and the superiority of one method over another cannot be asserted. Thus, it is crucial to carefully address methodological issues to better understand the utility and applicability of these indices, including the precise clarification of objectives and assumptions, as well as a detailed description of score composition. In this regard, while indices like the ELD-I tend to reflect the healthiness and sustainability of the diet, others may be more valid for examining one of these two domains. For example, WISH is particularly effective as an indicator of diet adequacy. The choice of an index will depend on the specific needs of researchers. In practical terms, quantitative scoring indices are valuable tools in studies where precision and granularity are important such as clinical trials or epidemiological studies. Despite the associated cost of reduced variability and loss of statistical power, binary scoring indices find utility in surveys, observational studies, and public health interventions. Therefore, understanding the advantages and disadvantages of each index is relevant for interpreting the results of such investigations.

Given the ongoing development of new indices for assessing adherence to the EAT-Lancet recommendations, it is essential to conduct comprehensive assessment of the measures in terms of reproducibility, validity, and comparisons between different methodologies. This becomes even more crucial with the forthcoming publication of version 2.0 of the EAT-Lancet report in 2024, which is expected to address the main concerns identified in recent years.

## Funding

This study is part of the FEAST (Food systems that support transitions to healthy and sustainable diets) project funded by the European Union’s Horizon Europe research and innovation program under grant agreement number 101060536 and by Innovate UK under grant number 10041509. Swiss participant in FEAST is supported by the Swiss State Secretariat for Education, Research and Innovation (SERI) under contract number 22.00156. More details in https://www.feast2030.eu/.

## Competing interests

The authors declare that they have no conflict of interest.

## Data Availability

Data from the Third French Individual and National Food Consumption Survey (INCA3) in available on the data.gouv.fr platform. Data on the environmental impacts of foods consumed in France is available on the agribalyse.ademe.fr platform.

https://www.data.gouv.fr/en/datasets/donnees-de-consommations-et-habitudes-alimentaires-de-letude-inca-3/

## Notes

### Competing Interest Statement

The authors have declared no competing interest.

### Author Declarations

The INCA3 study was carried out in accordance with the Declaration of Helsinki and received approval from the French Data Protection Authority (Decision DR 2013-228) on May 2, 2013, following a favorable opinion from the Advisory Committee on Information Processing in Health Research on January 30, 2013 (Opinion 13.055). Verbal informed consent was obtained from all participants before their voluntary inclusion in the study. Verbal consent was witnessed and formally recorded.

