## Supplementary material for "How do the indices based on the EAT-Lancet recommendations measure adherence to healthy and sustainable diets? A comparison of measurement performance in adults from a French national survey"

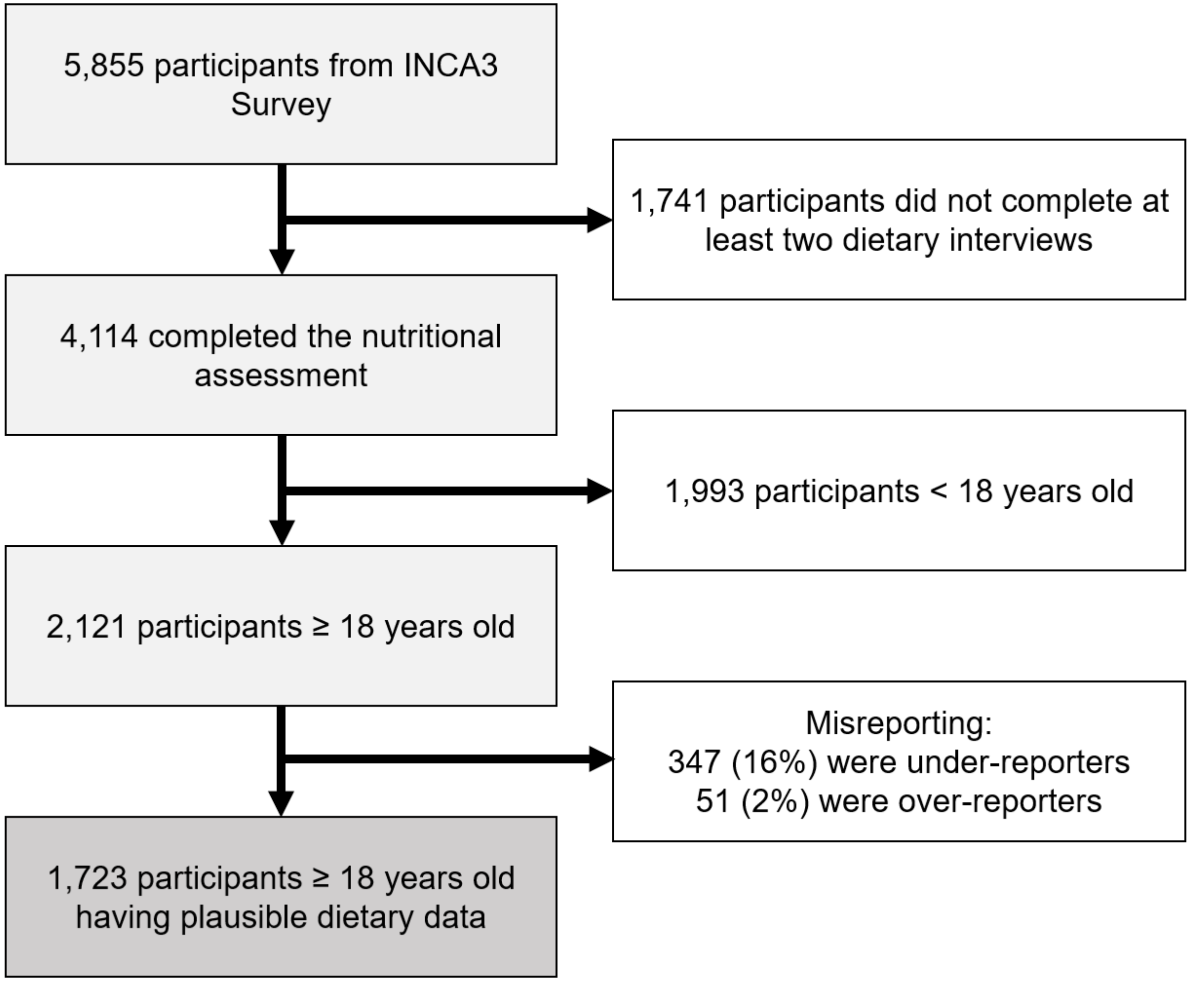

**Fig S1. Flowchart of sample selection process from the French Third Individual and National Study on Food Consumption Survey (INCA3).**

**World Index for Sustainability and Health**

The World Index for Sustainability and Health (WISH) was developed to assess the healthiness and environmental sustainability of the diet based on EAT-Lancet recommendations. It comprises 13 food components classified as neutral, protective or negative for human and planetary health. Each food component is assessed by a quantitative score, using reference values in grams, which limits the independent individual assessment of total energy intake. The WISH index excludes tubers and starchy vegetables on the argument that they are not in the WHO and Global Burden of Disease study. It was developed using data collected through repeated 24-hour recall from a sample of 396 adults in Vietnam (urban area). The WISH score has been linked to certain health outcomes, such as decreased depression levels and improved cognitive performance.

The following formula is applied to compute the score of an intake between the lower recommended intake and the recommended intake:

$$WISH = 10 * \frac{reported\ intake - lower\ recommended\ intake}{recommended\ intake - lower\ recommended\ intake}$$

On the other hand, the score between the recommended intake and the upper recommended intake is calculated according to the following formula:

$$WISH = 10 * \frac{(upper\ recommended\ intake - recommended\ intake) - (reported\ intake - recommended\ intake)}{upper\ recommended\ intake - recommended\ intake}$$

Saturated oils and added sugars are scored as a bivariate component: 10 points are allocated for those consumptions within the recommended range and zero points are given for those exceeding this recommendation. The total score is calculated by adding the score of each food component, and ranges between 0 up to 130 points. Moreover, the authors propose four sub-scores derived from the WISH index: healthy, less healthy, low environmental impact and high environmental impact.

The following table presents the food components of WISH, proving examples of food items and a more detailed explanation of the scoring system.

**Table S1. World Index for Sustainability and Health**

| Food components | Food items | Scoring |
|---|---|---|
| **Whole Grains** | Rice, wheat and corn (bran, germ, and endosperm in their natural proportion) or whole grain products in the form of breakfast cereals, bread, pasta, biscuits, muffins, tortillas, pancakes, and other sources. | ≥125 g/d = 10 points.<br><100 g/d = 0 point.<br>100–125 g/d = 0–10 points. |
| **Vegetables** | Fresh, frozen, cooked, canned, or dried vegetables, and excludes legumes and salted or pickled vegetables, juices, nuts, and seeds, and starchy vegetables such as potatoes or corn. | ≥300 g/d = 10 points.<br><200 g/d = 0 point.<br>200–300 g/d = 0–10 points. |
| **Fruits** | Fresh, frozen, cooked, canned, or dried fruits but excluding fruit juices, salted or pickled fruits and nuts or seeds. | ≥200 g/d = 10 points.<br><100 g/d = 0 point.<br>100–200 g/d = 0–10 points. |
| **Dairy Foods** | Whole or skimmed milk or derivative equivalents (e.g., cheese, yoghurt or curd), excluding butter and cream. | 250–500 g/d = 10 points.<br>0–250 g/d = 0–10 points.<br>>500 = 0 point. |
| **Red Meat** | Beef, pork, lamb, and goat including red meat in its processed form but excluding poultry, fish, eggs. | ≤14 g/d = 10 points.<br>>28 g/d = 0 point.<br>14–28 g/d = 0–10 points, inversely. |
| **Fish** | Fish and shellfish such as mussels and shrimp. | 28–100 g/d = 10 points.<br>0–28 g/d = 0–10 points.<br>>100 = 0 point. |
| **Eggs** | Eggs from chickens and ducks, but excludes fish eggs. | ≤13 g/d = 10 points.<br>>25 g/d = 0 point.<br>13–25 g/d = 0–10 points, inversely. |
| **Chicken and Other Poultry** | Meat from chicken, ducks, geese etc. | ≤29 g/d = 10 points.<br>>58 g/d = 0 point.<br>29–58 g/d = 0–10 points, inversely. |
| **Legumes** | Fresh, frozen, cooked, canned, or dried legumes, beans and lentils, but also includes soy foods and peas. | 0-75 g/d = 0–10 points.<br>≥75 g/d = 10 points. |
| **Nuts** | Tree nuts and ground nuts (including peanuts). | 50–75 g/d = 10 points.<br>0-50 g/d = 0–10 points.<br>≥75 g/d = 0 point. |
| **Unsaturated Oils** | Olive, soybean, rapeseed, sunflower and peanut oil. | ≥40 g/d = 10 points.<br><20 g/d = 0 point.<br>20–40 g/d = 0–10 points.<br>> 80 g/d = 0 point. |
| **Saturated Oils** | Dairy fats, lard or tallows, and palm oil. | ≤11.8 g/d= 10 points<br>> 11.8 g/d = 0 point. |
| **Added Sugars** | Added sugar and sugar sweetened beverages. | ≤31 g/d= 10 points<br>> 31 g/d = 0 point. |

Trijsburg L, Talsma EF, Crispim SP, Garrett J, Kennedy G, de Vries JHM, Brouwer ID. Method for the Development of WISH, a Globally Applicable Index for Healthy Diets from Sustainable Food Systems. Nutrients. 2020;13(1):93.

**Planetary Health Diet Index**

The Planetary Health Diet Index (PHDI) stands as one of the most widely recognized indices based on the reference dietary guidelines proposed by the EAT-Lancet Commission. Developed and validated using data from over 15,000 individuals of the ELSA-Brazil study, this index evaluates the caloric density of 16 food components through a continuous scoring system. The sample used for its development consisted in active and retired workers from six Brazilian public universities, aged 35 to 74, from three major regions collected between 2008 and 2010 that were assessed with FFQ. The PHDI covers all the food components proposed by EAT-Lancet, and the suggested ranges and midpoints for each component are calculated based on their energy contribution to a reference diet of 2500 kcal/day. Using a gradual scoring, the PHDI allows components to receive scores based on the amount of intake. These scores are determined as a caloric intake ratio. For any component of the PHDI, the caloric intake ratio is defined as the sum of calories from all foods ranked in that component divided by the total calories from all foods covered by the PHDI.

As for the scoring, each of the 16 components can award a maximum of 10 or 5 points, resulting in a total score ranging from 0 to 150 points. This approach provides the ability to assess diet quality by considering various aspects, offering a quantitative measurement that encompasses various nutritional components and their intake levels. As a special feature, PHDI includes interchangeability between some food components.

The authors of the PHDI have presented validity and reliability indicators. In terms of internal reliability, it has been considered adequate, and the results of construct validity indicate that the PHDI presents a positive correlation with nutritional indicators, energy independency, and is structurally valid. In addition, the GHGE calculation was carried out to confirm its validity. Additionally, it exhibits an inverse association with various health indicators, such as obesity, blood pressure, total cholesterol, LDL cholesterol, and non-HDL cholesterol, although no significant association with HDL cholesterol, triglycerides or insulin resistance has been found.

The following table presents the food components of the PHDI, proving examples of food items and a more detailed explanation of the scoring.

**Table S2. Planetary Health Diet Index**

| Food components | Foods | Scoring |
|---|---|---|
| **Nuts and peanuts** | Processed and raw nuts, pistachios, almonds, peanuts, and coconut pulp and milk | ≥11.6% = 10 points.<br>0% = 0 point.<br>0–11.6% = 0–10 points. |
| **Legumes** | Dry or canned beans, pulses, lentils, chickpeas, peas, soybeans, and soy food products. | ≥11.4% = 10 points.<br>0% = 0 point.<br>0–11.4 % = 0–10 points. |
| **Fruits** | Fresh and processed fruits, including culinary or industrial products such as fruit juices, nectars, and punches. | ≥5% = 10 points.<br>0% = 0 point.<br>0–5% = 0–10 points. |
| **Total vegetables** | Fresh, frozen, cooked, canned, or dried vegetables and excluded legumes and starchy vegetables. | ≥3.1% = 10 points.<br>0% = 0 point.<br>0–3.1% = 0–10 points. |
| **Whole grains** | Whole grains used as staple foods (e.g., brown rice, brown bread, oat flakes, canned grains, etc.), but not processed grains or refined products, such as polished rice and white bread. | ≥32.4% = 10 points.<br>0% = 0 point.<br>0–32.4% = 0–10 points. |
| **Eggs** | Eggs from chickens and other poultry. | 0–0.8% = 0–10 points.<br>0.8–1.5% = 0–10 point, inversely.<br>>1.5% = 0 point. |
| **Fish and seafood** | Fish and seafood. | 0–1.6% = 0–10 points.<br>1.6–3.1% = 0–10 point, inversely.<br>>3.1% = 0 point. |
| **Dairy** | Cow's milk, goat's milk, buffalo products, yogurt, and cheese, but no butter and sour cream. | 0–6.1% = 0–10 points.<br>6.1–12.2% = 0–10 point, inversely.<br>>12.2% = 0 point. |
| **Vegetable oils** | Vegetable oils. | 0–16.5% = 0–10 points.<br>16.5–30.7% = 0–10 point, inversely.<br>>30.7% = 0 point. |
| **Dark green vegetables to total vegetables ratio** | All dark green vegetables were included, such as broccoli and rocket, but not light green vegetables, such as lettuce. | 0–29.5% = 0–5 points.<br>≥29.5% = 0–5 point, inversely. |
| **Red and orange to total vegetables ratio** | Tomatoes, beets, carrots, and pumpkins are examples of foods included in this component. | 0–38.5% = 0–5 points.<br>≥38.5% = 0–5 point, inversely. |
| **Red meats** | Beef, lamb, and pork. Processed beef and pork (for example, sausages, ham, bologna, dried meat) were also accounted. | 0% = 10 points.<br>0–2.4% = 0–10 point, inversely.<br>>2.4% = 0 point. |
| **Chicken and substitutes** | Chickens, poultry, and their substitutes (i.e., eggs and fish and seafood). Processed poultry meat (e.g., smoked brisket and nuggets, smoked breast, and pate). | 0% = 10 points.<br>0–5% = 0–10 point, inversely.<br>>5% = 0 point. |
| **Animal fats** | Dairy fats (Butter and creams, including sour cream and cheese cream), lard, and tallow. | 0% = 10 points.<br>0–1.4% = 0–10 point, inversely.<br>>1.4% = 0 point. |
| **Added sugars** | All sweeteners, including white or brown sugars and honey used as ingredients in processed or culinary products and the added sugars to manufactured foods and beverages. | 0% = 10 points.<br>0–4.8% = 0–10 point, inversely.<br>>4.8% = 0 point. |

Cacau LT, De Carli E, de Carvalho AM, Lotufo PA, Moreno LA, Bensenor IM, Marchioni DM. Development and Validation of an Index Based on EAT-Lancet Recommendations: The Planetary Health Diet Index. Nutrients. 2021;13(5):1698.

### EAT-Lancet diet index

Data from 29,210 French adults (75% women) who participated in the NutriNet-Santé study were used to developed the EAT-Lancet Dietary Index (ELD-I). The ELD-I comprehensively covers 14 food components using cut-off points adapted from those proposed in the EPIC (European Prospective Investigation into Cancer and Nutrition)-Oxford study. The ELD-I is based on a quantitative scoring system that was designed to more accurately capture the variability in food consumption patterns, incorporating individual’s total energy intake and standardizing it to a daily intake of 2,500 kcal. The computation results in a continuous variable being either positive or negative. As the score increases, the individual's diet aligns more closely with the EAT-Lancet recommendations. Moreover, the validity of the ELD-I has been explored, with results confirming its ability to measure healthiness and sustainability of diets.

The following table presents the food components of the ELD-I, proving examples of food items and a more detailed explanation of the scoring.

**Table S3. EAT-Lancet diet index**

| Food components | Food items | Scoring computation |
|---|---|---|
| **Whole grains** | Rice, wheat, corn, and other. | $ELD-I_i = \frac{100\times\left\{\sum_{component\ i=1}^{14} \frac{a_i\times\left(cut-off_i-\frac{consumption_{ij}\times 2500}{Energy\ intake\ _j}\right)}{cut-off_i}\right\}}{14}$ Where *i* referred to on the 14 food groups and j is the individual. $\alpha_i = 1$ for component to limit and $\alpha_i = -1$ for component to promote. |
| **Potatoes and tuber** | Potatoes and cassava. | |
| **Vegetables** | All non-starchy vegetables. | |
| **Fruits** | All fruits. | |
| **Dairy foods** | Whole milk or derivative equivalents (e.g., cheese). | |
| **Beef, lamb, pork** | Red meats of beef, lamb and pork and processed meats. | |
| **Chicken and poultry** | Chicken and other poultry (e.g., turkey, duck, and quail). | |
| **Eggs** | Eggs from chicken, duck, and goose. | |
| **Fish** | Fish and shellfish (e.g., mussels and shrimps). | |
| **Legumes** | Dry beans, lentils, peas, soy foods. | |
| **Nuts** | Almonds, hazelnuts, pistachios and walnuts. | |
| **Saturated oil** | Dairy fats (e.g., cream and butter), lard or tallow. | |
| **Unsaturated oils** | Olive, soybean, rapeseed, sunflower, and peanut oil. | |
| **All sweet** | Added sugars and sugar-sweetened beverages. | |

Kesse-Guyot E, Rebouillat P, Brunin J, Langevin B, Allès B, Touvier M, et al. Environmental and nutritional analysis of the EAT-Lancet diet at the individual level: insights from the NutriNet-Santé study. J Clean Prod. 2021;296:126555.

### EAT-Lancet Index

The EAT-Lancet Index (ELI) was developed using data from 22,421 Swedish adults aged 45 to 73 who participated in the Malmö Diet and Cancer cohort between 1991 and 1996, which included food diaries and food frequency questionnaires. The ELI covers 14 food components categorized as emphasized and restricted foods, with daily gram quantities assessed through a semi-quantitative scoring system: from 0 to 3 points. This index has been related with reduced all-cause mortality, decreased mortality from cancer and cardiovascular diseases. Furthermore, it was observed that higher adherence to this index is associated with a lower risk of type 2 diabetes in Swedish adults, and this relationship is dose-response and independent of genetic susceptibility.

The following table presents the food components of the ELI, proving examples of food items and a more detailed explanation of the scoring.

**Table S4. EAT-Lancet Index**

| Food components | Food items | Scoring | | | |
|---|---|---|---|---|---|
| | | 3 points | 2 points | 1 point | 0 points |
| **Vegetables** | All vegetables except legumes. | >300 | 200–300 | 100–200 | <100 |
| **Fruits** | Fruits and berries. | >200 | 100–200 | 50–100 | <50 |
| **Unsaturated oils** | All plant oils and plant margarines. | >40 | 20–40 | 10–20 | <10 |
| **Legumes** | Dry beans, lentils, peas, soy. Peas, lentils, beans, tofu, soy containing meat replacement products. | >75 | 37.5–75 | 18.75–37.5 | <18.75 |
| **Nuts** | Peanuts or tree nuts. All nuts and seeds including peanuts, nut mixes such as almond paste. | >50 | 25–50 | 12.5–25 | <12.5 |
| **Whole grains** | Whole grains (e.g., cereals, rolled oats, crispbread) and whole grain foods (e.g., pastas, doughs and breads). | >232 | 116–232 | 58–116 | <58 |
| **Fish** | Fatty fish, lean fish, fish products, shellfish. | >28 | 14–28 | 7–14 | <7 |
| **Beef and lamb** | Beef, lamb, minced meat with pork and lamb, processed meats with beef and lamb including sausages. | <7 | 7–14 | 14–28 | >28 |
| **Pork** | Pork, minced meat of pork, processed meats with pork including ham, bacon, and sausages. | <7 | 7–14 | 14–28 | >28 |
| **Poultry** | Chicken, turkey, duck, goose, and other poultry. | <29 | 29–58 | 58–116 | >116 |
| **Eggs** | Boiled eggs, fried eggs and eggs in dishes such as omelet and pie. | <13 | 13–25 | 25–50 | >50 |
| **Dairy** | Whole milk or derivative equivalents. Regular milk, low-fat milk, yoghurt and other fermented milk products, hard cheese, soft cheese, cream, butter, butter-based spreads. All dairy foods were expressed as of milk equivalents. Equivalency factor: whole milk 1.0, Cheese 5.0, cream 2.7 and butter 6.5. | <250 | 250–500 | 500–1000 | >1000 |
| **Potatoes** | Boiled potatoes, fried potatoes, deep fried potatoes, potatoes included in dishes such as potato salad. | <50 | 50–100 | 100–200 | >200 |
| **Added sugar** | Sucrose and monosaccharides except sugars in fruits and vegetable. | <31 | 31–62 | 62–124 | >124 |

Stubbendorff A, Sonestedt E, Ramne S, Drake I, Hallström E, Ericson U. Development of an EAT-Lancet index and its relation to mortality in a Swedish population. Am J Clin Nutr. 2022;115(3):705-716.

### Healthy and Sustainable Diet Index

The Healthy and Sustainable Diet Index (HSDI), introduced by Shamah-Levy et al. in 2020, is based on national representative FFQ-derived data from Mexico in 2018-2019 (n = 11,506). The HSDI assesses 13 food components with a binary scoring system assigning one point for each component meeting recommended energy percentages. The authors found a negative relationship between HSDI and obesity in men, although no association was shown in women.

The following table presents the food components of the HSDI, proving examples of food items and a more detailed explanation of the scoring.

**Table S5. Healthy and Sustainable Diet Index**

| Food components | Food items | Scoring | |
|---|---|---|---|
| | | 1 point | 0 points |
| **Whole grains** | E.g., whole wheat bread, corn dough. | ≥32.44 | <32.44 |
| **Tubers or starchy vegetables** | Potato and sweet potato | ≤1.56 | >1.56 |
| **Vegetables** | All vegetables (e.g., turnip, tomato, carrot, zucchini, broccoli, cauliflower, green beans, cabbage, lettuce, cucumber, onion, pepper and mushrooms, etc.). | ≥3.12 | <3.12 |
| **Fruits** | All fruits (e.g., orange, apple, pear, banana, melon, watermelon, mango, pineapple, grapefruit, strawberry, grape, peach, plum, etc.). | ≥5.02 | <5.02 |
| **Milk and dairy** | Milk, cheese and yoghurt of all types. | ≤6.12 | >6.12 |
| **Beef and pork** | Beef or pork, all types of cuts and viscera. | ≤0.64 | >0.64 |
| **Chicken and other poultry** | Any type of poultry and viscera. | ≤2.48 | >2.48 |
| **Eggs** | Egg. | ≤1.00 | >1.00 |
| **Fish and seafood** | Fresh fish, dried fish, tuna, sardine, any seafood. | ≤1.60 | >1.60 |
| **Legumes, soybeans and tree nuts** | Beans, lentils, soymilk, chickpeas, peanuts. | ≥23.0 | <23.0 |
| **Unsaturated fats** | Oil from corn, sunflower, sesame, olive. | ≥14.16 | <4.16 |
| **Saturated fats** | Cream, butter, milk fat, lard, bacon. | ≤3.84 | >3.84 |
| **Added sugars** | Sweeteners added to beverages, soft drinks, natural or industrialized juices and nectars, candies, sugars in desserts, cookies or cakes, chocolates, etc. | ≤5.00 | >5.00 |

Shamah-Levy T, Gaona-Pineda EB, Mundo-Rosas V, Méndez Gómez-Humarán I, Rodríguez-Ramírez S. Asociación de un índice de dieta saludable y sostenible con sobrepeso y obesidad en adultos mexicanos [Association of a healthy and sustainable dietary index and overweight and obesity in Mexican adults]. Salud Publica Mex. 2020;62(6):745-753.

### EAT-Lancet Diet Score

The EAT-Lancet Diet Score (ELDS) was the first measure derived from the EAT-Lancet report. The recommended amount of energy intake is 2500 kcal per day, similar to the EAT- Lancet reference diet. For each food group, an intake threshold was established. The ELDS was developed using data from Food Frequency Questionnaire of 46,069 participants enrolled in the European Prospective Investigation into Cancer and Nutrition (EPIC)-Oxford study between 1993 and 2001. The ELDS assesses 14 food components using a binary scoring system: one point if the recommendation is met, otherwise zero point is allocated. The ELDS has been linked to ischemic heart disease and diabetes, although it does not show a clear relationship with mortality or stroke.

The following table presents the food components of the ELDS, proving examples of food items and a more detailed explanation of the scoring.

**Table S6. EAT-Lancet Diet Score**

| Food components | Food items | Scoring | |
|---|---|---|---|
| | | 1 point | 0 points |
| **Whole grains** | Rice, wheat, corn, and other. | ≤464 | >464 |
| **Tubers and starchy vegetables** | Potatoes and cassava. | ≤100 | >100 |
| **Vegetables** | All non-starchy vegetables. | ≥200 | <200 |
| **Fruits** | All fruit. | ≥100 | <100 |
| **Dairy foods** | Whole milk or derivative equivalents (e.g., cheese). | ≤500 | >500 |
| **Beef, lamb, pork** | Red meats of beef, lamb and pork and processed meats. | ≤28 | >28 |
| **Chicken, other poultry** | Chicken and other poultry (e.g., turkey, duck, and quail). | ≤58 | >58 |
| **Eggs** | Eggs from chicken, duck, and goose. | ≤25 | >25 |
| **Fish** | Fish and shellfish (e.g., mussels and shrimps). | ≤100 | >100 |
| **Dry beans, lentils, peas** | Dry beans, lentils, peas. | ≥100 | <100 |
| **Peanuts or tree nuts** | Almonds, hazelnuts, pistachios and walnuts. | ≥25 | <25 |
| **Added fats** | Dairy fats (e.g., cream and butter), lard or tallow. | 0.8 | 0.8 |
| **Added sugar** | Added sugars and sugar-sweetened beverages. | ≤31 | >31 |
| **Soy foods** | Soy foods (e.g., soy milk, tofu) | ≤50 | >50 |

Knuppel A, Papier K, Key TJ, Travis RC. EAT-Lancet score and major health outcomes: the EPIC-Oxford study. The Lancet. 2019;394(10194):213-4.

### PANDiet scoring system

The PANDiet is a measure of individual diet quality expressed as a 100-point score that determines the probability that overall nutrient intake is adequate [1]. The PANDiet is calculated as the mean of two sub-scores: The Adequacy Subscore (i.e., the probability of that intake to be adequate), and the Moderation Subscore (i.e., the probability of not exceeding intake of nutrients that should be limited). The total PANDiet score ranges from 0 to 100, with a higher score indicating a better nutritional quality of the diet. The following table presents the PANDiet scoring system.

**Table S7. PANDiet scoring system.**

| Adequacy subscore | | | |
|---|---|---|---|
| **Nutrient** | **RV (/day)** | **CV** | **Source** |
| **Protein** | 0.66 or 0.8 g/Kg bw | 12.5% | [2] |
| **LA** | 3.08% EIEA | 15% | [2] |
| **ALA** | 0.769% | 15% | [2] |
| **DHA** | 0.192 g | 15% | [2] |
| **EPA+DHA** | 0.385 g | 15% | [2] |
| **Fiber** | 23 g | 15% | [2] |
| **Vitamin A** | 580 or 490 µg | 15% | [2] |
| **Thiamin** | 0.3 mg/1000 kcal | 20% | [2] |
| **Riboflavin** | 1.3 mg | 15% | [2] |
| **Niacin** | 5.44 mg NE/1000 kcal | 10% | [2] |
| **Pantothenic acid** | 3.33 or 2.78 mg | 40% | [1] |
| **Vitamin B-6** | 1.5 or 1.3 mg | 10% | [2] |
| **Folate** | 250 µg | 15% | [2] |
| **Vitamin B-12** | 3.33 µg | 10% | [2] |
| **Vitamin C** | 90 mg | 10% | [2] |
| **Vitamin D** | 10 µg | 25% | [2] |
| **Vitamin E** | 5.26 or 4.74 mg | 40% | [2] |
| **Iodine** | 107 µg | 20% | [2] |
| **Magnesium** | 224 or 176 mg | 35% | [2] |
| **Phosphorus** | Calcium (mmol)/1.65 | 7.5%+CV calcium | [2] |
| **Potassium** | 2692 mg | 15% | [2] |
| **Selenium** | 54 µg | 15% | [2] |
| **Zinc** | Phytate (mg/d) / Zinc (mg) M / F<br>300 / 9.4 / 7.5<br>600 / 11.7 / 9.3<br>900 / 14.0 / 11.0<br>1200 / 16.3 / 12.7 | 10% | [3] |
| **Copper** | 0.86 or 0.68 mg | 40% | [2] |
| **Manganese** | 1.89 or 1.56 mg | 40% | [2] |
| **Calcium** | 860 (≤ 24 yo.) or 750 (>24 yo.) | 15% or 13% | [2] |
| **Iron** | 6 or 7 mg | 20% | [4] |

| Moderation subscore | | | |
|---|---|---|---|
| **Nutrient** | **RV (/day)** | **CV** | **Source** |
| **Protein** | 2.2 g/kg bw | 12.5% | [2] |
| **Carbohydrates** | 60.5% EIEA | 5% | [2] |
| **Total fat** | 44% EIEA | 5% | [2] |
| **SFA** | 12% EIEA | 15% | [2] |
| **Sugars** | 100 g | 15% | [2] |
| **Sodium** | 3312 or 2618 mg | 40% | [2] |

| Tolerable Upper Intake Limits | | Source |
|---|---|---|
| **Vitamin A (retinol only)** | 3000 µg | [2] |
| **Niacin** | 900 mg | [2] |
| **Vitamin B6** | 25 mg | [2] |
| **Folate** | 1170 µg | [2] |
| **Vitamin D** | 100 µg | [2] |
| **Vitamin E** | 300 mg | [2] |
| **Calcium** | 2500 mg | [2] |
| **Copper** | 10 mg | [2] |
| **Iodine** | 600 µg | [2] |
| **Dissociable magnesium** | 250 mg | [2] |
| **Selenium** | 300 µg | [2] |
| **Zinc** | 25 mg | [2] |

**Computation [1]**

$$F\left(\frac{\bar{y}-r}{\sqrt{SD_r^2+SD_y^2/n}}\right)$$

$F$ is the "normal" function in Stata
$\bar{y}$ is the mean intake
$SD^2_y$ is the day-to-day variability of intake
$n$ is the number of days of dietary data
$r$ is the nutrient reference value
$SD^2_r$ is the interindividual variability

*Note*. DHA and EPA+DHA are weighted by a factor of 1/2 as DHA is present twice. Niacin equivalents are calculated as the sum of dietary niacin and 1/60 dietary tryptophan. ALA, alpha-linolenic acid; bw, body weight; CV, coefficient of variation; DHA, docosahexaenoic acid; EIEA, energy intake excluding alcohol; EPA, eicosapentaenoic acid; LA, linoleic acid; NE, niacin equivalent; SFA, saturated fatty acids.

[1] Verger EO, Mariotti F, Holmes BA, Paineau D, Huneau JF. Evaluation of a Diet Quality Index Based on the Probability of Adequate Nutrient Intake (PANDiet) Using National French and US Dietary Surveys. PLoS ONE. 2012;7(8).
[2] Salomé M, Mariotti F, Nicaud MC, Dussiot A, Kesse-Guyot E, Maillard MN, Huneau JF, Fouillet H. Eropean Journal of Nutrition. 2022;61(4):1991-2002.
[3] EFSA (2014). Draft Scientific Opinion on Dietary Reference Values for Zinc. EFSA Journal 2014. Available online: https://www.efsa.europa.eu/sites/default/files/consultation/140514%2C0.pdf. Accessed 30 Nov 2023.
[4] ANSES (2021). Les références nutritionnelles en vitamines et minéraux. In: Anses—Agence nationale de sécurité sanitaire de l'alimentation, de l'environnement et du travail. Available online: https://www.anses.fr/fr/content/les-r%C3%A9f%C3%A9rences-nutritionnelles-en-vitamines-et-min%C3%A9raux. Accessed 30 Nov 2023.

### Global Diet Quality Score

The Global Diet Quality Score (GDQS) is a diet quality assessment tool focusing on 25 food groups considered critical for nutrient intake and/or risk of non-communicable diseases. The score is composed of 16 food groups categorized as healthy (scored positively to encourage higher intake), seven groups classified as unhealthy (scored negatively to penalize higher levels of intake), and two food groups that are considered unhealthy only if consumed in excessive amounts (score 0 if intake is low or excessive). The total score ranges from 0 to 49. Also, two additional GDQS-related sub-metrics are calculated: the GDQS+ (only for food groups considered healthy), which ranges from 0 to 32; and the GDQS- (only for food groups considered unhealthy or harmful in excessive amounts), which ranges from 0 to 17. The following table presents the GDQS scoring system.

**Table S8. Global Diet Quality Score**

| Food group | Food items | Categories of consumption | | | | Point values | | | |
|---|---|---|---|---|---|---|---|---|---|
| | | 1 | 2 | 3 | 4 | 1 | 2 | 3 | 4 |
| ***GDQS+*** | | | | | | | | | |
| ***Healthy*** | | | | | | | | | |
| Citrus fruits | Fruits in the genus Citrus | <24 | 24–69 | >69 | | 0 | 1 | 2 | |
| Deep orange fruits | Fruits containing ≥20 retinol equivalents/100 g | <25 | 25–123 | >123 | | 0 | 1 | 2 | |
| Other fruits | Fruits not belonging in the other fruit categories | <27 | 27–107 | >107 | | 0 | 1 | 2 | |
| Dark green leafy vegetables | Leafy vegetables containing >120 retinol equivalents/100 g | <13 | 13–37 | >37 | | 0 | 2 | 4 | |
| Cruciferous vegetables | Vegetables in the family Brassicaceae | <13 | 13–36 | >36 | | 0 | 0.25 | 0.5 | |
| Deep orange vegetables | Non-tuberous vegetables containing ≥120 retinol equivalents/100 g | <9 | 9–45 | >45 | | 0 | 0.25 | 0.5 | |
| Other vegetables | Vegetables not belonging in the other vegetable categories | <23 | 23–114 | >114 | | 0 | 0.25 | 0.5 | |
| Legumes | Legumes and foods derived from legumes. | <9 | 9–42 | >42 | | 0 | 2 | 4 | |
| Deep orange tubers | Tuberous vegetables containing ≥120 retinol equivalents/100 g | <12 | 12–63 | >63 | | 0 | 0.25 | 0.5 | |
| Nuts and seeds | Nuts, seeds, and products derived | <7 | 7–13 | >13 | | 0 | 2 | 4 | |
| Whole grains | Whole grains and whole-grain products. | <8 | 8–13 | >13 | | 0 | 1 | 2 | |
| Liquid oils | All types of oils that are liquid at room temperature. | <2 | 2–7.5 | >7.5 | | 0 | 1 | 2 | |
| Fish and shellfish | Fish (processed or unprocessed) and seafood (e.g., shellfish). | <14 | 14–71 | >71 | | 0 | 1 | 2 | |
| Poultry and game meat | Unprocessed poultry and game. | <16 | 16–44 | >44 | | 0 | 1 | 2 | |
| Low fat dairy | Reduced or naturally low-fat dairy products (≤2% milk fat). | <33 | 33–132 | >132 | | 0 | 1 | 2 | |
| Eggs | All types of eggs. Does not include mayonnaise | <6 | 6–32 | >32 | | 0 | 1 | 2 | |

| Food group | Food items | Categories of consumption | | | | Point values | | | |
|---|---|---|---|---|---|---|---|---|---|
| | | 1 | 2 | 3 | 4 | 1 | 2 | 3 | 4 |
| ***GDQS-*** | | | | | | | | | |
| ***Unhealthy in excessive amounts*** | | | | | | | | | |
| High fat dairy | High fat milk and dairy products (>2% milk fat), except butter, ice cream and whipped cream. | <35 | 35–142 | >142–734 | >734 | 0 | 1 | 2 | 0 |
| Red meat | Unprocessed red meat belonging to domesticated animals (i.e., not game), including organs. | <9 | 9–46 | >46 | | 0 | 1 | 0 | |
| ***Unhealthy*** | | | | | | | | | |
| Processed meat | Processed red meat, poultry, or game, including organs. | <9 | 9–30 | >30 | | 2 | 1 | 0 | |
| Refined grains and baked goods | Refined grains and refined grain products. | <7 | 7–33 | >33 | | 2 | 1 | 0 | |
| Sweets and ice cream | Sugar-sweetened foods that are not beverages (e.g., sugar, sweeteners, whipped cream). | <13 | 13–37 | >37 | | 2 | 1 | 0 | |
| Sugar-sweetened beverages | Sweetened drinks that do not contain any fruit juice (e.g., sodas, energy drinks, sports drinks). | <57 | 57–180 | >180 | | 2 | 1 | 0 | |
| Juice | Unsweetened or sweetened drinks that are at least partly composed of fruit juice. | <36 | 36–144 | >144 | | 2 | 1 | 0 | |
| White roots and tubers | Tuberous vegetables with <120 retinol equivalents/100 g. | <27 | 27–107 | >107 | | 2 | 1 | 0 | |
| Purchased deep fried foods | Deep fried foods fried in an amount of fat or oil sufficient to cover the food. | <9 | 9–45 | >45 | | 2 | 1 | 0 | |

Bromage S, Batis C, Bhupathiraju SN, Fawzi WW, Fung TT, Li Y, Deitchler M, Angulo E, Birk N, Castellanos-Gutiérrez A, He Y, Fang Y, Matsuzaki M, Zhang Y, Moursi M, Gicevic S, Holmes MD, Isanaka S, Kinra S, Sachs SE, Stampfer MJ, Stern D, Willett WC. Development and Validation of a Novel Food-Based Global Diet Quality Score (GDQS). J Nutr. 2021;151(12 Suppl 2):75S-92S.

#### Comprehensive Diet Quality Index

The Comprehensive Diet Quality Index (cDQI) is a recently developed measure that ranges from 0 to 85 and is obtained by adding two sub-indexes: the plant-based diet quality index (pDQI) and the animal-based diet quality index (aDQI). The main purpose of this metric is to distinguish between the consumption of plant and animal products that are considered healthy and those that are not. The cDQI aims to provide a comprehensive assessment of diet quality, considering both plant and animal components. The two sub-indices are calculated using thresholds based on literature evidence or crude consumption quintiles. The following table presents the cDQI scoring system.

**Table S9. Comprehensive Diet Quality Index**

| | | Max score (5) | Min Score (0) |
|---|---|---|---|
| **Plant-based Diet Quality Index (pDQI)** | | | |
| ***Healthful*** | | | |
| | Wholegrain products | ≥ 45g per 1000/kcal | No consumption |
| | Vegetables (excluding potatoes) | ≥ 125g per 1000/kcal | No consumption |
| | Fruit | ≥ 125g per 1000/kcal | No consumption |
| | Nuts, seeds, legumes | ≥ 14,175g per 1000/kcal | No consumption |
| | Vegetable oils | ≥ 14.86g per 1000/kcal | < 4.13g per 1000/kcal |
| | Coffee, tea | ≥477.35g per 1000/kcal | < 91.72g per 1000/kcal |
| ***Unhealthy*** | | | |
| | Fruit juices | No consumption | ≥79.38g per 1000/kcal |
| | Refined grains | <54g per 1000/kcal | ≥129g per 1000/kcal |
| | Potatoes | No consumption | ≥ 35g per 1000/kcal |
| | Sugar-sweetened beverages | No consumption | ≥ 226.8g per 1000/kcal |
| | Sweets and desserts | < 14.98g per 1000/kcal | ≥ 40.32g per 1000/kcal |
| Range pDQI | | **0 to 55** | |
| **Animal-based Diet Quality Index (aDQI)** | | | |
| ***Healthful*** | | | |
| | Fish, seafood | ≥ 14,175g per 1000/kcal | No consumption |
| | Dairy | ≥ 312g per 1000/kcal | No consumption |
| | Poultry | ≥ 17.09g per 1000/kcal | < 3.12g per 1000/kcal |
| ***Unhealthy*** | | | |
| | Processed meat | No consumption | ≥ 28,35g per 1000/kcal |
| | Red meat | No consumption | ≥ 45,36g per 1000/kcal |
| | Eggs | < 1.78g per 1000/kcal | ≥ 8.62g per 1000/kcal |
| Range aDQI | | **0 to 30** | |
| **cDQI Total Range** | | **0 to 85** | |

Brunin J, Allès B, Péneau S, Reuzé A, Pointereau P, Touvier M, et al. Do individual sustainable food purchase motives translate into an individual shift towards a more sustainable diet? A longitudinal analysis in the NutriNet-Santé cohort. Clean Responsib Consum. (2022) 5:100062.

### Dietary Inflammatory Index

The Dietary Inflammatory Index (DII) is a population-based index derived from the literature for the purpose of measuring the overall impact of diet on inflammatory potential. It is based on the individual inflammatory effects of up to 45 dietary parameters. Unlike single compound or single food approaches, a strategic advantage of the DII is that it allows the study of the dietary matrix and the complex interactions of nutrients and compounds present in foods, as well as overall dietary patterns.

The specific steps for the calculation were as follows:

1) dietary intake data were compared with the global standard dietary intake database, and a Z-score was calculated for each nutrient based on the mean and standard deviation of that nutrient intake:

$$Z\ score = \frac{individual\ reported\ intake - global\ daily\ mean\ intake}{global\ standard\ deviation}$$

2) the Z-scores were converted into centered proportions,
3) the centered proportion for each dietary parameter was multiplied by the specific inflammatory effect score to obtain a DII score,
4) DII scores were summed for all dietary parameters to obtain the total DII score.

Since its release, the DII index has undergone rigorous validations in both cross-sectional and longitudinal studies. These validations have yielded consistent results indicating that higher DII scores are linked to proinflammatory dietary patterns. In addition, a significant association has been observed between elevated DII scores and elevated plasma concentrations of proinflammatory cytokines over time and health outcomes.

The following table presents the DII scoring system for the 34 parameters used in the current study.

**Table S10. Dietary Inflammatory Index**

| Food parameter | Overall inflammatory effect score | Global daily mean intake (units/d) | SD |
|---|---|---|---|
| Alcohol (g) | −0.278 | 13.98 | 3.72 |
| Vitamin B12 (μg) | 0.106 | 5.15 | 2.70 |
| Vitamin B6 (mg) | −0.365 | 1.47 | 0.74 |
| β-Carotene (μg) | −0.584 | 3718 | 1720 |
| Carbohydrate (g) | 0.097 | 272.2 | 40.0 |
| Energy (kcal) | 0.180 | 2056 | 338 |
| Total fat (g) | 0.298 | 71.4 | 19.4 |
| Fiber (g) | −0.663 | 18.8 | 4.9 |
| Folic acid (μg) | −0.190 | 273.0 | 70.7 |
| Garlic (g) | −0.412 | 4.35 | 2.90 |
| Ginger (g) | −0.453 | 59.0 | 63.2 |
| Fe (mg) | 0.032 | 13.35 | 3.71 |
| Mg (mg) | −0.484 | 310.1 | 139.4 |
| MUFA (g) | −0.009 | 27.0 | 6.1 |
| Niacin (mg) | −0.246 | 25.90 | 11.77 |
| n-3 Fatty acids (g) | −0.436 | 1.06 | 1.06 |
| n-6 Fatty acids (g) | −0.159 | 10.80 | 7.50 |
| Onion (g) | −0.301 | 35.9 | 18.4 |
| Protein (g) | 0.021 | 79.4 | 13.9 |
| PUFA (g) | −0.337 | 13.88 | 3.76 |
| Riboflavin (mg) | −0.068 | 1.70 | 0.79 |
| Saffron (g) | −0.140 | 0.37 | 1.78 |
| Saturated fat (g) | 0.373 | 28.6 | 8.0 |
| Se (μg) | −0.191 | 67.0 | 25.1 |
| Thiamin (mg) | −0.098 | 1.70 | 0.66 |
| Turmeric (mg) | −0.785 | 533.6 | 754.3 |
| Vitamin A (RE) | −0.401 | 983.9 | 518.6 |
| Vitamin C (mg) | −0.424 | 118.2 | 43.46 |
| Vitamin D (μg) | −0.446 | 6.26 | 2.21 |
| Vitamin E (mg) | −0.419 | 8.73 | 1.49 |
| Zn (mg) | −0.313 | 9.84 | 2.19 |
| Pepper (g) | −0.131 | 10.00 | 7.07 |
| Thyme/oregano (mg) | −0.102 | 0.33 | 0.99 |
| Rosemary (mg) | −0.013 | 1.00 | 15.00 |

Shivappa N, Steck SE, Hurley TG, Hussey JR, Hébert JR. Designing and developing a literature-derived, population-based dietary inflammatory index. Public Health Nutr. 2014;17(8):1689-96.

**Composite Dietary Antioxidant Index**

The Composite Dietary Antioxidant Index (CDAI) is a comprehensive measure that assesses an individual's antioxidant profile by combining several dietary antioxidants, such as vitamins A, C and E, manganese, selenium and zinc. Its development was based on the cumulative effect of anti-inflammatory indicators related to various health problems mediated by oxidative processes.

Widely used to evaluate the antioxidant capacity of dietary components, the CDAI is calculated from the levels of vitamins A, C and E, as well as manganese, selenium and zinc. The formula used to calculate the CDAI has been previously validated and is expressed as follows:

$$CDAI = \sum_{i=1}^{n=6} \frac{Total\ nutrient\ intake - mean}{SD}$$

Here, the total nutrient intake reflects the daily intake of antioxidant for each individual; the mean represents the mean of the antioxidant in the whole sample, while the standard deviation (SD) indicates the variability of the antioxidant in the whole sample.

Wang L, Yi Z. Association of the Composite dietary antioxidant index with all-cause and cardiovascular mortality: A prospective cohort study. Front Cardiovasc Med. 2022;9:993930.

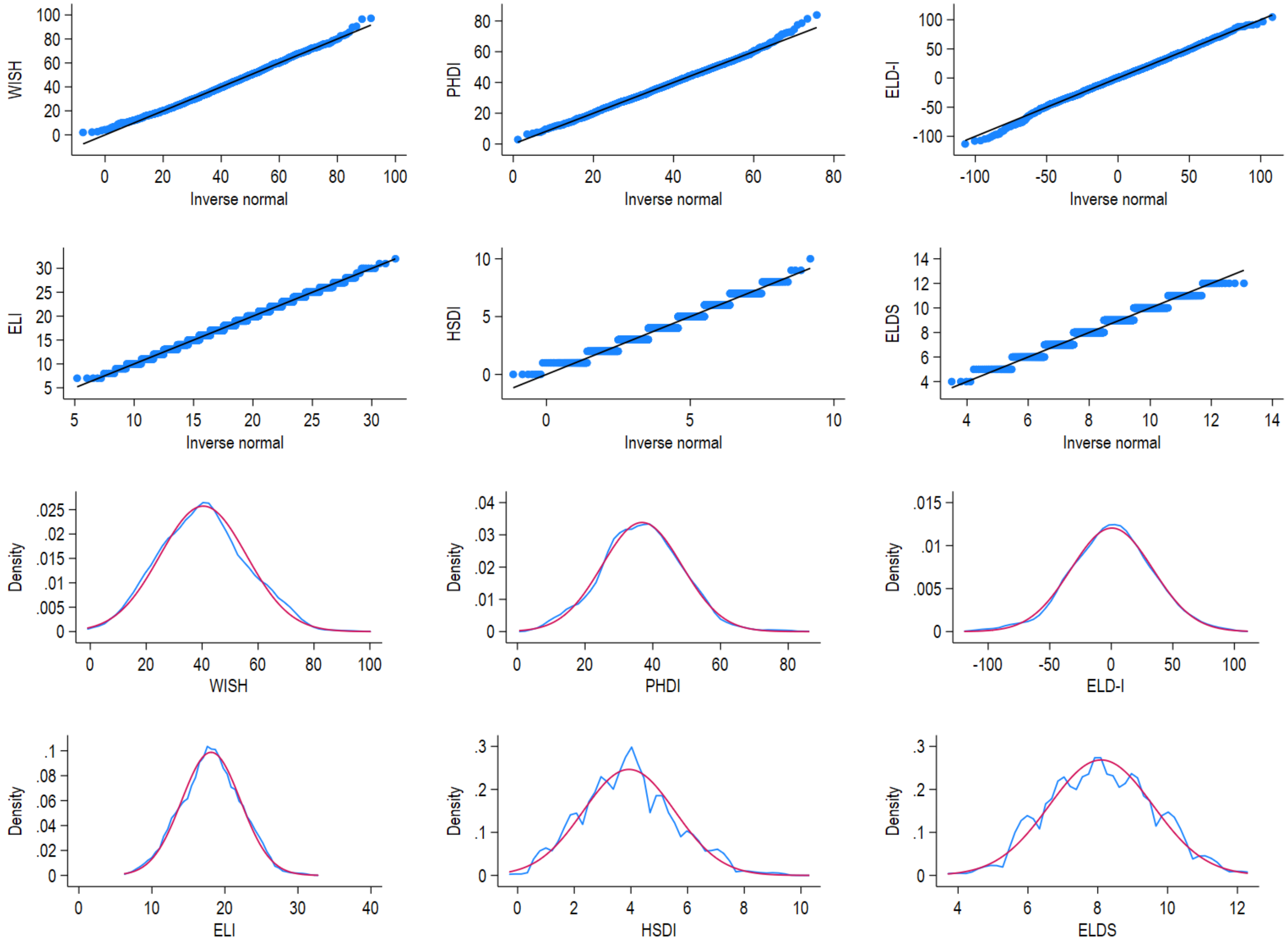

**Figure S2. Distribution plots.** QQ-plots (upper panel) and Kernel density estimates (lower panel) of normal densities.

**Table S11. Inter-item and Item-total correlation matrices of WISH**

| Food components | 1 | 2 | 3 | 4 | 5 | 6 | 7 | 8 | 9 | 10 | 11 | 12 | 13 | 14 |
|---|---|---|---|---|---|---|---|---|---|---|---|---|---|---|
| 1. Whole grains | 1 | 0.0818 | 0.3581 | 0.0014 | 0.0583 | 0.6530 | 0.1643 | 0.3422 | 0.0386 | 0.2030 | 0.0171 | 0.4462 | 0.6574 | 0.0000 |
| 2. Fruits | 0.04 | 1.00 | 0.1659 | 0.0131 | 0.3477 | 0.0039 | 0.8658 | 0.0029 | 0.0526 | 0.0592 | 0.4150 | 0.0001 | 0.0000 | 0.0000 |
| 3. Dairy foods | 0.02 | 0.03 | 1 | 0.5167 | 0.0158 | 0.5356 | 0.0029 | 0.0212 | 0.2383 | 0.0012 | 0.2458 | 0.0141 | 0.0039 | 0.0000 |
| 4. Nuts | 0.08 | 0.06 | 0.02 | 1 | 0.7977 | 0.5229 | 0.0469 | 0.1525 | 0.0000 | 0.0000 | 0.8216 | 0.0106 | 0.0008 | 0.0000 |
| 5. Unsaturated oils | 0.05 | 0.02 | -0.06 | 0.01 | 1 | 0.0000 | 0.0760 | 0.5966 | 0.3201 | 0.0024 | 0.0830 | 0.0606 | 0.0476 | 0.0000 |
| 6. Saturated oils | -0.01 | 0.07 | 0.01 | 0.02 | 0.10 | 1 | 0.0000 | 0.9292 | 0.0035 | 0.1365 | 0.0067 | 0.0000 | 0.0003 | 0.0000 |
| 7. Eggs | -0.03 | 0.00 | -0.07 | -0.05 | 0.04 | 0.13 | 1 | 0.0608 | 0.3842 | 0.9739 | 0.0000 | 0.0000 | 0.9804 | 0.0000 |
| 8. Chicken and other poultry | -0.02 | 0.07 | 0.06 | 0.03 | -0.01 | 0.00 | -0.05 | 1 | 0.0000 | 0.5026 | 0.6145 | 0.3269 | 0.3428 | 0.0000 |
| 9. Red meat | 0.05 | 0.05 | 0.03 | 0.15 | -0.02 | 0.07 | -0.02 | -0.16 | 1 | 0.0057 | 0.0008 | 0.1584 | 0.0084 | 0.0000 |
| 10. Fish | 0.03 | 0.05 | 0.08 | 0.14 | -0.07 | 0.04 | 0.00 | 0.02 | 0.07 | 1 | 0.5568 | 0.0495 | 0.1199 | 0.0000 |
| 11. Legumes | 0.06 | -0.02 | 0.03 | -0.01 | -0.04 | -0.07 | -0.10 | 0.01 | -0.08 | 0.01 | 1 | 0.0009 | 0.0952 | 0.0001 |
| 12. Added sugars | 0.02 | 0.10 | -0.06 | -0.06 | 0.05 | 0.22 | 0.13 | 0.02 | 0.03 | 0.05 | -0.08 | 1 | 0.0035 | 0.0000 |
| 13. Vegetables | 0.01 | 0.32 | 0.07 | 0.08 | 0.05 | 0.09 | 0.00 | 0.02 | 0.06 | 0.04 | 0.04 | 0.07 | 1 | 0.0000 |
| 14. WISH total score | 0.13 | 0.49 | 0.25 | 0.21 | 0.15 | 0.45 | 0.32 | 0.28 | 0.23 | 0.38 | 0.10 | 0.46 | 0.50 | 1 |

*Note*. Pearson correlation coefficients are presented in the lower hemimatrix and p-values in the upper hemimatrix.

**Table S12. Inter-item and Item-total correlation matrices of PHDI**

| Food components | 1 | 2 | 3 | 4 | 5 | 6 | 7 | 8 | 9 | 10 | 11 | 12 | 13 | 14 | 15 | 16 | 17 |
|---|---|---|---|---|---|---|---|---|---|---|---|---|---|---|---|---|---|
| 1. Nuts and peanuts | 1 | 0.3615 | 0.1406 | 0.0056 | 0.0485 | 0.9254 | 0.0048 | 0.8601 | 0.0261 | 0.3357 | 0.0000 | 0.3099 | 0.9768 | 0.4560 | 0.1164 | 0.0414 | 0.0000 |
| 2. Legumes | -0.02 | 1 | 0.1227 | 0.333 | 0.6061 | 0.6075 | 0.0909 | 0.0000 | 0.4562 | 0.7133 | 0.0407 | 0.6528 | 0.0097 | 0.0094 | 0.2480 | 0.4666 | 0.0001 |
| 3. Fruits | 0.04 | -0.04 | 1 | 0.0000 | 0.0146 | 0.3298 | 0.1798 | 0.8030 | 0.6500 | 0.0289 | 0.9164 | 0.0222 | 0.0529 | 0.0000 | 0.0000 | 0.0000 | 0.0000 |
| 4. Vegetables | 0.07 | -0.02 | 0.20 | 1 | 0.5190 | 0.9709 | 0.6802 | 0.1185 | 0.1556 | 0.6184 | 0.5198 | 0.0947 | 0.2761 | 0.0024 | 0.0000 | 0.0000 | 0.0000 |
| 5. Whole cereals | 0.05 | 0.01 | 0.06 | 0.02 | 1 | 0.1390 | 0.2186 | 0.0135 | 0.9390 | 0.0009 | 0.0924 | 0.2256 | 0.5275 | 0.0063 | 0.0853 | 0.8937 | 0.0000 |
| 6. Eggs | 0.00 | -0.01 | -0.02 | 0.00 | 0.04 | 1 | 0.9100 | 0.6957 | 0.5961 | 0.7777 | 0.0048 | 0.3979 | 0.0006 | 0.0001 | 0.9610 | 0.0049 | 0.0000 |
| 7. Fish and seafood | 0.07 | 0.04 | -0.03 | -0.01 | 0.03 | 0.00 | 1 | 0.0840 | 0.0323 | 0.0987 | 0.0473 | 0.0436 | 0.8617 | 0.0617 | 0.0387 | 0.3588 | 0.0000 |
| 8. Tubers and potatoes | 0.00 | 0.13 | -0.01 | -0.04 | 0.06 | 0.01 | 0.04 | 1 | 0.6357 | 0.3182 | 0.0949 | 0.4500 | 0.1117 | 0.0435 | 0.9336 | 0.0065 | 0.0000 |
| 9. Dairy | 0.05 | -0.02 | -0.01 | 0.03 | 0.00 | -0.01 | -0.05 | -0.01 | 1 | 0.0000 | 0.5167 | 0.5375 | 0.0260 | 0.2156 | 0.0640 | 0.0001 | 0.0000 |
| 10. Vegetable oils | 0.02 | -0.01 | -0.05 | -0.01 | -0.08 | -0.01 | 0.04 | 0.02 | 0.14 | 1 | 0.9178 | 0.8021 | 0.0000 | 0.4762 | 0.0071 | 0.1809 | 0.0000 |
| 11. Read meat | 0.11 | -0.05 | 0.00 | 0.02 | 0.04 | -0.07 | 0.05 | -0.04 | -0.02 | 0.00 | 1 | 0.1815 | 0.2534 | 0.6370 | 0.2636 | 0.1135 | 0.0000 |
| 12. Chicken and substitutes | -0.01 | -0.02 | 0.01 | 0.01 | -0.07 | -0.02 | 0.15 | 0.05 | 0.04 | -0.01 | -0.04 | 1 | 0.5156 | 0.2033 | 0.1195 | 0.0851 | 0.0000 |
| 13. Animal fats | 0.00 | -0.06 | 0.05 | 0.03 | -0.02 | -0.08 | 0.00 | -0.04 | -0.05 | 0.11 | 0.03 | 0.06 | 1 | 0.0000 | 0.0012 | 0.4424 | 0.0000 |
| 14. Added sugars | -0.02 | -0.06 | 0.13 | 0.07 | 0.07 | -0.10 | -0.05 | -0.05 | -0.03 | -0.02 | 0.01 | -0.02 | 0.24 | 1 | 0.0001 | 0.0682 | 0.0000 |
| 15. DGV/total ratio | 0.04 | 0.03 | 0.18 | 0.26 | 0.04 | 0.00 | 0.05 | 0.00 | -0.04 | 0.06 | 0.03 | -0.02 | 0.08 | 0.09 | 1 | 0.0000 | 0.0000 |
| 16. ReV/total ratio | 0.05 | -0.02 | 0.19 | 0.28 | 0.00 | -0.07 | 0.02 | 0.07 | 0.09 | 0.03 | 0.04 | 0.04 | -0.02 | 0.04 | 0.37 | 1 | 0.0000 |
| 17. PHDI total score | 0.31 | 0.09 | 0.45 | 0.40 | 0.16 | 0.25 | 0.31 | 0.19 | 0.24 | 0.29 | 0.19 | 0.36 | 0.26 | 0.25 | 0.39 | 0.39 | 1 |

*Note.* Pearson correlation coefficients are presented in the lower hemimatrix and p-values in the upper hemimatrix.

**Table S13. Inter-item and Item-total correlation matrices of ELD-I**

| Food components | 1 | 2 | 3 | 4 | 5 | 6 | 7 | 8 | 9 | 10 | 11 | 12 | 13 | 14 | 15 |
|---|---|---|---|---|---|---|---|---|---|---|---|---|---|---|---|
| 1. Whole grains | 1 | 0.0076 | 0.0022 | 0.7599 | 0.0126 | 0.0001 | 0.4525 | 0.9719 | 0.7839 | 0.1071 | 0.1346 | 0.0000 | 0.0067 | 0.0218 | 0.0039 |
| 2. Potatoes and tuber | -0.06 | 1 | 0.0000 | 0.5783 | 0.2539 | 0.3237 | 0.1697 | 0.0000 | 0.2202 | 0.4633 | 0.0005 | 0.5976 | 0.0106 | 0.0001 | 0.0000 |
| 3. Beef, lamb, pork | -0.07 | 0.17 | 1 | 0.0000 | 0.0158 | 0.0000 | 0.0521 | 0.0236 | 0.0000 | 0.0000 | 0.3693 | 0.0093 | 0.0000 | 0.0006 | 0.0000 |
| 4. Chicken and poultry | -0.01 | -0.01 | -0.24 | 1 | 0.0065 | 0.2236 | 0.1018 | 0.2311 | 0.4604 | 0.1805 | 0.0216 | 0.6033 | 0.0544 | 0.5399 | 0.9611 |
| 5. Eggs | 0.06 | -0.03 | -0.06 | -0.07 | 1 | 0.5770 | 0.9339 | 0.1711 | 0.0001 | 0.1126 | 0.0224 | 0.9693 | 0.4950 | 0.1672 | 0.0000 |
| 6. Fish | 0.09 | -0.02 | -0.15 | 0.03 | -0.01 | 1 | 0.4230 | 0.9769 | 0.0161 | 0.0000 | 0.0017 | 0.0000 | 0.1051 | 0.0729 | 0.0001 |
| 7. Legumes | 0.02 | -0.03 | 0.05 | -0.04 | 0.00 | 0.02 | 1 | 0.0986 | 0.0773 | 0.2153 | 0.8974 | 0.2922 | 0.5831 | 0.8579 | 0.0005 |
| 8. Unsaturated oils | 0.00 | 0.24 | 0.05 | -0.03 | 0.03 | 0.00 | -0.04 | 1 | 0.0000 | 0.0000 | 0.0083 | 0.0182 | 0.5577 | 0.0019 | 0.3204 |
| 9. Saturated oils | 0.01 | -0.03 | -0.13 | 0.02 | 0.10 | -0.06 | 0.04 | -0.16 | 1 | 0.0000 | 0.0000 | 0.0000 | 0.0148 | 0.0000 | 0.0000 |
| 10. Added sugars | -0.04 | -0.02 | -0.15 | -0.03 | 0.04 | -0.12 | -0.03 | -0.10 | 0.12 | 1 | 0.0000 | 0.0000 | 0.7890 | 0.0108 | 0.0000 |
| 11. Vegetables | -0.04 | 0.08 | 0.02 | -0.06 | -0.06 | -0.08 | 0.00 | -0.06 | 0.13 | 0.17 | 1 | 0.0000 | 0.0480 | 0.0000 | 0.0000 |
| 12. Fruits | -0.12 | -0.01 | 0.06 | -0.01 | 0.00 | -0.12 | 0.03 | -0.06 | 0.17 | 0.17 | 0.34 | 1 | 0.0364 | 0.0070 | 0.0000 |
| 13. Nuts | -0.07 | 0.06 | 0.16 | 0.05 | -0.02 | -0.04 | 0.01 | -0.01 | 0.06 | -0.01 | 0.05 | 0.05 | 1 | 0.5351 | 0.0000 |
| 14. Dairy foods | 0.06 | -0.10 | -0.08 | -0.01 | -0.03 | 0.04 | 0.00 | -0.07 | -0.13 | -0.06 | -0.12 | -0.07 | 0.02 | 1 | 0.0000 |
| 15. ELDI total score | -0.07 | 0.27 | 0.45 | 0.00 | 0.28 | -0.09 | 0.08 | -0.02 | 0.46 | 0.33 | 0.49 | 0.58 | 0.24 | -0.11 | 1 |

*Note*. Pearson correlation coefficients are presented in the lower hemimatrix and p-values in the upper hemimatrix.

**Table S14. Inter-item and Item-total correlation matrices of ELI**

| Food components | 1 | 2 | 3 | 4 | 5 | 6 | 7 | 8 | 9 | 10 | 11 | 12 | 13 | 14 | 15 |
|---|---|---|---|---|---|---|---|---|---|---|---|---|---|---|---|
| 1. Vegetables | 1 | 0.0000 | 0.0978 | 0.0530 | 0.0017 | 0.6018 | 0.0551 | 0.0072 | 0.0157 | 0.2384 | 0.6921 | 0.0000 | 0.0581 | 0.0001 | 0.0000 |
| 2. Fruits | 0.34 | 1 | 0.1818 | 0.8000 | 0.0100 | 0.0000 | 0.0100 | 0.9900 | 0.1300 | 0.0200 | 0.7300 | 0.0000 | 0.0600 | 0.0000 | 0.0000 |
| 3. Unsaturated oils | 0.04 | 0.03 | 1 | 0.7597 | 0.0124 | 0.1152 | 0.1199 | 0.0001 | 0.1946 | 0.3910 | 0.1208 | 0.3230 | 0.0000 | 0.7137 | 0.0000 |
| 4. Legumes | 0.05 | -0.01 | 0.01 | 1 | 0.7742 | 0.0010 | 0.1231 | 0.0062 | 0.0485 | 0.8431 | 0.0022 | 0.0083 | 0.7134 | 0.0140 | 0.0000 |
| 5. Nuts | 0.08 | 0.06 | 0.06 | 0.01 | 1 | 0.0117 | 0.0000 | 0.0000 | 0.0363 | 0.9987 | 0.0018 | 0.6073 | 0.4267 | 0.0007 | 0.0000 |
| 6. Whole grains | 0.01 | 0.09 | 0.04 | 0.08 | 0.06 | 1 | 0.0187 | 0.0304 | 0.6571 | 0.6165 | 0.2615 | 0.0255 | 0.2065 | 0.5681 | 0.0000 |
| 7. Fish | 0.05 | 0.06 | -0.04 | 0.04 | 0.1 | 0.06 | 1 | 0.0000 | 0.0000 | 0.3231 | 0.9068 | 0.0442 | 0.2104 | 0.0052 | 0.0000 |
| 8. Beef and lamb | 0.06 | 0.00 | -0.09 | -0.07 | 0.12 | 0.05 | 0.11 | 1 | 0.3824 | 0.0000 | 0.9774 | 0.0349 | 0.0000 | 0.7478 | 0.0000 |
| 9. Pork | 0.06 | 0.04 | 0.03 | -0.05 | 0.05 | 0.01 | 0.11 | -0.02 | 1 | 0.0011 | 0.4074 | 0.0059 | 0.8425 | 0.0129 | 0.0000 |
| 10. Poultry | 0.03 | 0.06 | 0.02 | 0.00 | 0.00 | -0.01 | 0.02 | -0.14 | -0.08 | 1 | 0.0727 | 0.0027 | 0.3663 | 0.1829 | 0.0000 |
| 11. Eggs | -0.01 | 0.01 | -0.04 | -0.07 | -0.08 | -0.03 | 0.00 | 0.00 | -0.02 | -0.04 | 1 | 0.3634 | 0.6756 | 0.0000 | 0.0000 |
| 12. Dairy | -0.10 | -0.12 | 0.02 | -0.06 | -0.01 | -0.05 | -0.05 | -0.05 | 0.07 | -0.07 | 0.02 | 1 | 0.1400 | 0.0007 | 0.0000 |
| 13. Potatoes | 0.05 | -0.04 | -0.19 | 0.01 | -0.02 | 0.03 | 0.03 | 0.11 | 0.00 | -0.02 | -0.01 | -0.04 | 1 | 0.0650 | 0.0000 |
| 14. Added sugar | 0.09 | 0.08 | -0.01 | -0.06 | -0.08 | -0.01 | 0.07 | 0.01 | 0.06 | 0.03 | 0.17 | 0.08 | 0.04 | 1 | 0.0000 |
| 15. ELI total score | 0.48 | 0.44 | 0.14 | 0.13 | 0.22 | 0.17 | 0.47 | 0.34 | 0.37 | 0.15 | 0.27 | 0.10 | 0.24 | 0.37 | 1 |

*Note*. Pearson correlation coefficients are presented in the lower hemimatrix and p-values in the upper hemimatrix.

**Table S15. Inter-item and Item-total correlation matrices of HSDI**

| Food components | 1 | 2 | 3 | 4 | 5 | 6 | 7 | 8 | 9 | 10 | 11 | 12 | 13 | 14 |
|---|---|---|---|---|---|---|---|---|---|---|---|---|---|---|
| 1. Whole grain foods | 1 | 0.0076 | 0.1589 | 0.3641 | 0.0092 | 0.0653 | 0.3110 | 0.3046 | 1.0000 | 1.0000 | 0.4796 | 1.0000 | 0.7189 | 0.0001 |
| 2. Tubers or starchy vegetables | 0.45 | 1 | 0.0000 | 0.0145 | 0.0240 | 0.0044 | 0.6916 | 0.0075 | 0.9604 | 0.5222 | 0.6180 | 0.1018 | 0.6438 | 0.0001 |
| 3. Vegetables | 0.29 | 0.21 | 1 | 0.0212 | 0.0000 | 0.3826 | 0.3210 | 0.9211 | 0.0010 | 0.5229 | 0.0000 | 0.0234 | 0.0003 | 0.0001 |
| 4. Milk and dairy products | 0.12 | -0.12 | -0.11 | 1 | 0.8399 | 0.4643 | 0.2575 | 0.8421 | 0.0958 | 0.0245 | 0.6387 | 0.1746 | 0.2146 | 0.0001 |
| 5. Fruits | 0.44 | 0.09 | 0.44 | 0.01 | 1 | 0.5152 | 0.8824 | 0.0547 | 0.0000 | 0.5175 | 0.0000 | 0.0814 | 0.0004 | 0.0001 |
| 6. Beef and pork | 0.33 | 0.18 | 0.06 | 0.05 | 0.04 | 1 | 0.3364 | 0.0868 | 0.0411 | 0.1051 | 0.4507 | 0.0101 | 1.0000 | 0.0001 |
| 7. Chicken and other poultry | 0.18 | -0.02 | -0.04 | -0.05 | 0.01 | -0.06 | 1 | 0.0003 | 0.4949 | 1.0000 | 0.0445 | 0.8052 | 0.0202 | 0.0001 |
| 8. Eggs | -0.19 | -0.10 | 0.00 | -0.01 | 0.07 | -0.11 | -0.14 | 1 | 0.0576 | 0.5040 | 0.0000 | 0.7121 | 0.0002 | 0.0001 |
| 9. Fish and seafood | 0.06 | 0.00 | -0.13 | 0.08 | -0.20 | -0.13 | -0.03 | -0.07 | 1 | 1.0000 | 0.0859 | 0.6211 | 0.0229 | 0.0001 |
| 10. Legumes, soy and tree nuts | 0.62 | -0.12 | 0.12 | 0.48 | -0.13 | 0.48 | -0.04 | -0.17 | -0.04 | 1 | 0.1784 | 1.0000 | 1.0000 | 0.0770 |
| 11. Saturated fats | -0.16 | 0.02 | 0.16 | 0.02 | 0.17 | 0.05 | -0.08 | 0.24 | -0.07 | 0.26 | 1 | 0.0000 | 0.0000 | 0.0001 |
| 12. Unsaturated oils | 0.08 | 0.12 | 0.17 | 0.11 | 0.12 | 0.24 | -0.02 | 0.03 | -0.04 | 0.33 | 0.37 | 1 | 0.0067 | 0.0001 |
| 13. Added sugars | -0.11 | -0.02 | 0.15 | 0.06 | 0.14 | -0.01 | -0.09 | 0.14 | -0.09 | 0.11 | 0.24 | 0.19 | 1 | 0.0001 |
| 14. HSDI total score* | 0.10 | 0.35 | 0.46 | 0.22 | 0.46 | 0.16 | 0.23 | 0.33 | 0.20 | 0.04 | 0.47 | 0.24 | 0.41 | 1 |

*Note.* Tetrachoric correlation and point-biserial correlation (*) coefficients are presented in the lower hemimatrix and p-values in the upper hemimatrix.

**Table S16. Inter-item and Item-total correlation matrices of ELDS**

| Food components | 1 | 2 | 3 | 4 | 5 | 6 | 7 | 8 | 9 | 10 | 11 | 12 | 13 | 14 | 15 |
|---|---|---|---|---|---|---|---|---|---|---|---|---|---|---|---|
| 1. Whole grains | 1 | 0.0394 | 0.0413 | 0.0000 | 0.0284 | 0.0139 | 0.7927 | 0.2983 | 0.0566 | 0.4556 | 0.0003 | 0.0289 | 0.9585 | 0.2009 | 0.0001 |
| 2. Tubers and starchy vegetables | 0.09 | 1 | 0.0004 | 0.8518 | 0.6249 | 0.0173 | 0.8773 | 0.5824 | 0.0937 | 0.5565 | 1.0000 | 0.5475 | 0.1790 | 0.8514 | 0.0001 |
| 3. Vegetables | 0.08 | 0.15 | 1 | 0.0000 | 0.4233 | 0.1887 | 0.7528 | 0.6180 | 0.8558 | 0.8126 | 0.0144 | 0.1766 | 0.1483 | 0.3990 | 0.0001 |
| 4. Fruits | 0.17 | -0.01 | 0.36 | 1 | 0.4670 | 0.1127 | 0.2384 | 0.9173 | 0.0042 | 0.4577 | 0.0652 | 0.2003 | 0.0059 | 0.5228 | 0.0001 |
| 5. Dairy foods | -0.13 | -0.04 | -0.05 | -0.05 | 1 | 0.4781 | 0.5196 | 0.9189 | 0.0902 | 1.0000 | 0.2583 | 0.7989 | 0.4139 | 0.2116 | 0.0001 |
| 6. Beef, lamb, pork | 0.12 | 0.14 | 0.06 | 0.08 | -0.05 | 1 | 0.0000 | 0.3230 | 0.0041 | 0.4973 | 0.2536 | 0.0045 | 0.6724 | 0.0088 | 0.0001 |
| 7. Chicken, other poultry | -0.01 | -0.01 | 0.02 | 0.05 | 0.04 | -0.24 | 1 | 0.5629 | 0.0767 | 0.3497 | 0.5989 | 1.0000 | 0.8471 | 0.0040 | 0.0001 |
| 8. Eggs | -0.04 | 0.03 | -0.02 | -0.01 | 0.01 | -0.05 | -0.03 | 1 | 0.4031 | 0.1483 | 0.8902 | 0.4531 | 0.0037 | 0.5309 | 0.0001 |
| 9. Fish | -0.11 | 0.10 | -0.01 | -0.17 | 0.13 | -0.19 | -0.13 | -0.05 | 1 | 0.6431 | 1.0000 | 1.0000 | 0.7099 | 0.6692 | 0.0001 |
| 10. Dry beans, lentils, peas | -0.10 | 0.09 | -0.05 | 0.09 | -0.02 | 0.19 | 0.12 | 0.16 | 0.08 | 1 | 0.4559 | 0.5278 | 0.8091 | 0.2507 | 0.0001 |
| 11. Soy foods | -0.26 | 0.01 | -0.19 | -0.16 | -0.23 | -0.09 | 0.05 | -0.02 | -0.02 | 0.09 | 1 | 0.2887 | 0.2703 | 1.0000 | 0.1409 |
| 12. Peanuts or tree nuts | 0.15 | 0.06 | 0.10 | 0.10 | -0.03 | 0.22 | 0.02 | -0.05 | 0.02 | -0.06 | -0.13 | 1 | 0.6174 | 0.0787 | 0.0001 |
| 13. Added sugars | 0.00 | 0.06 | 0.06 | 0.11 | 0.06 | 0.02 | 0.01 | 0.11 | -0.02 | 0.05 | -0.08 | 0.04 | 1 | 0.4819 | 0.0001 |
| 14. Added fats | -0.07 | -0.01 | -0.04 | -0.03 | -0.09 | -0.18 | 0.18 | 0.04 | 0.04 | 0.24 | 0.02 | -0.22 | -0.04 | 1 | 0.0001 |
| 15. ELDS total score | 0.36 | 0.35 | 0.46 | 0.45 | 0.13 | 0.24 | 0.25 | 0.33 | 0.12 | 0.11 | 0.04 | 0.18 | 0.40 | 0.19 | 1 |

*Note.* Tetrachoric correlation and point-biserial correlation (*) coefficients are presented in the lower hemimatrix and p-values in the upper hemimatrix.

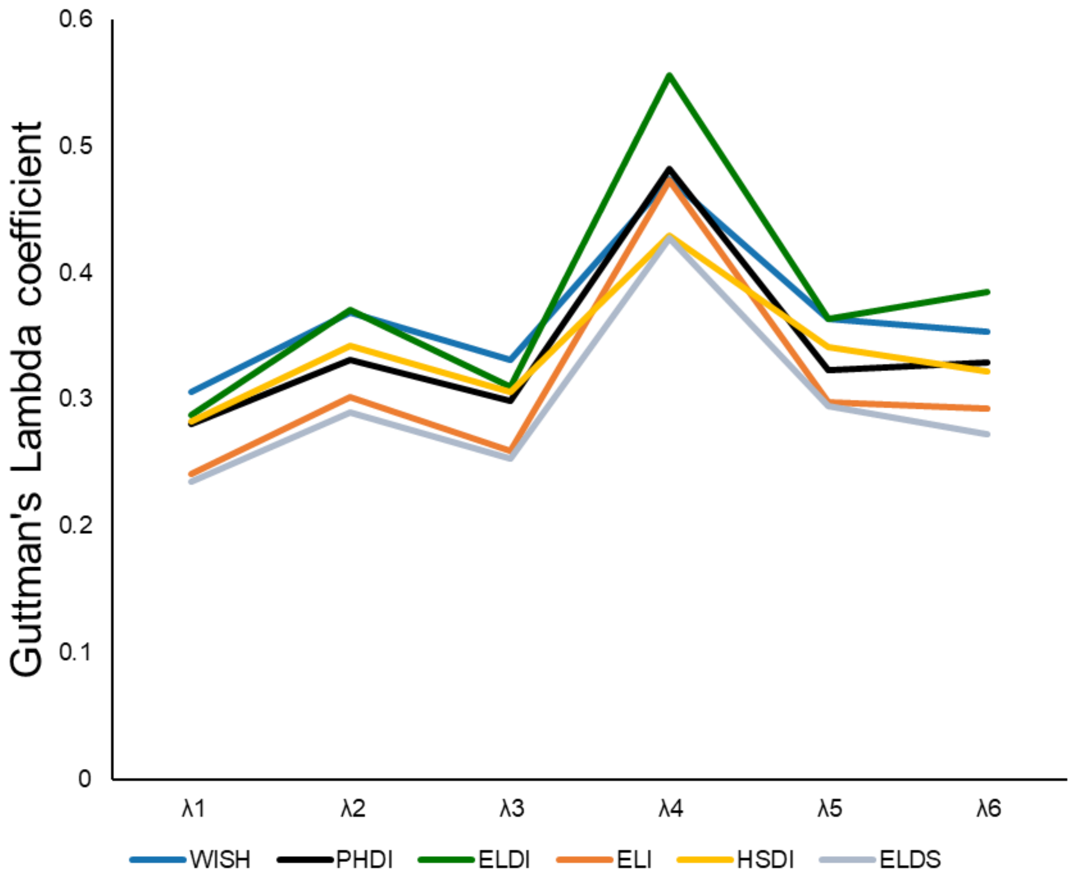

**Figure S3. Internal consistency reliability of EAT-Lancet indices.**

**Table S17. Correlation between indices and energy intake**

| | WISH | PHDI | ELDI | ELI | HSDI | ELDS | EI |
|---|---|---|---|---|---|---|---|
| **WISH** | 1 | < 0.0001 | < 0.0001 | < 0.0001 | < 0.0001 | < 0.0001 | < 0.0001 |
| **PHDI** | 0.5282 | 1 | < 0.0001 | < 0.0001 | < 0.0001 | < 0.0001 | 0.0003 |
| **ELDI** | 0.6247 | 0.4492 | 1 | < 0.0001 | < 0.0001 | < 0.0001 | < 0.0001 |
| **ELI** | 0.7513 | 0.5238 | 0.6295 | 1 | < 0.0001 | < 0.0001 | < 0.0001 |
| **HSDI** | 0.5681 | 0.5173 | 0.5407 | 0.4195 | 1 | < 0.0001 | < 0.0001 |
| **ELDS** | 0.6845 | 0.4553 | 0.5829 | 0.637 | 0.4948 | 1 | < 0.0001 |
| **Energy intake (kcal/d)** | -0.2542 | -0.0881 | -0.1066 | -0.2789 | -0.2274 | -0.3055 | 1 |

Note. Pearson correlation coefficients are presented in the lower hemimatrix and p-values in the upper hemimatrix.

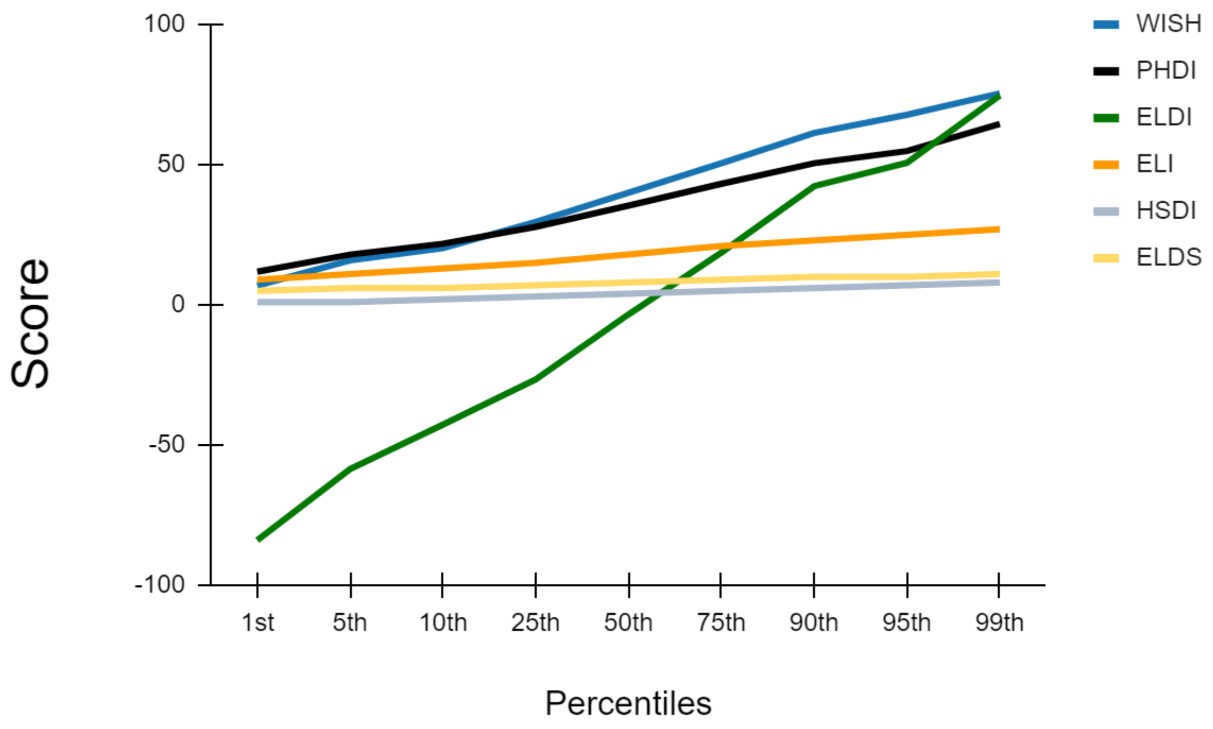

**Figure S4. Percentile distribution of the of EAT-Lancet indices.**

| Table S18. PANDiet score across quintiles of WISH | | | | | | | | |
|---|---|---|---|---|---|---|---|---|
| | **Q1** | **Q2** | **Q3** | **Q4** | **Q5** | **p** | **η2 (95%CI)** | **Trend** |
| **PANDiet score** | 62.32 (4.92) | 63.53 (4.82) | 65.53 (5.52) | 65.89 (5.01) | 67.73 (5.24) | < 0.0001 | 0.121 (0.092 — 0.148) | < 00001 |
| **AS** | 61.39 (11.87) | 61.94 (11.69) | 65.00 (13.03) | 64.23 (11.49) | 65.04 (11.77) | < 0.0001 | 0.017 (0.006 — 0.029) | < 00001 |
| **Protein** | 0.91 (0.15) | 0.90 (0.16) | 0.89 (0.21) | 0.86 (0.21) | 0.87 (0.21) | 0.0012 | 0.010 (0.002 — 0.020) | 0.3343 |
| **LA** | 0.45 (0.27) | 0.40 (0.29) | 0.44 (0.31) | 0.44 (0.32) | 0.44 (0.32) | 0.2165 | | |
| **ALA** | 0.13 (0.21) | 0.10 (0.17) | 0.08 (0.17) | 0.12 (0.21) | 0.14 (0.23) | 0.0001 | 0.014 (0.003 — 0.024) | 0.0007 |
| **DHA** | 0.12 (0.25) | 0.20 (0.30) | 0.25 (0.34) | 0.25 (0.32) | 0.33 (0.34) | < 0.0001 | 0.048 (0.028 — 0.067) | < 00001 |
| **EPA+DHA** | 0.10 (0.24) | 0.17 (0.28) | 0.22 (0.32) | 0.22 (0.31) | 0.29 (0.33) | < 0.0001 | 0.043 (0.025 — 0.062) | < 00001 |
| **Fiber** | 0.27 (0.26) | 0.33 (0.30) | 0.39 (0.33) | 0.38 (0.32) | 0.43 (0.30) | < 0.0001 | 0.032 (0.016 — 0.048) | < 00001 |
| **Vitamin A** | 0.42 (0.31) | 0.48 (0.32) | 0.50 (0.33) | 0.51 (0.32) | 0.48 (0.33) | 0.0016 | 0.010 (0.002 — 0.019) | 0.0380 |
| **Thiamin** | 0.92 (0.10) | 0.93 (0.09) | 0.94 (0.11) | 0.95 (0.08) | 0.96 (0.07) | < 0.0001 | 0.024 (0.011 — 0.039) | < 00001 |
| **Riboflavin** | 0.73 (0.27) | 0.75 (0.26) | 0.80 (0.26) | 0.76 (0.27) | 0.74 (0.29) | 0.016 | 0.007 (0.000 — 0.015) | 0.1720 |
| **Niacin** | 1.00 (0.00) | 1.00 (0.04) | 1.00 (0.01) | 1.00 (0.04) | 1.00 (0.03) | 0.4173 | | |
| **Pantothenic acid** | 0.88 (0.13) | 0.87 (0.13) | 0.88 (0.17) | 0.88 (0.15) | 0.87 (0.16) | 0.5537 | | |
| **Vitamin B-6** | 0.64 (0.31) | 0.66 (0.30) | 0.73 (0.29) | 0.71 (0.29) | 0.71 (0.28) | < 0.0001 | 0.015 (0.004 — 0.026) | 0.0002 |
| **Folate** | 0.61 (0.32) | 0.61 (0.32) | 0.69 (0.32) | 0.70 (0.29) | 0.75 (0.25) | < 0.0001 | 0.029 (0.014 — 0.045) | < 00001 |
| **Vitamin B-12** | 0.61 (0.32) | 0.66 (0.33) | 0.72 (0.30) | 0.64 (0.30) | 0.66 (0.32) | 0.0001 | 0.013 (0.003 — 0.024) | 0.1378 |
| **Vitamin C** | 0.31 (0.35) | 0.37 (0.37) | 0.45 (0.38) | 0.50 (0.36) | 0.57 (0.36) | < 0.0001 | 0.061 (0.040 — 0.082) | < 00001 |
| **Vitamin D** | 0.02 (0.07) | 0.03 (0.08) | 0.03 (0.07) | 0.04 (0.11) | 0.05 (0.11) | 0.0004 | 0.012 (0.003 — 0.022) | < 00001 |
| **Vitamin E** | 0.85 (0.17) | 0.81 (0.17) | 0.85 (0.19) | 0.83 (0.18) | 0.86 (0.15) | 0.0054 | 0.009 (0.001 — 0.017) | < 00001 |
| **Iodine** | 0.67 (0.28) | 0.68 (0.29) | 0.74 (0.28) | 0.71 (0.27) | 0.76 (0.25) | < 0.0001 | 0.015 (0.004 — 0.026) | < 00001 |
| **Magnesium** | 0.86 (0.15) | 0.87 (0.13) | 0.89 (0.15) | 0.88 (0.13) | 0.88 (0.15) | 0.0047 | 0.009 (0.001 — 0.017) | 0.0011 |
| **Phosphorus** | 1.00 (0.00) | 1.00 (0.01) | 1.00 (0.00) | 1.00 (0.02) | 1.00 (0.00) | 0.5079 | | |
| **Potassium** | 0.61 (0.32) | 0.61 (0.31) | 0.69 (0.30) | 0.68 (0.29) | 0.68 (0.29) | < 0.0001 | 0.015 (0.004 — 0.026) | 0.7435 |
| **Selenium** | 0.95 (0.09) | 0.92 (0.14) | 0.94 (0.16) | 0.95 (0.08) | 0.95 (0.14) | 0.014 | 0.007 (0.000 — 0.015) | < 00001 |
| **Zinc** | 0.45 (0.31) | 0.41 (0.34) | 0.45 (0.34) | 0.36 (0.32) | 0.29 (0.31) | < 0.0001 | 0.035 (0.018 — 0.052) | 0.0103 |
| **Copper** | 0.87 (0.13) | 0.86 (0.13) | 0.88 (0.14) | 0.88 (0.14) | 0.87 (0.14) | 0.3206 | | |
| **Manganese** | 0.85 (0.17) | 0.84 (0.17) | 0.85 (0.18) | 0.86 (0.16) | 0.85 (0.18) | 0.4562 | | |
| **Calcium** | 0.63 (0.33) | 0.62 (0.34) | 0.67 (0.34) | 0.64 (0.32) | 0.68 (0.31) | 0.0493 | 0.006 (0.000 — 0.012) | 0.0666 |
| **Iron** | 0.82 (0.22) | 0.83 (0.20) | 0.82 (0.24) | 0.82 (0.21) | 0.78 (0.25) | 0.0454 | 0.006 (0.000 — 0.013) | 0.5863 |
| **MS** | 63.24 (9.37) | 65.11 (9.24) | 66.06 (10.66) | 67.55 (9.79) | 70.41 (9.67) | < 0.0001 | 0.056 (0.036 — 0.077) | < 00001 |
| **Protein** | 0.94 (0.13) | 0.93 (0.14) | 0.95 (0.13) | 0.96 (0.10) | 0.97 (0.09) | 0.0002 | 0.013 (0.003 — 0.023) | 0.5299 |
| **Carbohydrates** | 0.88 (0.13) | 0.86 (0.15) | 0.89 (0.15) | 0.88 (0.16) | 0.88 (0.15) | 0.1959 | | |
| **Total fat** | 0.84 (0.17) | 0.87 (0.16) | 0.86 (0.17) | 0.85 (0.18) | 0.85 (0.19) | 0.4772 | | |
| **SFA** | 0.29 (0.23) | 0.30 (0.23) | 0.31 (0.24) | 0.34 (0.28) | 0.41 (0.29) | < 0.0001 | 0.025 (0.011 — 0.039) | < 00001 |
| **Sugars** | 0.43 (0.33) | 0.51 (0.34) | 0.56 (0.36) | 0.58 (0.34) | 0.66 (0.32) | < 0.0001 | 0.050 (0.031 — 0.070) | < 00001 |
| **Sodium** | 0.42 (0.26) | 0.43 (0.25) | 0.40 (0.26) | 0.43 (0.26) | 0.45 (0.27) | 0.1861 | | |

Note. Data presented as mean (standard deviation). Q = quintile.

**Table S19. PANDiet score across quintiles of PHDI**

| | Q1 | Q2 | Q3 | Q4 | Q5 | p | η2 (95%CI) | Trend |
|---|---|---|---|---|---|---|---|---|
| **PANDiet score** | 63.59 (5.24) | 65.11 (5.12) | 65.24 (5.23) | 65.33 (5.71) | 66.54 (5.38) | < 00001 | 0.059 (0.038 ─ 0.081) | < 00001 |
| **AS** | 60.52 (13.24) | 63.74 (10.67) | 63.56 (11.75) | 64.08 (11.99) | 66.18 (11.98) | < 00001 | 0.022 (0.010 ─ 0.037) | < 00001 |
| **Protein** | 0.90 (0.18) | 0.90 (0.18) | 0.88 (0.19) | 0.90 (0.17) | 0.87 (0.23) | 0.0915 | | |
| **LA** | 0.39 (0.26) | 0.41 (0.29) | 0.42 (0.30) | 0.45 (0.32) | 0.52 (0.31) | < 00001 | 0.021 (0.008 ─ 0.034) | < 00001 |
| **ALA** | 0.11 (0.20) | 0.12 (0.19) | 0.10 (0.18) | 0.13 (0.21) | 0.12 (0.22) | 0.4103 | | |
| **DHA** | 0.20 (0.31) | 0.25 (0.33) | 0.23 (0.32) | 0.22 (0.32) | 0.20 (0.30) | 0.1484 | | |
| **EPA+DHA** | 0.18 (0.30) | 0.22 (0.31) | 0.20 (0.31) | 0.20 (0.30) | 0.18 (0.28) | 0.3398 | | |
| **Fiber** | 0.27 (0.27) | 0.31 (0.27) | 0.36 (0.30) | 0.39 (0.32) | 0.48 (0.35) | < 00001 | 0.053 (0.032 ─ 0.073) | < 00001 |
| **Vitamin A** | 0.40 (0.32) | 0.50 (0.32) | 0.49 (0.31) | 0.51 (0.33) | 0.49 (0.33) | < 00001 | 0.015 (0.005 ─ 0.027) | 0.0027 |
| **Thiamin** | 0.92 (0.12) | 0.93 (0.1) | 0.95 (0.07) | 0.95 (0.08) | 0.95 (0.08) | < 00001 | 0.022 (0.009 ─ 0.036) | 0.0005 |
| **Riboflavin** | 0.74 (0.29) | 0.79 (0.22) | 0.75 (0.26) | 0.74 (0.28) | 0.77 (0.30) | 0.0765 | | |
| **Niacin** | 1.00 (0.03) | 1.00 (0.00) | 1.00 (0.03) | 1.00 (0.04) | 1.00 (0.01) | 0.5041 | | |
| **Pantothenic acid** | 0.87 (0.16) | 0.88 (0.13) | 0.87 (0.15) | 0.89 (0.13) | 0.88 (0.16) | 0.4201 | | |
| **Vitamin B-6** | 0.63 (0.34) | 0.71 (0.26) | 0.70 (0.28) | 0.68 (0.29) | 0.74 (0.28) | < 00001 | 0.016 (0.005 ─ 0.028) | < 00001 |
| **Folate** | 0.57 (0.34) | 0.67 (0.30) | 0.67 (0.29) | 0.71 (0.29) | 0.74 (0.28) | < 00001 | 0.036 (0.019 ─ 0.053) | < 00001 |
| **Vitamin B-12** | 0.66 (0.32) | 0.69 (0.31) | 0.64 (0.30) | 0.62 (0.32) | 0.68 (0.32) | 0.0409 | 0.006 (0.000 ─ 0.013) | 0.6497 |
| **Vitamin C** | 0.30 (0.36) | 0.41 (0.37) | 0.44 (0.36) | 0.49 (0.37) | 0.56 (0.36) | < 00001 | 0.051 (0.031 ─ 0.071) | < 00001 |
| **Vitamin D** | 0.03 (0.08) | 0.03 (0.09) | 0.04 (0.10) | 0.04 (0.10) | 0.03 (0.08) | 0.1868 | | |
| **Vitamin E** | 0.79 (0.21) | 0.82 (0.17) | 0.85 (0.17) | 0.87 (0.15) | 0.89 (0.16) | < 00001 | 0.033 (0.017 ─ 0.050) | < 00001 |
| **Iodine** | 0.70 (0.28) | 0.71 (0.28) | 0.71 (0.28) | 0.73 (0.27) | 0.70 (0.28) | 0.6559 | | |
| **Magnesium** | 0.84 (0.17) | 0.89 (0.11) | 0.88 (0.14) | 0.88 (0.15) | 0.90 (0.13) | < 00001 | 0.024 (0.010 ─ 0.038) | < 00001 |
| **Phosphorus** | 1.00 (0.01) | 1.00 (0.00) | 1.00 (0.01) | 1.00 (0.02) | 1.00 (0.01) | 0.6008 | | |
| **Potassium** | 0.58 (0.33) | 0.68 (0.26) | 0.66 (0.30) | 0.63 (0.31) | 0.72 (0.29) | < 00001 | 0.023 (0.010 ─ 0.037) | < 00001 |
| **Selenium** | 0.93 (0.17) | 0.93 (0.12) | 0.94 (0.13) | 0.96 (0.08) | 0.96 (0.11) | 0.0006 | 0.011 (0.002 ─ 0.021) | 0.0456 |
| **Zinc** | 0.43 (0.32) | 0.43 (0.33) | 0.37 (0.32) | 0.35 (0.32) | 0.40 (0.35) | 0.0015 | 0.010 (0.002 ─ 0.020) | < 00001 |
| **Copper** | 0.85 (0.15) | 0.86 (0.13) | 0.88 (0.14) | 0.88 (0.13) | 0.91 (0.12) | < 00001 | 0.021 (0.008 ─ 0.034) | < 00001 |
| **Manganese** | 0.80 (0.21) | 0.83 (0.17) | 0.87 (0.14) | 0.87 (0.16) | 0.89 (0.15) | < 00001 | 0.033 (0.017 ─ 0.049) | < 00001 |
| **Calcium** | 0.64 (0.32) | 0.65 (0.34) | 0.63 (0.34) | 0.65 (0.34) | 0.66 (0.32) | 0.7134 | | |
| **Iron** | 0.80 (0.24) | 0.81 (0.21) | 0.82 (0.20) | 0.80 (0.23) | 0.85 (0.23) | 0.1066 | | |
| **MS** | 64.67 (10.54) | 66.48 (9.96) | 66.92 (9.33) | 66.58 (9.78) | 66.89 (10.39) | 0.0104 | 0.008 (0.001 ─ 0.016) | 0.0366 |
| **Protein** | 0.92 (0.16) | 0.95 (0.12) | 0.96 (0.10) | 0.97 (0.09) | 0.96 (0.10) | < 00001 | 0.023 (0.010 ─ 0.038) | 0.6091 |
| **Carbohydrates** | 0.87 (0.15) | 0.88 (0.14) | 0.87 (0.15) | 0.88 (0.16) | 0.90 (0.14) | 0.1697 | | |
| **Total fat** | 0.87 (0.17) | 0.86 (0.17) | 0.87 (0.16) | 0.84 (0.18) | 0.83 (0.19) | 0.0051 | 0.009 (0.001 ─ 0.017) | 0.0096 |
| **SFA** | 0.30 (0.24) | 0.32 (0.26) | 0.34 (0.25) | 0.33 (0.26) | 0.34 (0.29) | 0.2523 | | |
| **Sugars** | 0.48 (0.35) | 0.55 (0.35) | 0.55 (0.34) | 0.57 (0.36) | 0.57 (0.35) | 0.0041 | 0.009 (0.001 ─ 0.018) | 0.0318 |
| **Sodium** | 0.44 (0.27) | 0.44 (0.25) | 0.42 (0.26) | 0.41 (0.25) | 0.42 (0.29) | 0.4908 | | |

Note. Data presented as mean (standard deviation). Q = quintile.

| **Table S20. PANDiet score across quintiles of ELD-I** | | | | | | | | |
|---|---|---|---|---|---|---|---|---|
| | **Q1** | **Q2** | **Q3** | **Q4** | **Q5** | **P** | **η2 (95%CI)** | **Trend** |
| **PANDiet score** | 62.97 (5.76) | 64.28 (5.27) | 65.26 (5.24) | 65.97 (4.66) | 66.36 (5.00) | < 00001 | 0.053 (0.033 ─ 0.073) | < 00001 |
| **AS** | 0.60 (0.13) | 0.63 (0.12) | 0.65 (0.10) | 0.66 (0.10) | 0.64 (0.13) | < 00001 | 0.026 (0.011 ─ 0.040) | < 00001 |
| **Protein** | 0.88 (0.21) | 0.91 (0.16) | 0.92 (0.14) | 0.89 (0.18) | 0.84 (0.24) | < 00001 | 0.017 (0.006 ─ 0.029) | 0.3432 |
| **LA** | 0.44 (0.28) | 0.50 (0.29) | 0.42 (0.29) | 0.39 (0.30) | 0.40 (0.32) | < 00001 | 0.017 (0.006 ─ 0.030) | 0.1223 |
| **ALA** | 0.11 (0.20) | 0.14 (0.20) | 0.10 (0.18) | 0.11 (0.21) | 0.11 (0.20) | 0.1301 | | |
| **DHA** | 0.22 (0.31) | 0.17 (0.29) | 0.21 (0.32) | 0.27 (0.33) | 0.25 (0.33) | 0.0011 | 0.011 (0.002 ─ 0.020) | 0.0001 |
| **EPA+DHA** | 0.19 (0.29) | 0.15 (0.27) | 0.19 (0.32) | 0.23 (0.32) | 0.22 (0.32) | 0.0030 | 0.009 (0.001 ─ 0.018) | < 00001 |
| **Fiber** | 0.24 (0.27) | 0.30 (0.30) | 0.37 (0.30) | 0.42 (0.29) | 0.49 (0.32) | < 00001 | 0.079 (0.055 ─ 0.103) | < 00001 |
| **Vitamin A** | 0.42 (0.32) | 0.44 (0.32) | 0.49 (0.33) | 0.52 (0.31) | 0.54 (0.33) | < 00001 | 0.020 (0.008 ─ 0.034) | < 00001 |
| **Thiamin** | 0.92 (0.11) | 0.94 (0.10) | 0.94 (0.09) | 0.94 (0.09) | 0.95 (0.08) | < 00001 | 0.019 (0.007 ─ 0.032) | < 00001 |
| **Riboflavin** | 0.72 (0.28) | 0.75 (0.28) | 0.79 (0.24) | 0.81 (0.23) | 0.72 (0.31) | < 00001 | 0.018 (0.006 ─ 0.030) | 0.2640 |
| **Niacin** | 1.00 (0.03) | 1.00 (0.02) | 1.00 (0.00) | 1.00 (0.00) | 0.99 (0.05) | 0.0220 | 0.007 (0.000 ─ 0.014) | 0.0073 |
| **Pantothenic acid** | 0.85 (0.17) | 0.88 (0.13) | 0.90 (0.11) | 0.90 (0.12) | 0.86 (0.18) | < 00001 | 0.018 (0.006 ─ 0.031) | 0.0859 |
| **Vitamin B-6** | 0.65 (0.32) | 0.67 (0.31) | 0.71 (0.27) | 0.74 (0.26) | 0.71 (0.29) | 0.0004 | 0.012 (0.003 ─ 0.022) | < 00001 |
| **Folate** | 0.53 (0.35) | 0.64 (0.31) | 0.72 (0.28) | 0.74 (0.25) | 0.75 (0.27) | < 00001 | 0.073 (0.049 ─ 0.095) | < 00001 |
| **Vitamin B-12** | 0.69 (0.32) | 0.66 (0.33) | 0.69 (0.30) | 0.67 (0.29) | 0.57 (0.34) | < 00001 | 0.019 (0.007 ─ 0.032) | 0.0002 |
| **Vitamin C** | 0.25 (0.34) | 0.38 (0.37) | 0.43 (0.35) | 0.52 (0.35) | 0.64 (0.33) | < 00001 | 0.127 (0.098 ─ 0.155) | < 00001 |
| **Vitamin D** | 0.03 (0.07) | 0.03 (0.07) | 0.04 (0.11) | 0.04 (0.10) | 0.03 (0.08) | 0.0223 | 0.007 (0.000 ─ 0.014) | 0.6472 |
| **Vitamin E** | 0.80 (0.21) | 0.86 (0.16) | 0.86 (0.15) | 0.85 (0.17) | 0.85 (0.17) | < 00001 | 0.020 (0.007 ─ 0.032) | < 00001 |
| **Iodine** | 0.65 (0.29) | 0.69 (0.29) | 0.73 (0.26) | 0.76 (0.25) | 0.73 (0.28) | < 00001 | 0.020 (0.007 ─ 0.033) | < 00001 |
| **Magnesium** | 0.85 (0.16) | 0.87 (0.14) | 0.89 (0.11) | 0.89 (0.13) | 0.88 (0.15) | 0.0001 | 0.014 (0.004 ─ 0.024) | < 00001 |
| **Phosphorus** | 1.00 (0.01) | 1.00 (0.01) | 1.00 (0.00) | 1.00 (0.01) | 1.00 (0.02) | 0.6737 | | |
| **Potassium** | 0.59 (0.33) | 0.65 (0.31) | 0.66 (0.28) | 0.70 (0.27) | 0.67 (0.30) | < 00001 | 0.015 (0.004 ─ 0.027) | < 00001 |
| **Selenium** | 0.93 (0.14) | 0.95 (0.12) | 0.94 (0.14) | 0.94 (0.13) | 0.96 (0.10) | 0.0727 | | |
| **Zinc** | 0.53 (0.32) | 0.45 (0.34) | 0.38 (0.32) | 0.34 (0.31) | 0.24 (0.28) | < 00001 | 0.090 (0.064 ─ 0.114) | < 00001 |
| **Copper** | 0.84 (0.15) | 0.88 (0.12) | 0.89 (0.13) | 0.89 (0.12) | 0.88 (0.13) | < 00001 | 0.023 (0.009 ─ 0.036) | < 00001 |
| **Manganese** | 0.77 (0.21) | 0.85 (0.17) | 0.88 (0.13) | 0.89 (0.14) | 0.87 (0.16) | < 00001 | 0.064 (0.042 ─ 0.085) | < 00001 |
| **Calcium** | 0.57 (0.35) | 0.64 (0.33) | 0.68 (0.32) | 0.69 (0.33) | 0.67 (0.31) | < 00001 | 0.017 (0.006 ─ 0.029) | 0.0001 |
| **Iron** | 0.80 (0.25) | 0.85 (0.20) | 0.85 (0.19) | 0.83 (0.19) | 0.76 (0.27) | < 00001 | 0.021 (0.008 ─ 0.034) | 0.9655 |
| **MS** | 0.66 (0.11) | 0.65 (0.10) | 0.66 (0.10) | 0.66 (0.09) | 0.69 (0.09) | 0.0004 | 0.012 (0.003 ─ 0.022) | 0.0007 |
| **Protein** | 0.91 (0.17) | 0.95 (0.10) | 0.96 (0.10) | 0.95 (0.10) | 0.98 (0.07) | < 00001 | 0.038 (0.020 ─ 0.055) | 0.0085 |
| **Carbohydrates** | 0.89 (0.13) | 0.91 (0.13) | 0.87 (0.15) | 0.88 (0.15) | 0.84 (0.18) | < 00001 | 0.024 (0.010 ─ 0.038) | 0.0001 |
| **Total fat** | 0.81 (0.19) | 0.85 (0.17) | 0.87 (0.16) | 0.88 (0.17) | 0.89 (0.16) | < 00001 | 0.031 (0.016 ─ 0.047) | < 00001 |
| **SFA** | 0.29 (0.24) | 0.29 (0.24) | 0.29 (0.23) | 0.35 (0.26) | 0.43 (0.30) | < 00001 | 0.042 (0.024 ─ 0.060) | < 00001 |
| **Sugars** | 0.55 (0.35) | 0.51 (0.37) | 0.55 (0.34) | 0.53 (0.34) | 0.56 (0.33) | 0.2676 | | |
| **Sodium** | 0.49 (0.27) | 0.41 (0.26) | 0.41 (0.26) | 0.39 (0.24) | 0.42 (0.28) | < 00001 | 0.018 (0.006 ─ 0.030) | < 00001 |

Note. Data presented as mean (standard deviation). Q = quintile.

| Table S21. PANDiet score across quintiles of ELI | | | | | | | | |
|---|---|---|---|---|---|---|---|---|
| | **Q1** | **Q2** | **Q3** | **Q4** | **Q5** | **P** | **η2 (95%CI)** | **Trend** |
| **PANDiet score** | 62.92 (4.98) | 64.05 (5.18) | 65.96 (5.20) | 65.75 (5.44) | 67.51 (5.50) | < 00001 | 0.083 (0.058 ─ 0.107) | < 00001 |
| **AS** | 0.62 (0.12) | 0.62 (0.13) | 0.66 (0.12) | 0.64 (0.11) | 0.64 (0.12) | 0.0001 | 0.014 (0.004 ─ 0.025) | 0.0020 |
| **Protein** | 0.93 (0.13) | 0.85 (0.22) | 0.91 (0.18) | 0.89 (0.17) | 0.86 (0.22) | < 00001 | 0.031 (0.015 ─ 0.046) | 0.0002 |
| **LA** | 0.43 (0.27) | 0.41 (0.29) | 0.44 (0.31) | 0.40 (0.31) | 0.50 (0.32) | 0.0026 | 0.009 (0.001 ─ 0.019) | 0.0076 |
| **ALA** | 0.12 (0.19) | 0.09 (0.17) | 0.13 (0.22) | 0.10 (0.19) | 0.15 (0.23) | 0.0009 | 0.011 (0.002 ─ 0.021) | < 00001 |
| **DHA** | 0.07 (0.20) | 0.21 (0.31) | 0.24 (0.32) | 0.30 (0.33) | 0.42 (0.36) | < 00001 | 0.116 (0.088 ─ 0.143) | < 00001 |
| **EPA+DHA** | 0.06 (0.18) | 0.18 (0.29) | 0.22 (0.32) | 0.26 (0.32) | 0.38 (0.36) | < 00001 | 0.108 (0.080 ─ 0.134) | < 00001 |
| **Fiber** | 0.30 (0.30) | 0.31 (0.28) | 0.41 (0.33) | 0.41 (0.31) | 0.42 (0.31) | < 00001 | 0.031 (0.015 ─ 0.047) | < 00001 |
| **Vitamin A** | 0.45 (0.34) | 0.47 (0.33) | 0.49 (0.32) | 0.52 (0.31) | 0.49 (0.32) | 0.1306 | | |
| **Thiamin** | 0.93 (0.09) | 0.92 (0.11) | 0.95 (0.07) | 0.93 (0.10) | 0.96 (0.07) | < 00001 | 0.021 (0.008 ─ 0.035) | 0.0001 |
| **Riboflavin** | 0.76 (0.26) | 0.76 (0.26) | 0.80 (0.27) | 0.74 (0.28) | 0.70 (0.30) | 0.0003 | 0.012 (0.003 ─ 0.023) | < 00001 |
| **Niacin** | 1.00 (0.01) | 1.00 (0.03) | 1.00 (0.00) | 1.00 (0.05) | 1.00 (0.04) | 0.3479 | | |
| **Pantothenic acid** | 0.89 (0.13) | 0.87 (0.15) | 0.90 (0.16) | 0.88 (0.13) | 0.85 (0.17) | 0.0002 | 0.012 (0.003 ─ 0.023) | 0.0003 |
| **Vitamin B-6** | 0.67 (0.30) | 0.68 (0.31) | 0.74 (0.29) | 0.71 (0.25) | 0.66 (0.30) | 0.0040 | 0.009 (0.001 ─ 0.018) | 0.7934 |
| **Folate** | 0.62 (0.33) | 0.61 (0.33) | 0.72 (0.29) | 0.75 (0.25) | 0.72 (0.27) | < 00001 | 0.036 (0.019 ─ 0.052) | < 00001 |
| **Vitamin B-12** | 0.65 (0.32) | 0.66 (0.31) | 0.69 (0.31) | 0.64 (0.33) | 0.64 (0.32) | 0.1816 | | |
| **Vitamin C** | 0.31 (0.36) | 0.39 (0.37) | 0.50 (0.37) | 0.57 (0.35) | 0.52 (0.37) | < 00001 | 0.062 (0.041 ─ 0.084) | < 00001 |
| **Vitamin D** | 0.02 (0.06) | 0.03 (0.08) | 0.03 (0.08) | 0.06 (0.12) | 0.04 (0.12) | < 00001 | 0.023 (0.010 ─ 0.037) | < 00001 |
| **Vitamin E** | 0.82 (0.19) | 0.82 (0.19) | 0.86 (0.16) | 0.85 (0.16) | 0.89 (0.12) | < 00001 | 0.022 (0.009 ─ 0.035) | < 00001 |
| **Iodine** | 0.68 (0.27) | 0.69 (0.29) | 0.75 (0.27) | 0.74 (0.26) | 0.72 (0.28) | 0.0063 | 0.008 (0.001 ─ 0.017) | 0.0426 |
| **Magnesium** | 0.87 (0.13) | 0.87 (0.15) | 0.89 (0.15) | 0.87 (0.14) | 0.88 (0.15) | 0.0852 | | |
| **Phosphorus** | 1.00 (0.01) | 1.00 (0.00) | 1.00 (0.01) | 1.00 (0.02) | 1.00 (0.01) | 0.6414 | | |
| **Potassium** | 0.64 (0.31) | 0.63 (0.32) | 0.70 (0.29) | 0.67 (0.27) | 0.64 (0.30) | 0.0052 | 0.009 (0.001 ─ 0.017) | 0.5545 |
| **Selenium** | 0.95 (0.11) | 0.93 (0.14) | 0.95 (0.12) | 0.94 (0.11) | 0.94 (0.14) | 0.0535 | | |
| **Zinc** | 0.51 (0.32) | 0.44 (0.33) | 0.42 (0.33) | 0.31 (0.30) | 0.17 (0.25) | < 00001 | 0.111 (0.084 ─ 0.138) | < 00001 |
| **Copper** | 0.87 (0.12) | 0.87 (0.13) | 0.89 (0.14) | 0.87 (0.15) | 0.87 (0.13) | 0.2748 | | |
| **Manganese** | 0.83 (0.17) | 0.84 (0.17) | 0.87 (0.18) | 0.84 (0.18) | 0.87 (0.15) | 0.0095 | 0.008 (0.001 ─ 0.016) | < 00001 |
| **Calcium** | 0.67 (0.31) | 0.63 (0.34) | 0.68 (0.34) | 0.66 (0.33) | 0.58 (0.34) | 0.0026 | 0.009 (0.001 ─ 0.019) | < 00001 |
| **Iron** | 0.85 (0.19) | 0.81 (0.23) | 0.85 (0.22) | 0.78 (0.24) | 0.76 (0.26) | < 00001 | 0.021 (0.008 ─ 0.034) | 0.0001 |
| **MS** | 0.63 (0.10) | 0.66 (0.10) | 0.66 (0.09) | 0.67 (0.11) | 0.71 (0.09) | < 00001 | 0.056 (0.035 ─ 0.077) | < 00001 |
| **Protein** | 0.94 (0.13) | 0.94 (0.13) | 0.96 (0.10) | 0.94 (0.13) | 0.98 (0.06) | < 00001 | 0.017 (0.006 ─ 0.029) | 0.0016 |
| **Carbohydrates** | 0.87 (0.14) | 0.87 (0.15) | 0.91 (0.13) | 0.85 (0.17) | 0.90 (0.15) | < 00001 | 0.017 (0.006 ─ 0.029) | 0.0198 |
| **Total fat** | 0.85 (0.17) | 0.86 (0.17) | 0.85 (0.17) | 0.87 (0.17) | 0.84 (0.19) | 0.6511 | | |
| **SFA** | 0.28 (0.23) | 0.31 (0.25) | 0.31 (0.23) | 0.40 (0.31) | 0.40 (0.27) | < 00001 | 0.032 (0.016 ─ 0.048) | < 00001 |
| **Sugars** | 0.45 (0.34) | 0.56 (0.34) | 0.54 (0.36) | 0.56 (0.35) | 0.67 (0.31) | < 00001 | 0.037 (0.020 ─ 0.054) | < 00001 |
| **Sodium** | 0.41 (0.26) | 0.43 (0.26) | 0.39 (0.27) | 0.41 (0.24) | 0.49 (0.27) | 0.0002 | 0.013 (0.003 ─ 0.023) | 0.0282 |

Note. Data presented as mean (standard deviation). Q = quintile.

**Table S22. PANDiet score across quartiles of HSDI**

| | Q1 | Q2 | Q3 | Q4 | P | η2 (95%CI) | Trend |
|---|---|---|---|---|---|---|---|
| **PANDiet score** | 0.64 (0.05) | 0.65 (0.05) | 0.66 (0.06) | 0.66 (0.06) | < 00001 | 0.026 (0.012 ─ 0.042) | < 00001 |
| **AS** | 0.63 (0.12) | 0.64 (0.12) | 0.64 (0.12) | 0.63 (0.13) | 0.7841 | | |
| **Protein** | 0.92 (0.15) | 0.89 (0.18) | 0.88 (0.21) | 0.83 (0.25) | < 00001 | 0.026 (0.012 ─ 0.042) | < 00001 |
| **LA** | 0.45 (0.29) | 0.42 (0.29) | 0.42 (0.30) | 0.43 (0.33) | 0.1980 | | |
| **ALA** | 0.12 (0.20) | 0.12 (0.20) | 0.10 (0.18) | 0.12 (0.21) | 0.3095 | | |
| **DHA** | 0.25 (0.32) | 0.25 (0.33) | 0.16 (0.28) | 0.16 (0.30) | < 00001 | 0.018 (0.007 ─ 0.031) | < 00001 |
| **EPA+DHA** | 0.22 (0.31) | 0.22 (0.32) | 0.14 (0.27) | 0.15 (0.28) | < 00001 | 0.014 (0.004 ─ 0.026) | < 00001 |
| **Fiber** | 0.30 (0.28) | 0.37 (0.31) | 0.40 (0.32) | 0.43 (0.33) | < 00001 | 0.027 (0.013 ─ 0.042) | < 00001 |
| **Vitamin A** | 0.47 (0.31) | 0.48 (0.33) | 0.48 (0.33) | 0.47 (0.33) | 0.9027 | | |
| **Thiamin** | 0.92 (0.10) | 0.94 (0.09) | 0.95 (0.08) | 0.95 (0.08) | 0.0001 | 0.013 (0.004 ─ 0.024) | 0.0001 |
| **Riboflavin** | 0.77 (0.25) | 0.76 (0.27) | 0.77 (0.27) | 0.72 (0.31) | 0.0848 | | |
| **Niacin** | 1.00 (0.00) | 1.00 (0.01) | 1.00 (0.04) | 0.99 (0.05) | 0.0013 | 0.009 (0.002 ─ 0.019) | 0.0001 |
| **Pantothenic acid** | 0.89 (0.12) | 0.88 (0.15) | 0.87 (0.16) | 0.85 (0.19) | 0.0007 | 0.010 (0.002 ─ 0.020) | 0.0002 |
| **Vitamin B-6** | 0.68 (0.29) | 0.70 (0.29) | 0.71 (0.29) | 0.67 (0.31) | 0.3808 | | |
| **Folate** | 0.63 (0.32) | 0.69 (0.30) | 0.68 (0.29) | 0.71 (0.29) | 0.0002 | 0.012 (0.003 ─ 0.022) | 0.0049 |
| **Vitamin B-12** | 0.68 (0.31) | 0.67 (0.30) | 0.66 (0.31) | 0.58 (0.34) | 0.0001 | 0.013 (0.004 ─ 0.024) | < 00001 |
| **Vitamin C** | 0.32 (0.35) | 0.48 (0.38) | 0.47 (0.37) | 0.58 (0.35) | < 00001 | 0.071 (0.048 ─ 0.094) | < 00001 |
| **Vitamin D** | 0.04 (0.09) | 0.03 (0.08) | 0.04 (0.11) | 0.02 (0.07) | 0.1821 | | |
| **Vitamin E** | 0.84 (0.17) | 0.84 (0.18) | 0.84 (0.17) | 0.84 (0.18) | 0.9552 | | |
| **Iodine** | 0.72 (0.27) | 0.71 (0.28) | 0.71 (0.28) | 0.67 (0.31) | 0.0334 | 0.005 (0.000 ─ 0.012) | 0.0209 |
| **Magnesium** | 0.87 (0.14) | 0.87 (0.14) | 0.89 (0.14) | 0.87 (0.16) | 0.4355 | | |
| **Phosphorus** | 1.00 (0.01) | 1.00 (0.00) | 1.00 (0.01) | 1.00 (0.02) | 0.5941 | | |
| **Potassium** | 0.64 (0.30) | 0.65 (0.31) | 0.67 (0.29) | 0.66 (0.32) | 0.4193 | | |
| **Selenium** | 0.95 (0.12) | 0.94 (0.14) | 0.94 (0.14) | 0.94 (0.12) | 0.4628 | | |
| **Zinc** | 0.41 (0.33) | 0.41 (0.33) | 0.36 (0.32) | 0.38 (0.35) | 0.1243 | | |
| **Copper** | 0.87 (0.13) | 0.87 (0.14) | 0.88 (0.13) | 0.88 (0.13) | 0.2273 | | |
| **Manganese** | 0.85 (0.17) | 0.83 (0.18) | 0.86 (0.17) | 0.87 (0.15) | 0.0255 | 0.005 (0.000 ─ 0.013) | 0.0328 |
| **Calcium** | 0.67 (0.32) | 0.66 (0.33) | 0.61 (0.35) | 0.60 (0.34) | 0.0026 | 0.008 (0.001 ─ 0.017) | 0.0010 |
| **Iron** | 0.83 (0.21) | 0.81 (0.24) | 0.83 (0.21) | 0.79 (0.25) | 0.0433 | 0.005 (0.000 ─ 0.012) | 0.0295 |
| **MS** | 0.64 (0.10) | 0.66 (0.11) | 0.68 (0.10) | 0.69 (0.09) | < 00001 | 0.031 (0.016 ─ 0.047) | < 00001 |
| **Protein** | 0.94 (0.13) | 0.94 (0.14) | 0.97 (0.07) | 0.97 (0.08) | < 00001 | 0.017 (0.006 ─ 0.029) | 0.3136 |
| **Carbohydrates** | 0.89 (0.13) | 0.87 (0.16) | 0.88 (0.13) | 0.86 (0.17) | 0.0080 | 0.007 (0.001 ─ 0.015) | 0.3111 |
| **Total fat** | 0.84 (0.17) | 0.87 (0.17) | 0.86 (0.17) | 0.85 (0.18) | 0.0152 | 0.006 (0.000 ─ 0.014) | 0.2586 |
| **SFA** | 0.28 (0.24) | 0.34 (0.27) | 0.36 (0.25) | 0.37 (0.28) | < 00001 | 0.021 (0.009 ─ 0.035) | < 00001 |
| **Sugars** | 0.50 (0.35) | 0.51 (0.36) | 0.59 (0.34) | 0.66 (0.31) | < 00001 | 0.031 (0.016 ─ 0.048) | < 00001 |
| **Sodium** | 0.42 (0.26) | 0.45 (0.26) | 0.42 (0.25) | 0.42 (0.29) | 0.1895 | | |

Note. Data presented as mean (standard deviation). Q = quartile.

| Table S23. PANDiet score across quartiles of ELDS | | | | | | | |
|---|---|---|---|---|---|---|---|
| | Q1 | Q2 | Q3 | Q4 | P | η2 (95%CI) | Trend |
| **PANDiet score** | 0.64 (0.05) | 0.65 (0.05) | 0.65 (0.06) | 0.66 (0.05) | < 00001 | 0.027 (0.013 ─ 0.042) | < 00001 |
| **AS** | 0.63 (0.12) | 0.64 (0.12) | 0.62 (0.13) | 0.64 (0.10) | 0.0352 | 0.005 (0.000 ─ 0.012) | 0.8075 |
| **Protein** | 0.90 (0.17) | 0.92 (0.16) | 0.86 (0.23) | 0.86 (0.22) | < 00001 | 0.016 (0.006 ─ 0.029) | 0.0003 |
| **LA** | 0.48 (0.28) | 0.41 (0.30) | 0.41 (0.32) | 0.39 (0.30) | < 00001 | 0.016 (0.006 ─ 0.028) | 0.0201 |
| **ALA** | 0.12 (0.19) | 0.10 (0.19) | 0.12 (0.20) | 0.12 (0.21) | 0.3060 | | |
| **DHA** | 0.22 (0.32) | 0.23 (0.33) | 0.20 (0.30) | 0.24 (0.32) | 0.2644 | | |
| **EPA+DHA** | 0.19 (0.30) | 0.21 (0.31) | 0.17 (0.28) | 0.22 (0.31) | 0.1921 | | |
| **Fiber** | 0.32 (0.29) | 0.35 (0.33) | 0.37 (0.31) | 0.41 (0.30) | 0.0003 | 0.011 (0.003 ─ 0.021) | < 00001 |
| **Vitamin A** | 0.43 (0.32) | 0.51 (0.32) | 0.49 (0.33) | 0.51 (0.32) | 0.0002 | 0.012 (0.003 ─ 0.022) | 0.0044 |
| **Thiamin** | 0.92 (0.10) | 0.94 (0.10) | 0.94 (0.09) | 0.95 (0.08) | < 00001 | 0.019 (0.007 ─ 0.032) | < 00001 |
| **Riboflavin** | 0.76 (0.26) | 0.78 (0.27) | 0.74 (0.28) | 0.74 (0.28) | 0.0593 | | |
| **Niacin** | 1.00 (0.00) | 1.00 (0.01) | 0.99 (0.05) | 1.00 (0.03) | 0.0183 | 0.006 (0.000 ─ 0.014) | 0.0002 |
| **Pantothenic acid** | 0.89 (0.12) | 0.88 (0.15) | 0.85 (0.18) | 0.88 (0.14) | 0.0001 | 0.013 (0.004 ─ 0.024) | 0.0021 |
| **Vitamin B-6** | 0.70 (0.30) | 0.70 (0.29) | 0.65 (0.31) | 0.70 (0.27) | 0.0641 | | |
| **Folate** | 0.63 (0.32) | 0.67 (0.32) | 0.65 (0.30) | 0.75 (0.25) | < 00001 | 0.018 (0.007 ─ 0.031) | 0.0002 |
| **Vitamin B-12** | 0.68 (0.31) | 0.71 (0.29) | 0.60 (0.33) | 0.60 (0.32) | < 00001 | 0.021 (0.009 ─ 0.035) | < 00001 |
| **Vitamin C** | 0.35 (0.37) | 0.44 (0.38) | 0.45 (0.37) | 0.56 (0.35) | < 00001 | 0.040 (0.022 ─ 0.058) | < 00001 |
| **Vitamin D** | 0.04 (0.09) | 0.03 (0.10) | 0.03 (0.09) | 0.03 (0.07) | 0.7361 | | |
| **Vitamin E** | 0.85 (0.17) | 0.85 (0.18) | 0.81 (0.20) | 0.84 (0.15) | 0.0049 | 0.007 (0.001 ─ 0.016) | 0.8071 |
| **Iodine** | 0.71 (0.27) | 0.72 (0.29) | 0.68 (0.28) | 0.72 (0.28) | 0.1101 | | |
| **Magnesium** | 0.87 (0.14) | 0.88 (0.14) | 0.86 (0.16) | 0.88 (0.13) | 0.1107 | | |
| **Phosphorus** | 1.00 (0.01) | 1.00 (0.01) | 1.00 (0.02) | 1.00 (0.00) | 0.6774 | | |
| **Potassium** | 0.66 (0.30) | 0.66 (0.31) | 0.61 (0.32) | 0.66 (0.28) | 0.0220 | 0.006 (0.000 ─ 0.013) | 0.4565 |
| **Selenium** | 0.94 (0.13) | 0.94 (0.15) | 0.94 (0.12) | 0.95 (0.10) | 0.3633 | | |
| **Zinc** | 0.46 (0.31) | 0.44 (0.35) | 0.36 (0.33) | 0.26 (0.29) | < 00001 | 0.048 (0.030 ─ 0.068) | < 00001 |
| **Copper** | 0.88 (0.12) | 0.88 (0.14) | 0.86 (0.15) | 0.88 (0.13) | 0.3073 | | |
| **Manganese** | 0.84 (0.17) | 0.85 (0.17) | 0.85 (0.19) | 0.86 (0.15) | 0.1162 | | |
| **Calcium** | 0.65 (0.32) | 0.66 (0.34) | 0.62 (0.33) | 0.64 (0.33) | 0.3766 | | |
| **Iron** | 0.84 (0.20) | 0.83 (0.23) | 0.78 (0.25) | 0.80 (0.23) | 0.0002 | 0.011 (0.003 ─ 0.022) | < 00001 |
| **MS** | 0.64 (0.10) | 0.66 (0.10) | 0.67 (0.09) | 0.69 (0.09) | < 00001 | 0.028 (0.014 ─ 0.044) | < 00001 |
| **Protein** | 0.92 (0.16) | 0.96 (0.10) | 0.96 (0.09) | 0.98 (0.07) | < 00001 | 0.041 (0.024 ─ 0.059) | < 00001 |
| **Carbohydrates** | 0.88 (0.14) | 0.88 (0.14) | 0.87 (0.16) | 0.88 (0.15) | 0.5520 | | |
| **Total fat** | 0.85 (0.17) | 0.86 (0.17) | 0.86 (0.18) | 0.85 (0.19) | 0.6586 | | |
| **SFA** | 0.32 (0.25) | 0.32 (0.26) | 0.33 (0.26) | 0.33 (0.26) | 0.9018 | | |
| **Sugars** | 0.46 (0.34) | 0.55 (0.36) | 0.59 (0.34) | 0.63 (0.33) | < 00001 | 0.036 (0.019 ─ 0.053) | < 00001 |
| **Sodium** | 0.43 (0.26) | 0.40 (0.27) | 0.42 (0.27) | 0.46 (0.25) | 0.0119 | 0.006 (0.000 ─ 0.015) | 0.4962 |

Note. Data presented as mean (standard deviation). Q = quartile.

**Table S24. Environmental impact across quintiles of WISH**

| | Q1 | Q2 | Q3 | Q4 | Q5 | P | η2 (95%CI) | Trend |
|---|---|---|---|---|---|---|---|---|
| **Product Environmental Footprint** | 0.75 (0.24) | 0.72 (0.28) | 0.77 (0.29) | 0.71 (0.21) | 0.68 (0.23) | < 00001 | 0.015 (0.005 ─ 0.027) | 0.0098 |
| **Greenhouse gas emission** | 5.93 (2.09) | 5.59 (2.45) | 6.02 (2.69) | 5.36 (1.79) | 5.01 (1.91) | < 00001 | 0.026 (0.012 ─ 0.041) | < 0.0001 |
| **Ozone depletion** | 0.73 (0.37) | 0.69 (0.32) | 0.70 (0.34) | 0.67 (0.27) | 0.65 (0.28) | 0.0165 | 0.007 (0.000 ─ 0.015) | 0.0063 |
| **Ionizing radiation** | 1.24 (0.38) | 1.17 (0.34) | 1.23 (0.38) | 1.15 (0.32) | 1.11 (0.33) | < 00001 | 0.018 (0.006 ─ 0.031) | 0.0001 |
| **Photochemical ozone formation** | 17.33 (9.94) | 17.26 (10.73) | 19.42 (10.71) | 18.72 (10.09) | 17.81 (9.06) | 0.0148 | 0.007 (0.000 ─ 0.015) | < 0.0001 |
| **Particulate matter emissions** | 0.58 (0.22) | 0.55 (0.26) | 0.60 (0.27) | 0.54 (0.20) | 0.50 (0.20) | < 00001 | 0.020 (0.008 ─ 0.033) | 0.0014 |
| **Acidification** | 0.08 (0.03) | 0.07 (0.04) | 0.08 (0.04) | 0.07 (0.03) | 0.07 (0.03) | < 00001 | 0.022 (0.009 ─ 0.035) | 0.0003 |
| **Terrestrial eutrophication** | 25.74 (8.96) | 24.86 (10.67) | 27.31 (12.23) | 25.17 (9.43) | 23.79 (8.69) | 0.0002 | 0.013 (0.003 ─ 0.023) | 0.4082 |
| **Freshwater eutrophication** | 0.33 (0.13) | 0.31 (0.15) | 0.33 (0.16) | 0.29 (0.11) | 0.27 (0.12) | < 00001 | 0.026 (0.012 ─ 0.041) | < 0.0001 |
| **Marine eutrophication** | 0.83 (0.24) | 0.79 (0.30) | 0.81 (0.26) | 0.78 (0.29) | 0.76 (0.31) | 0.0082 | 0.008 (0.001 ─ 0.016) | < 0.0001 |
| **Land use** | 80.79 (27.05) | 74.52 (28.22) | 77.04 (26.22) | 73.27 (22.15) | 70.60 (26.31) | < 00001 | 0.018 (0.006 ─ 0.030) | < 0.0001 |
| **Freshwater ecotoxicity** | 348.88 (152.00) | 332.04 (177.91) | 355.03 (182.90) | 315.68 (135.85) | 296.69 (154.76) | < 00001 | 0.017 (0.005 ─ 0.029) | < 0.0001 |
| **Water use** | 7.20 (3.87) | 7.76 (4.72) | 7.99 (4.16) | 8.25 (3.97) | 9.13 (5.01) | < 00001 | 0.021 (0.008 ─ 0.034) | < 0.0001 |
| **Fossils resource** | 57.82 (17.76) | 55.09 (17.02) | 58.86 (17.81) | 55.84 (14.28) | 54.08 (15.16) | 0.0007 | 0.011 (0.002 ─ 0.021) | 0.2178 |
| **Metals and minerals resource** | 31.67 (12.09) | 30.35 (12.96) | 31.67 (12.64) | 29.79 (9.60) | 28.94 (9.99) | 0.0067 | 0.008 (0.001 ─ 0.017) | 0.0012 |
| **Ratio PANDiet/PEF** | 0.90 (0.26) | 0.99 (0.34) | 0.97 (0.34) | 1.00 (0.29) | 1.11 (0.38) | < 00001 | 0.040 (0.023 ─ 0.058) | < 0.0001 |

*Note*. Data presented as mean (standard deviation). Q = quintile.

**Table S25. Environmental impact across quintiles of PHDI**

| | Q1 | Q2 | Q3 | Q4 | Q5 | p | η2 (95%CI) | Trend |
|---|---|---|---|---|---|---|---|---|
| **Product Environmental Footprint** | 0.75 (0.31) | 0.74 (0.25) | 0.71 (0.23) | 0.70 (0.22) | 0.75 (0.26) | 0.0361 | 0.006 (0.000 – 0.013) | 0.4293 |
| **Greenhouse gas emission** | 5.95 (2.67) | 5.62 (2.17) | 5.40 (1.91) | 5.30 (1.99) | 5.77 (2.37) | 0.0006 | 0.011 (0.002 – 0.021) | 0.0408 |
| **Ozone depletion** | 0.72 (0.36) | 0.72 (0.31) | 0.71 (0.34) | 0.63 (0.27) | 0.66 (0.30) | 0.0002 | 0.013 (0.003 – 0.023) | 0.0007 |
| **Ionizing radiation** | 1.23 (0.43) | 1.18 (0.32) | 1.16 (0.31) | 1.13 (0.33) | 1.20 (0.37) | 0.0012 | 0.011 (0.002 – 0.020) | 0.0365 |
| **Photochemical ozone formation** | 17.80 (11.14) | 19.53 (10.82) | 18.39 (10.96) | 17.16 (8.81) | 17.36 (7.95) | 0.0158 | 0.007 (0.000 – 0.015) | 0.8221 |
| **Particulate matter emissions** | 0.58 (0.27) | 0.57 (0.23) | 0.54 (0.21) | 0.52 (0.20) | 0.57 (0.25) | 0.0061 | 0.008 (0.001 – 0.017) | 0.0409 |
| **Acidification** | 0.08 (0.04) | 0.08 (0.03) | 0.07 (0.03) | 0.07 (0.03) | 0.08 (0.03) | 0.0045 | 0.009 (0.001 – 0.018) | 0.0233 |
| **Terrestrial eutrophication** | 26.61 (12.78) | 25.44 (9.16) | 25.30 (9.57) | 24.05 (8.50) | 25.68 (9.63) | 0.0220 | 0.007 (0.000 – 0.014) | 0.2968 |
| **Freshwater eutrophication** | 0.32 (0.16) | 0.31 (0.14) | 0.29 (0.12) | 0.29 (0.12) | 0.31 (0.15) | 0.0016 | 0.010 (0.002 – 0.019) | 0.0159 |
| **Marine eutrophication** | 0.83 (0.31) | 0.79 (0.25) | 0.78 (0.26) | 0.76 (0.26) | 0.82 (0.30) | 0.0108 | 0.008 (0.000 – 0.016) | 0.0626 |
| **Land use** | 76.51 (30.56) | 75.26 (24.45) | 75.43 (23.71) | 73.09 (25.27) | 77.90 (26.70) | 0.2290 | | |
| **Freshwater ecotoxicity** | 345.14 (176.41) | 322.32 (163.75) | 320.73 (143.57) | 318.64 (154.27) | 354.03 (174.56) | 0.0119 | 0.008 (0.000 – 0.016) | 0.9026 |
| **Water use** | 6.77 (4.03) | 8.19 (4.66) | 8.18 (4.61) | 8.54 (4.31) | 8.71 (3.92) | < 0.0001 | 0.026 (0.012 – 0.041) | < 0.0001 |
| **Fossils resource** | 57.63 (20.63) | 57.26 (15.04) | 55.98 (15.40) | 54.13 (14.88) | 57.18 (15.73) | 0.0429 | 0.006 (0.000 – 0.013) | 0.2954 |
| **Metals and minerals resource** | 31.06 (12.77) | 31.20 (11.95) | 30.60 (11.79) | 28.96 (10.36) | 30.99 (10.93) | 0.0863 | | |
| **Ratio PANDiet/PEF** | 0.97 (0.37) | 0.97 (0.28) | 1.00 (0.29) | 1.03 (0.32) | 1.00 (0.36) | 0.1236 | | |

*Note*. Data presented as mean (standard deviation). Q = quintile.

| Table S26. Environmental impact across quintiles of ELD-I | | | | | | | | |
|---|---|---|---|---|---|---|---|---|
| | **Q1** | **Q2** | **Q3** | **Q4** | **Q5** | **p** | **η2 (95%CI)** | **Trend** |
| **Product Environmental Footprint** | 0.84 (0.33) | 0.74 (0.24) | 0.72 (0.22) | 0.70 (0.22) | 0.62 (0.18) | < 00001 | 0.078 (0.054 ─ 0.101) | < 00001 |
| **Greenhouse gas emission** | 6.82 (2.98) | 5.69 (1.94) | 5.52 (1.90) | 5.15 (1.73) | 4.51 (1.43) | < 00001 | 0.118 (0.089 ─ 0.144) | < 00001 |
| **Ozone depletion** | 0.75 (0.34) | 0.70 (0.34) | 0.67 (0.31) | 0.70 (0.28) | 0.62 (0.32) | < 00001 | 0.016 (0.005 ─ 0.028) | < 00001 |
| **Ionizing radiation** | 1.24 (0.39) | 1.24 (0.38) | 1.17 (0.32) | 1.17 (0.36) | 1.07 (0.29) | < 00001 | 0.028 (0.013 ─ 0.043) | < 00001 |
| **Photochemical ozone formation** | 20.12 (11.41) | 17.57 (9.32) | 18.14 (11.17) | 18.49 (8.85) | 15.53 (8.88) | < 00001 | 0.021 (0.009 ─ 0.035) | 0.0045 |
| **Particulate matter emissions** | 0.68 (0.29) | 0.56 (0.21) | 0.55 (0.21) | 0.51 (0.18) | 0.44 (0.17) | < 00001 | 0.114 (0.086 ─ 0.140) | < 00001 |
| **Acidification** | 0.09 (0.04) | 0.08 (0.03) | 0.07 (0.03) | 0.07 (0.03) | 0.06 (0.02) | < 00001 | 0.128 (0.099 ─ 0.156) | < 00001 |
| **Terrestrial eutrophication** | 29.13 (12.02) | 26.21 (11.16) | 25.10 (8.47) | 23.84 (7.63) | 21.67 (8.44) | < 00001 | 0.061 (0.040 ─ 0.083) | < 00001 |
| **Freshwater eutrophication** | 0.39 (0.18) | 0.31 (0.12) | 0.30 (0.11) | 0.27 (0.10) | 0.23 (0.09) | < 00001 | 0.140 (0.110 ─ 0.169) | < 00001 |
| **Marine eutrophication** | 0.86 (0.30) | 0.82 (0.29) | 0.79 (0.26) | 0.78 (0.25) | 0.72 (0.26) | < 00001 | 0.028 (0.013 ─ 0.043) | < 00001 |
| **Land use** | 79.86 (29.14) | 77.62 (27.04) | 75.35 (24.83) | 74.06 (23.77) | 69.39 (24.67) | < 00001 | 0.018 (0.006 ─ 0.030) | < 00001 |
| **Freshwater ecotoxicity** | 412.73 (218.12) | 334.72 (132.59) | 325.55 (134.91) | 301.57 (142.57) | 256.90 (101.85) | < 00001 | 0.102 (0.075 ─ 0.127) | < 00001 |
| **Water use** | 6.89 (4.31) | 7.67 (4.13) | 7.86 (3.64) | 9.20 (5.23) | 8.86 (4.12) | < 00001 | 0.037 (0.020 ─ 0.054) | < 00001 |
| **Fossils resource** | 59.49 (19.00) | 57.47 (17.43) | 56.05 (15.58) | 56.70 (15.53) | 51.55 (13.40) | < 00001 | 0.024 (0.010 ─ 0.038) | < 00001 |
| **Metals and minerals resource** | 32.74 (12.94) | 31.22 (13.05) | 29.80 (10.35) | 31.10 (10.99) | 27.31 (9.34) | < 00001 | 0.024 (0.010 ─ 0.038) | < 00001 |
| **Ratio PANDiet/PEF** | 0.86 (0.34) | 0.95 (0.29) | 0.99 (0.29) | 1.03 (0.31) | 1.16 (0.34) | < 00001 | 0.088 (0.062 ─ 0.112) | < 00001 |

*Note*. Data presented as mean (standard deviation). Q = quintile.

**Table S27. Environmental impact across quintiles of ELI**

| | Q1 | Q2 | Q3 | Q4 | Q5 | p | η2 (95%CI) | Trend |
|---|---|---|---|---|---|---|---|---|
| **Product Environmental Footprint** | 0.78 (0.25) | 0.75 (0.29) | 0.74 (0.26) | 0.70 (0.22) | 0.62 (0.19) | < 0.0001 | 0.037 (0.020 ─ 0.054) | < 0.0001 |
| **Greenhouse gas emission** | 6.21 (2.15) | 5.84 (2.59) | 5.69 (2.22) | 5.15 (1.85) | 4.45 (1.56) | < 0.0001 | 0.064 (0.042 ─ 0.085) | < 0.0001 |
| **Ozone depletion** | 0.73 (0.37) | 0.70 (0.31) | 0.67 (0.29) | 0.71 (0.33) | 0.62 (0.27) | 0.0004 | 0.012 (0.003 ─ 0.022) | 0.0010 |
| **Ionizing radiation** | 1.28 (0.39) | 1.18 (0.35) | 1.22 (0.36) | 1.11 (0.29) | 1.03 (0.28) | < 0.0001 | 0.054 (0.034 ─ 0.075) | < 0.0001 |
| **Photochemical ozone formation** | 16.34 (9.60) | 18.31 (10.55) | 18.51 (10.19) | 19.48 (10.19) | 19.04 (10.07) | 0.0004 | 0.012 (0.002 ─ 0.022) | < 0.0001 |
| **Particulate matter emissions** | 0.60 (0.22) | 0.58 (0.27) | 0.56 (0.23) | 0.52 (0.20) | 0.46 (0.18) | < 0.0001 | 0.039 (0.022 ─ 0.057) | < 0.0001 |
| **Acidification** | 0.08 (0.03) | 0.08 (0.04) | 0.08 (0.03) | 0.07 (0.03) | 0.06 (0.03) | < 0.0001 | 0.045 (0.026 ─ 0.063) | < 0.0001 |
| **Terrestrial eutrophication** | 26.50 (9.13) | 26.54 (12.41) | 25.58 (10.03) | 24.01 (8.23) | 22.65 (8.05) | < 0.0001 | 0.019 (0.007 ─ 0.032) | < 0.0001 |
| **Freshwater eutrophication** | 0.34 (0.13) | 0.32 (0.16) | 0.31 (0.14) | 0.28 (0.11) | 0.23 (0.10) | < 0.0001 | 0.065 (0.043 ─ 0.087) | < 0.0001 |
| **Marine eutrophication** | 0.85 (0.26) | 0.80 (0.28) | 0.81 (0.29) | 0.76 (0.29) | 0.71 (0.28) | < 0.0001 | 0.025 (0.011 ─ 0.040) | < 0.0001 |
| **Land use** | 82.10 (27.52) | 75.79 (26.39) | 77.16 (26.76) | 71.54 (24.09) | 65.39 (21.88) | < 0.0001 | 0.040 (0.023 ─ 0.058) | < 0.0001 |
| **Freshwater ecotoxicity** | 373.42 (145.64) | 346.84 (187.81) | 342.20 (170.36) | 295.19 (136.60) | 247.55 (112.28) | < 0.0001 | 0.064 (0.042 ─ 0.085) | < 0.0001 |
| **Water use** | 7.21 (4.07) | 7.59 (4.28) | 8.51 (4.80) | 9.44 (4.56) | 8.28 (3.93) | < 0.0001 | 0.028 (0.013 ─ 0.043) | < 0.0001 |
| **Fossils resource** | 58.55 (18.20) | 56.72 (17.12) | 58.03 (16.55) | 55.19 (14.74) | 51.57 (13.60) | < 0.0001 | 0.019 (0.007 ─ 0.031) | < 0.0001 |
| **Metals and minerals resource** | 31.91 (11.63) | 30.88 (11.57) | 30.98 (13.39) | 30.32 (10.87) | 27.41 (9.74) | 0.0001 | 0.014 (0.004 ─ 0.025) | < 0.0001 |
| **Ratio PANDiet/PEF** | 0.88 (0.25) | 0.97 (0.33) | 0.99 (0.33) | 1.03 (0.35) | 1.18 (0.34) | < 0.0001 | 0.076 (0.053 ─ 0.100) | < 0.0001 |

*Note*. Data presented as mean (standard deviation). Q = quintile.

**Table S28. Environmental impact across quartiles of HSDI**

| | Q1 | Q2 | Q3 | Q4 | p | η2 (95%CI) | Trend |
|---|---|---|---|---|---|---|---|
| **Product Environmental Footprint** | 0.75 (0.25) | 0.74 (0.27) | 0.71 (0.24) | 0.69 (0.25) | 0.0052 | 0.007 (0.001 — 0.016) | < 0.0001 |
| **Greenhouse gas emission** | 5.78 (2.16) | 5.75 (2.49) | 5.39 (2.08) | 5.25 (2.23) | 0.0016 | 0.009 (0.001 — 0.018) | < 0.0001 |
| **Ozone depletion** | 0.72 (0.33) | 0.71 (0.32) | 0.67 (0.32) | 0.60 (0.30) | < 0.0001 | 0.018 (0.006 — 0.030) | < 0.0001 |
| **Ionizing radiation** | 1.20 (0.33) | 1.19 (0.37) | 1.17 (0.39) | 1.14 (0.37) | 0.1002 | | |
| **Photochemical ozone formation** | 19.57 (10.72) | 18.29 (10.61) | 16.87 (10.09) | 15.46 (7.09) | < 0.0001 | 0.022 (0.009 — 0.036) | < 0.0001 |
| **Particulate matter emissions** | 0.58 (0.23) | 0.56 (0.25) | 0.54 (0.23) | 0.52 (0.23) | 0.0007 | 0.010 (0.002 — 0.019) | < 0.0001 |
| **Acidification** | 0.08 (0.03) | 0.08 (0.04) | 0.07 (0.03) | 0.07 (0.03) | 0.0004 | 0.011 (0.002 — 0.021) | < 0.0001 |
| **Terrestrial eutrophication** | 26.60 (11.32) | 25.51 (9.53) | 24.29 (9.00) | 23.85 (9.13) | 0.0002 | 0.011 (0.003 — 0.022) | < 0.0001 |
| **Freshwater eutrophication** | 0.32 (0.13) | 0.31 (0.15) | 0.30 (0.13) | 0.29 (0.14) | 0.0090 | 0.007 (0.001 — 0.015) | < 0.0001 |
| **Marine eutrophication** | 0.82 (0.28) | 0.79 (0.26) | 0.79 (0.33) | 0.77 (0.27) | 0.0244 | 0.005 (0.000 — 0.013) | < 0.0001 |
| **Land use** | 76.59 (25.48) | 75.39 (24.51) | 73.69 (27.80) | 75.69 (29.81) | 0.4707 | | |
| **Freshwater ecotoxicity** | 333.64 (156.38) | 339.47 (173.20) | 323.42 (153.64) | 324.19 (174.56) | 0.4794 | | |
| **Water use** | 7.41 (4.13) | 8.67 (4.48) | 8.10 (4.31) | 8.30 (4.71) | < 0.0001 | 0.014 (0.004 — 0.026) | < 0.0001 |
| **Fossils resource** | 57.94 (16.27) | 57.01 (16.99) | 54.92 (17.45) | 53.79 (15.99) | 0.0014 | 0.009 (0.002 — 0.019) | < 0.0001 |
| **Metals and minerals resource** | 31.41 (12.44) | 31.26 (11.30) | 29.36 (10.74) | 28.85 (11.16) | 0.0025 | 0.008 (0.001 — 0.018) | < 0.0001 |
| **Ratio PANDiet/PEF** | 0.94 (0.28) | 0.98 (0.33) | 1.03 (0.34) | 1.08 (0.40) | < 0.0001 | 0.026 (0.012 — 0.041) | < 0.0001 |

*Note*. Data presented as mean (standard deviation). Q = quartile.

| Table S29. Environmental impact across quartiles of ELDS | | | | | | | |
|---|---|---|---|---|---|---|---|
| | **Q1** | **Q2** | **Q3** | **Q4** | **p** | **η2 (95%CI)** | **Trend** |
| **Product Environmental Footprint** | 0.80 (0.27) | 0.75 (0.27) | 0.67 (0.20) | 0.64 (0.21) | < 0.0001 | 0.062 (0.041 – 0.084) | < 0.0001 |
| **Greenhouse gas emission** | 6.30 (2.47) | 5.77 (2.33) | 5.05 (1.73) | 4.76 (1.79) | < 0.0001 | 0.074 (0.051 – 0.097) | < 0.0001 |
| **Ozone depletion** | 0.74 (0.36) | 0.72 (0.32) | 0.65 (0.28) | 0.58 (0.25) | < 0.0001 | 0.035 (0.019 – 0.052) | < 0.0001 |
| **Ionizing radiation** | 1.25 (0.38) | 1.22 (0.38) | 1.12 (0.31) | 1.07 (0.30) | < 0.0001 | 0.042 (0.025 – 0.061) | < 0.0001 |
| **Photochemical ozone formation** | 19.94 (11.13) | 18.80 (10.92) | 16.39 (9.03) | 15.54 (7.22) | < 0.0001 | 0.030 (0.015 – 0.047) | < 0.0001 |
| **Particulate matter emissions** | 0.63 (0.25) | 0.58 (0.24) | 0.50 (0.19) | 0.46 (0.19) | < 0.0001 | 0.071 (0.049 – 0.094) | < 0.0001 |
| **Acidification** | 0.09 (0.04) | 0.08 (0.03) | 0.07 (0.03) | 0.06 (0.03) | < 0.0001 | 0.073 (0.050 – 0.096) | < 0.0001 |
| **Terrestrial eutrophication** | 28.26 (11.64) | 25.77 (10.15) | 22.99 (7.69) | 22.49 (8.02) | < 0.0001 | 0.055 (0.035 – 0.076) | < 0.0001 |
| **Freshwater eutrophication** | 0.35 (0.15) | 0.31 (0.14) | 0.27 (0.11) | 0.25 (0.11) | < 0.0001 | 0.071 (0.049 – 0.094) | < 0.0001 |
| **Marine eutrophication** | 0.86 (0.27) | 0.81 (0.28) | 0.75 (0.30) | 0.73 (0.25) | < 0.0001 | 0.034 (0.019 – 0.052) | < 0.0001 |
| **Land use** | 81.28 (27.61) | 76.71 (25.75) | 70.04 (23.80) | 69.63 (25.17) | < 0.0001 | 0.035 (0.019 – 0.053) | < 0.0001 |
| **Freshwater ecotoxicity** | 370.39 (173.91) | 337.17 (168.37) | 300.11 (133.44) | 286.67 (149.41) | < 0.0001 | 0.042 (0.024 – 0.060) | < 0.0001 |
| **Water use** | 7.67 (4.19) | 8.18 (4.97) | 7.96 (3.67) | 8.47 (4.60) | 0.0492 | 0.005 (0.000 – 0.012) | 0.0002 |
| **Fossils resource** | 60.23 (18.18) | 58.24 (17.29) | 52.83 (13.72) | 51.12 (13.44) | < 0.0001 | 0.049 (0.031 – 0.069) | < 0.0001 |
| **Metals and minerals resource** | 32.63 (12.53) | 32.00 (12.74) | 28.48 (9.67) | 27.20 (9.29) | < 0.0001 | 0.037 (0.020 – 0.054) | < 0.0001 |
| **Ratio PANDiet/PEF** | 0.88 (0.27) | 0.97 (0.31) | 1.06 (0.34) | 1.14 (0.37) | < 0.0001 | 0.089 (0.064 – 0.113) | < 0.0001 |

*Note*. Data presented as mean (standard deviation). Q = quartile.
